# Association between plausible genetic factors and weight loss from GLP1-RA and bariatric surgery: a multi-ancestry study in 10 960 individuals from 9 biobanks

**DOI:** 10.1101/2024.09.11.24313458

**Authors:** Jakob German, Mattia Cordioli, Veronica Tozzo, Sarah Urbut, Kadri Arumäe, Roelof A.J. Smit, Jiwoo Lee, Josephine H. Li, Adrian Janucik, Yi Ding, Akintunde Akinkuolie, Henrike Heyne, Andrea Eoli, Chadi Saad, Yasser Al-Sarraj, Rania Abdel-latif, Shaban Mohammed, Moza Al Hail, Alexandra Barry, Zhe Wang, Estonian Biobank research team, Tatiana Cajuso, Andrea Corbetta, Pradeep Natarajan, Samuli Ripatti, Anthony Philippakis, Lukasz Szczerbinski, Bogdan Pasaniuc, Zoltan Kutalik, Hamdi Mbarek, Ruth J.F. Loos, Uku Vainik, Andrea Ganna

## Abstract

Obesity is a significant public health concern. GLP-1 receptor agonists (GLP1-RA), predominantly in use as a type 2 diabetes treatment, are a promising pharmacological approach for weight loss, while bariatric surgery (BS) remains a durable, but invasive, intervention. Despite observed heterogeneity in weight loss effects, the genetic effects on weight loss from GLP1-RA and BS have not been extensively explored in large sample sizes, and most studies have focused on differences in race and ethnicity, rather than genetic ancestry. We studied whether genetic factors, previously shown to affect body weight, impact weight loss due to GLP1-RA therapy or BS in 10,960 individuals from 9 multi-ancestry biobank studies in 6 countries. The average weight change between 6 and 12 months from therapy initiation was -3.93% for GLP1-RA users, with marginal differences across genetic ancestries. For BS patients the weight change between 6 and 48 months from the operation was -21.17%. There were no significant associations between weight loss due to GLP1-RA and polygenic scores for BMI or type 2 diabetes or specific missense variants in the *GLP1R, PCSK1 and APOE* genes, after multiple-testing correction. A higher polygenic score for BMI was significantly linked to lower weight loss after BS (+0.7% for 1 standard deviation change in the polygenic score, P = 1.24×10^-4^), but the effect was modest and further reduced in sensitivity analyses. Our findings suggest that existing polygenic scores related to weight and type 2 diabetes and missense variants in the drug target gene do not have a large impact on GLP1-RA effectiveness. Our results also confirm the effectiveness of these treatments across all major continental ancestry groups considered.

## Introduction

The obesity epidemic presents a significant public health burden, driving the need for effective treatment options.^1,2^ Despite 75% of adults with obesity having attempted to lose weight, most have not achieved lasting success.^3^ Amid these challenges, glucagon-like peptide-1 receptor agonists (GLP1-RA) have emerged as a promising solution. As first discovered in the late 1980s, glucagon-like peptide-1 (GLP-1) is linked to the incretin effect, which promotes insulin release from pancreatic beta cells, thus playing a crucial role in regulating blood sugar levels.^4,5^ This evidence led to the approval of the first GLP1-RA for treatment of type 2 diabetes in 2005.^6–8^ Nine years later, an association between GLP-1 and reduced food intake paved the way for the development and approval of the first GLP1-RA for obesity treatment.^5,9^

Clinical trials conducted in the following years demonstrated weight loss effects ranging from 6% to 16% for liraglutide and semaglutide, leading to a substantial increase in GLP1-RA usage worldwide^10–12^. Additional trials have further revealed cardiovascular^13,14^ and renal benefits^15^, with ongoing trials currently underway.^16^ Overall, anticipated trends suggest that by 2030, the US will witness approximately 30 million patients using GLP1-RA drugs, representing roughly 9% of its population, underscoring their potential in combating the obesity crisis and exploring new therapeutic applications.^17–19^

Meanwhile, bariatric surgery (BS) stands as a durable and effective treatment for individuals with severe obesity (insurance payer threshold in the US: BMI ≥ 40 kg/m²)^20,21^ and metabolic comorbidities (BMI ≥ 35 kg/m²), with some clinical guidelines now recommending its use for patients with a BMI ≥ 30 kg/m².^22,23^ Meta-analyses combining results from randomized controlled trials and observational studies found that, on average, individuals undergoing BS experienced a weight loss of 26 kg. Separately, a randomized controlled trial reported a total weight loss of 25% after 5 years in patients undergoing sleeve gastrectomy and Roux-en-Y gastric bypass.^24,25^ Although metabolic surgery carries typical risks for such interventions, including nutrient malabsorption, large-scale observational studies have demonstrated remarkable long-term benefits, such as reduced cardiovascular outcomes, lower all-cause mortality, and higher remission rates of type 2 diabetes, hypertension, and hyperlipidemia.^26–32^

Despite the robust weight loss effects observed with both GLP1-RA treatment and BS, there exists heterogeneity in their effectiveness. One out of every five patients undergoing BS may not achieve the desired weight loss within the first year or may experience weight regain within two years.^33–36^ The variability of GLP1-RA treatment effects on glycemic outcomes has been widely investigated and can be partially explained by clinical indicators like low β-cell function or C-peptide levels as well as genetic variants.^37–42^ However, with the exception of type 2 diabetes, sex, and baseline weight, the factors influencing the diversity in weight loss effects of these drugs are still poorly understood.^43–45^ Additionally, differences in patient adherence to treatment regimens may further contribute to this variability.^46^

Genetics has been considered a potential factor for heterogeneity in weight loss treatments, but its role remains largely unexplored.^43^ A previous study has examined the role of a common missense variant in the *GLP1R* genes in 57 women and found a weak link with GLP1-RA treatment response.^39,47^ A greater number and better-powered studies have been conducted in individuals with BS ^48,49^, however, a consistent genome-wide association for weight loss has yet to be identified. A previous study has shown that a polygenic score for BMI had a significant impact on weight loss after biliopancreatic diversion with duodenal switch in 865 patients.^50^

Finally, there has been limited exploration into the effectiveness of these obesity treatments across different genetic ancestries and across various healthcare settings.^10,51^

In our study we seek to i) characterize the observed GLP1-RA and BS body weight-lowering effects using real-world data from 10 960 individuals across 9 biobanks in 6 countries, with 6 major continental ancestry groups; ii) identify whether plausible genetic factors (i.e. polygenic scores for BMI and T2D and coding variants in *GLP1R*, *PCSK1* and *APOE* gene) associate with heterogeneity in GLP1-RA and BS weight loss effects, and thus identify patient groups that will benefit most from such treatments; iii) compare the effects of GLP1-RA treatment and BS on weight loss across major continental ancestry groups.

## Results

### GLP1-RA and bariatric surgery associated weight loss in 10 960 individuals from 9 cohorts and 6 major continental ancestry groups

We investigated changes in body weight associated with GLP1-RA treatment and BS across 9 biobank studies: Helsinki University Hospital (HUS, Finland; N = 633), Estonian Biobank (ESTBB; N = 464), UK Biobank (UKBB, United Kingdom; N = 810), All of Us (AoU, USA; N = 559), BioMe Biobank (BioMe, USA; N = 2,170), Mass General Brigham Biobank (MGBB, USA; N = 2,141), Atlas Biobank (UCLA-ATLAS, USA; N = 1,445), Qatar Biobank (QBB, Qatar; N = 2,383), Bialystok Bariatric Surgery Study (BBSS, Poland, N = 355). We included individuals that were at least 18 years of age when starting a GLP1-RA treatment or undergoing BS. Individuals undergo GLP1-RA treatment for different medical reasons, not only for weight loss. We defined GLP1-RA usage based on prescription or purchase data (ATC codes starting with A10B*; including exenatide, liraglutide, lixisenatide, albiglutide, dulaglutide, semaglutide, and beinaglutide) covering at least 12 months, and excluding individuals with indication of treatment discontinuation within this time window. For BS, we considered individuals undergoing the following procedures: Roux-en-Y gastric bypass, sleeve gastrectomy, adjustable gastric band, vertical banded gastroplasty, biliopancreatic diversion with duodenal switch. More information can be found in the **Methods** section.

We included only individuals who had an initial body weight measurement recorded at most 12 months prior to the baseline (defined as treatment initiation or surgery) and a follow-up measurement taken at least 6 but no more than 12 months after the baseline for GLP1-RA treatments, or within 48 months after baseline for BS. In case of multiple measurements within this time frame, we considered the median of such measurements (**Supplementary Figure 1** for a schematic of the approach used). We defined our main outcome of interest as the percentage change in body weight, calculated as the difference between the second body weight measurement (or median of multiple measurements) and the initial one, divided by the initial measurement.

Our study population included a total of 6,750 GLP1-RA users and 4,210 individuals undergoing BS (baseline characteristics are reported in **Table 1**). Overall, GLP1-RA users 1) had a lower proportion of women (61.0% weighted average across studies) when compared to BS patients (73.1% weighted average across studies), 2) were on average older (57.30 *vs* 46.50), and 3) had a lower initial body weight (99.80 kg *vs* 116.96 kg). The proportion of individuals diagnosed with type 2 diabetes at time of GLP1-RA initiation was at least 48% across all studies and major continental ancestry groups.

**Table 1.** Baseline characteristics for 6 750 GLP1-RA users and 4 210 patients undergoing bariatric surgery included in our study population. For each cohort and major continental ancestry group (AFR: African, AMR: admixed American, EAS: East Asian, EUR: European, MID: Middle Eastern, SAS: South Asian), we report the total number of individuals eligible for analysis, the proportion of females, the mean (SD) body weight (in Kg) at baseline, the mean (SD) age (in years) at baseline, the prevalence of T2D, and (for GLP1-RA users only) the most commonly used type of medication and the proportion of semaglutide users. HUS, Helsinki University Hospital; ESTBB, Estonian Biobank; UKBB, UK Biobank; AoU, All of US; MGBB, Mass General Brigham Biobank; BioMe, BioMe; UCLA-ATLAS, Atlas Biobank; QBB, Qatar Biobank; BBSS, Bialystok Bariatric Surgery Study

| Study | Ancestry | Number of individuals | Proportion of women (%) | Mean body weight (in kg) at baseline (SD) | Mean BMI (in kg/m<sup>2</sup>) at baseline (SD) | Mean age at baseline (SD) | T2D prevalence (%) | Most common GLP1-RA type | Proportion of semaglutide (%) |
| --- | --- | --- | --- | --- | --- | --- | --- | --- | --- |
| **GLP1-RA** |  |  |  |  |  |  |  |  |  |
| HUS | EUR | 255 | 68.00 | 113.70 (22.90) | 39.56 (7.93) | 53.79 (13.52) | 65.49 | semaglutide | 60.40 |
| ESTBB | EUR | 191 | 62.30 | 109.18 (20.30) | 37.68 (6.40) | 56.95 (10.96) | 89.01 | semaglutide | 72.77 |
| UKBB | EUR | 614 | 44.30 | 105.00 (19.90) | 36.70 (6.14) | 60.50 (7.15) | 99.80 | liraglutide | 0.00 |
| AoU | EUR | 117 | 64.96 | 106.66 (24.18) | 37.23 (7.84) | 57.01 (11.03) | 82.91 | liraglutide | 11.97 |
|  | AFR | 91 | 83.52 | 104.8 (25.01) | 37.94 (8.77) | 54.57 (11.93) | 92.31 | liraglutide | 18.68 |
| MGBB | EUR | 857 | 57.01 | 99.99 (21.48) | 35.26 (6.79) | 59.88 (12.17) | 56.33 | semaglutide | 40.72 |
|  | AFR | 156 | 54.32 | 104.65 (23.80) | 37.25 (7.85) | 54.31 (11.91) | 60.24 | dulaglutide | 36.14 |
| BioMe | AMR | 614 | 66.45 | 91.19 (22.45) | 34.34 (7.19) | 59.07 (11.29) | 67.75 | dulaglutide | 29.32 |
|  | AFR | 728 | 69.78 | 101.49 (25.42) | 36.15 (8.18) | 57.00 (12.24) | 69.50 | dulaglutide | 29.12 |
|  | EUR | 369 | 44.44 | 98.26 (21.93) | 33.93 (6.74) | 60.32 (11.57) | 47.97 | semaglutide | 37.13 |
|  | SAS | 77 | 50.65 | 83.79 (20.10) | 30.61 (6.70) | 56.48 (12.59) | 70.13 | semaglutide | 41.56 |
| UCLA-ATLAS | AFR | 133 | 64.00 | 101.6 (23.20) | 35.50 (7.10) | 61.00 (13.00) | 91.00 | semaglutide | 57.10 |
|  | AMR | 336 | 66.00 | 94.40 (22.60) | 35.10 (7.10) | 56.00 (13.00) | 84.00 | liraglutide | 26.50 |
|  | EAS | 120 | 47.00 | 81.50 (16.30) | 29.70 (5.20) | 60.00 (13.00) | 92.00 | semaglutide | 61.70 |
|  | EUR | 856 | 53.00 | 101.60 (22.00) | 34.80 (6.90) | 63.00 (13.00) | 80.00 | semaglutide | 67.80 |
| QBB | MID | 1236 | 72.17 | 97.68 (24.54) | 37.67 (8.90) | 49.52 (10.75) | 75.97 | liraglutide | 3.64 |
| **Bariatric surgery** |  |  |  |  |  |  |  |  |  |
| HUS | EUR | 378 | 81.00 | 113.63 (19.50) | 40.00 (5.71) | 49.90 (9.39) | 37.04 |  |  |
| ESTBB | EUR | 273 | 76.56 | 121.24 (20.57) | 41.80 (5.58) | 44.72 (10.07) | 28.21 |  |  |
| UKBB | EUR | 196 | 69.40 | 121.00 (29.70) | 44.20 (9.74) | 55.50 (8.21) | 50.00 |  |  |
| AoU | EUR | 168 | 81.55 | 121.32 (29.88) | 43.48 (9.85) | 50.74 (12.62) | 18.45 |  |  |
|  | AFR | 183 | 89.62 | 127.23 (32.55) | 49.33 (27.56) | 46.09 (11.67) | 48.63 |  |  |
| MGBB | EUR | 978 | 71.06 | 121.21 (26.52) | 42.93 (7.90) | 49.37 (12.16) | 39.98 |  |  |
|  | AFR | 150 | 86.67 | 122.89 (23.92) | 45.16 (8.27) | 43.68 (11.67) | 42.67 |  |  |
| BioMe | AMR | 143 | 79.72 | 112.36 (20.81) | 42.55 (6.13) | 44.62 (11.48) | 30.07 |  |  |
|  | AFR | 188 | 82.98 | 124.88 (25.63) | 45.45 (7.90) | 45.35 (12.16) | 29.26 |  |  |
|  | EUR | 51 | 70.59 | 120.23 (25.29) | 43.03 (7.95) | 49.51 (13.34) | 17.65 |  |  |
| QBB | MID | 1147 | 69.49 | 102.14 (23.18) | 38.55 (7.87) | 41.66 (10.66) | 40.02 |  |  |
| BBSS | EUR | 355 | 56.06 | 138.48 (27.07) | 47.10 (7.11) | 47.10 (7.11) | 35.21 |  |  |

Among GLP1-RA users, we observed an average body weight change (**Figure 1**) across studies of -3.93% (ranging from -1.08% to -7.07%), in line with results from observational studies.^52,53^ Among BS patients, we observed an average change of -21.17% (-15.00% to -27.72%).

**Figure 1.**
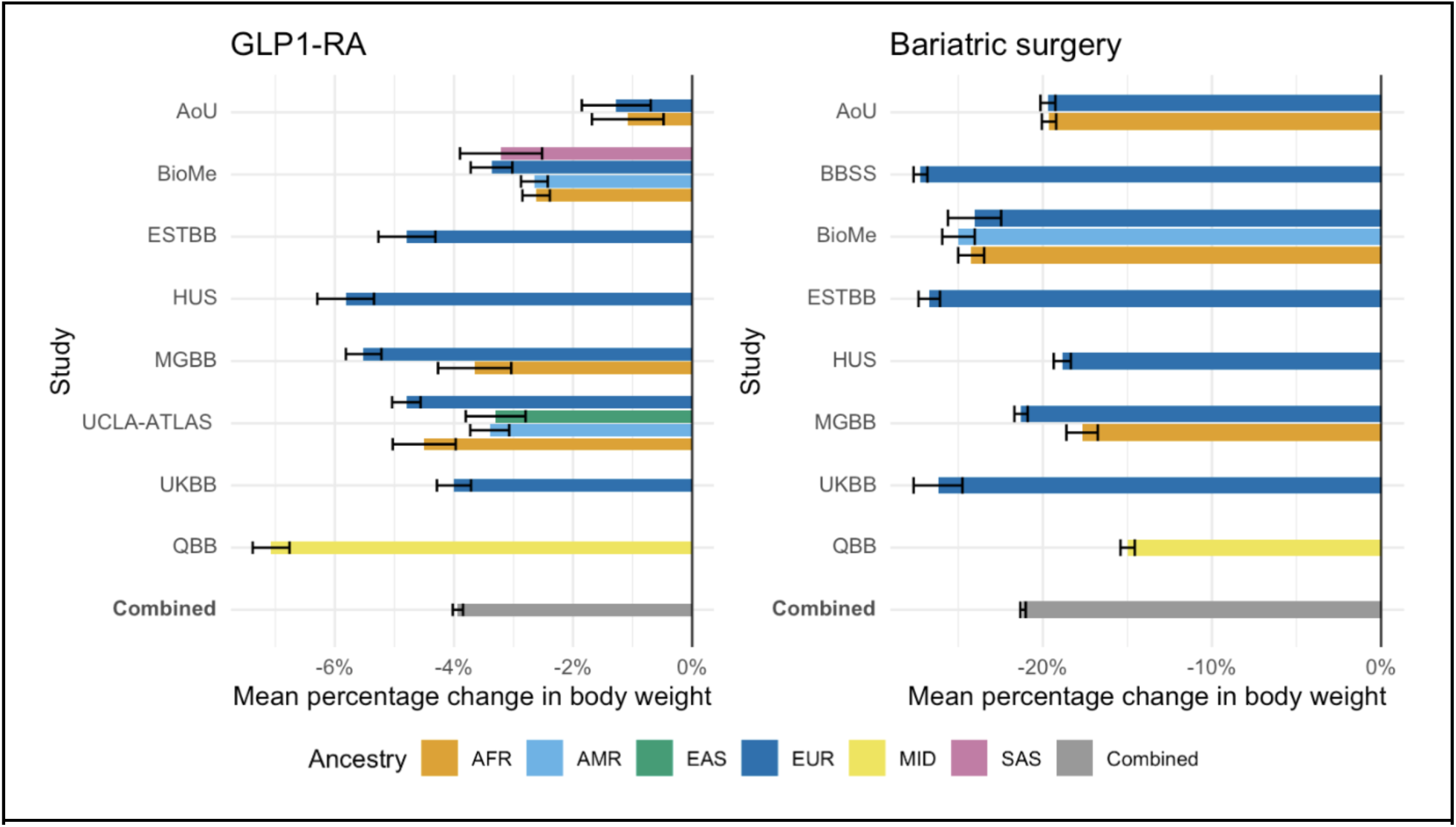
Average percentage change in body weight for 6 750 GLP1-RA users and 4 210 bariatric surgery patients. Bars represent the mean percentage change in body weight within each study and ancestry, and the overall combined weighted mean change. Error bars represent standard errors.

### Demographic factors associated with GLP1-RA and bariatric surgery weight change

We explored the effect of baseline characteristics on body weight changes associated with GLP1-RA treatment and BS. We fitted a multivariable linear model with percentage change in body weight as outcome and baseline body weight, sex, age at baseline, the first 20 principal genetic components and medication type (for GLP1-RA users only) as predictors. Each cohort was analyzed separately, stratifying by major continental ancestry group, and the effects were then meta-analyzed (**Table 2**). Baseline body weight was significantly associated with greater weight loss for both GLP1-RA and BS treatments (*β_GLP1-RA_* = -0.05 % weight change compared to baseline, *P* = 1.63 × 10^-35^; *β_BS_* = -0.14 % weight change compared to baseline, *P* = 5.49 × 10^-69^) and women had a significantly greater weight loss for both treatments (*β_GLP1-RA_* = -1.54 % weight change compared to baseline, *P* = 2.47 × 10^-16^; *β_BS_* = -2.66 % weight change compared to baseline, *P* = 6.83 × 10^-10^). Being older at treatment initiation was only associated with less weight loss for bariatric surgery (*β_BS_* = 0.15 % weight change compared to baseline, *P* = 2.54 × 10^-15^). The effects of baseline weight and sex on weight loss were highly heterogeneous across ancestries and studies (I^2^ for GLP1-RA = 0.99 and 0.83 for baseline weight and sex, respectively) (**Supplementary Figures 2-4 and Supplementary Tables 1-2**).

**Table 2.**
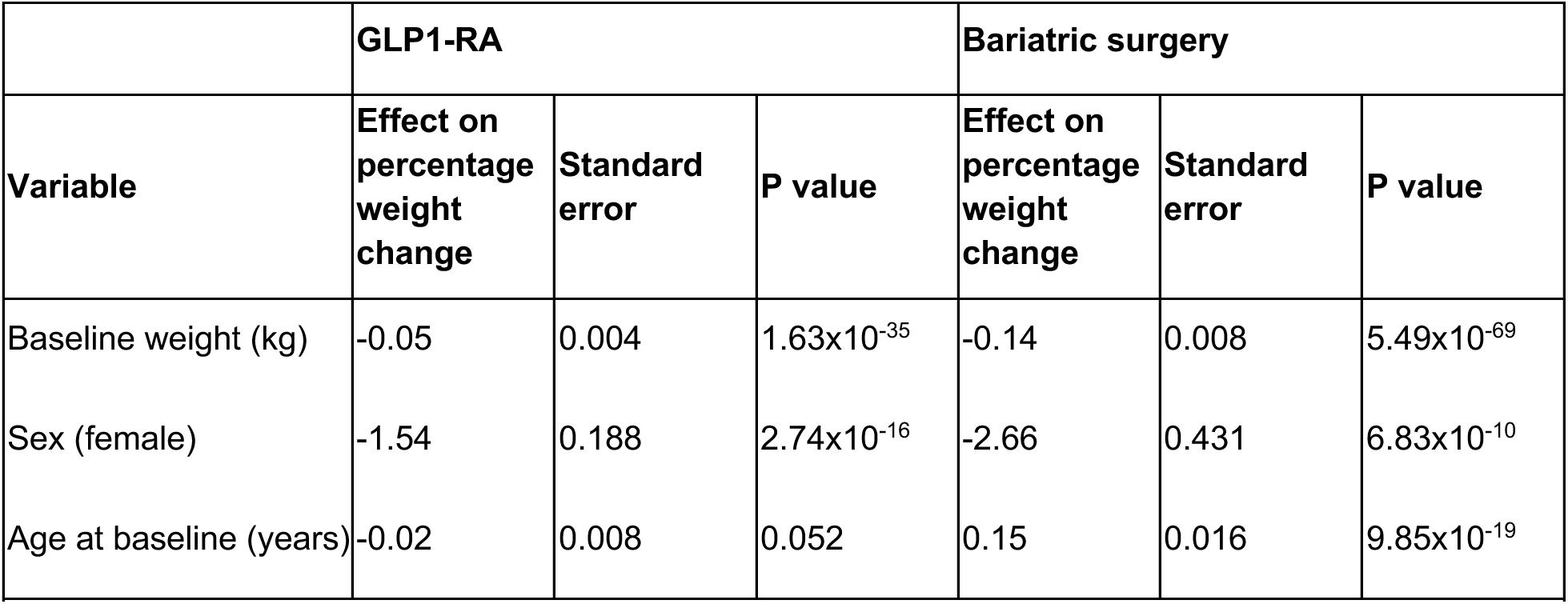
Effect of baseline weight, sex and age at baseline of percentage weight changes associated with GLP1-RA and bariatric surgery. Multivariable model adjusting for the three variables described in the table and the first 20 principal genetic components and medication type (for GLP1-RA users only). Negative effects indicate larger weight loss.

|  | GLP1-RA |  |  | Bariatric surgery |  |  |
| --- | --- | --- | --- | --- | --- | --- |
| Variable | Effect on percentage weight change | Standard error | P value | Effect on percentage weight change | Standard error | P value |
| Baseline weight (kg) | -0.05 | 0.004 | $1.63 \times 10^{-35}$ | -0.14 | 0.008 | $5.49 \times 10^{-69}$ |
| Sex (female) | -1.54 | 0.188 | $2.74 \times 10^{-16}$ | -2.66 | 0.431 | $6.83 \times 10^{-10}$ |
| Age at baseline (years) | -0.02 | 0.008 | 0.052 | 0.15 | 0.016 | $9.85 \times 10^{-19}$ |

### Marginal effect of ancestry on GLP1-RA and bariatric surgery associated weight loss

We evaluate whether the effect of GLP1-RA and BS associated weight loss was significantly different across genetically-defined continental ancestry groups after accounting for differences in weight at baseline, age, sex and, only for GLP1-RA, medication type (**Table 3**). Only multi-ancestry biobanks (AoU, BioMe, MGBB, UCLA-ATLAS) were included in this analysis (N = 6 315). Compared to individuals of European ancestry, all other ancestry groups had lower change in body weight. However, this effect was statistically significant only among GLP1-RA users of African (*β* = 0.76 % weight change from baseline weight, *P* = 0.02) and admixed American (*β* = 0.87 % weight change from baseline weight, *P* = 3 × 10^-3^) ancestries.

**Table 3.** Associations between weight change and major continental ancestry groups. Meta-analysis effect sizes, standard errors and P values for the association between weight change and major continental ancestry groups (AFR: African, AMR: admixed American, EAS: East Asian, EUR: European, SAS: South Asian). Coefficients are derived from a multivariable model including percentage weight change as outcome and ancestry as categorical variable (with EUR as reference category), and adjusting for sex, age and weight at baseline and, for GLP1-RA only, medication type. Negative effects indicate larger weight loss

| GLP1-RA |  |  |  |
| --- | --- | --- | --- |
| Major continental ancestry group | Effect on percentage weight change | Standard error | P value |
| AFR | 0.758 | 0.327 | 0.021 |
| AMR | 0.874 | 0.295 | 0.003 |
| EAS | 1.093 | 0.664 | 0.100 |
| SAS | 0.118 | 0.759 | 0.876 |
| Bariatric surgery |  |  |  |
| Major continental ancestry group | Effect on percentage weight change | Standard error | P value |
| AFR | 1.688 | 1.308 | 0.197 |
| AMR | -0.542 | 1.428 | 0.704 |

### Plausible genetic factors do not associate with weight change due to GLP1-RA

We investigated the effect of genetic exposures with plausible effects on body weight changes. Genetic exposures were selected upon discussion between authors and before initiating analyses. First, we considered polygenic scores for BMI based on previous observation that a polygenic score was associated with BMI trajectories.^54,55^ We also consider the only replicated genome-wide variant associated with BMI change in the largest study of BMI trajectories (*rs429358* in *APOE*).^54^ Second, we considered a polygenic score for T2D based on the observation that biomarkers of type 2 diabetes were associated with effectiveness of GLP1-RA.^38,40^ We confirmed that both polygenic scores for BMI and T2Ds were significantly associated with their respective traits across all major continental ancestry groups (**Supplementary Table 3**) although the effects were larger in individuals of European ancestry.

Third, we considered 13 SNPs in the *GLP1R* and *PCSK1* genes. We considered all missense variants in the *GLP1R* gene with at least 1% minor allele frequency in at least one of major continental ancestry group, as per gnomAD (version 4.1)^56^, three of which have been functional characterized as gain or loss of function (**Supplementary Table 4**).^57^ We also considered two non-synonymous variants in the *PCSK1* gene that have been associated with BMI variation in the population.^58^ Finally, we sought to replicate the effect of two variants associated with weight loss in one of the largest studies of BS^49^(**Methods**). After analyses, two missense variants in the *GLP1R* genes, *rs2295006* and *rs201672448* were too rare to be tested across studies and were not considered further.

We estimated the effect of each genetic exposure on weight change in each cohort and major continental ancestry groups separately, fitting a linear model adjusted for baseline body weight, sex, age at baseline, first 20 principal genetic components, and medication type (for GLP1-RA users only), and we subsequently combined the effects through ancestry-specific and multi-ancestry meta-analyses (**Figure 2, Supplementary Tables 5-6**). We did not adjust for T2D diagnosis because we considered it a potential mediator between the genetic exposure and weight change outcome.

**Figure 2.**
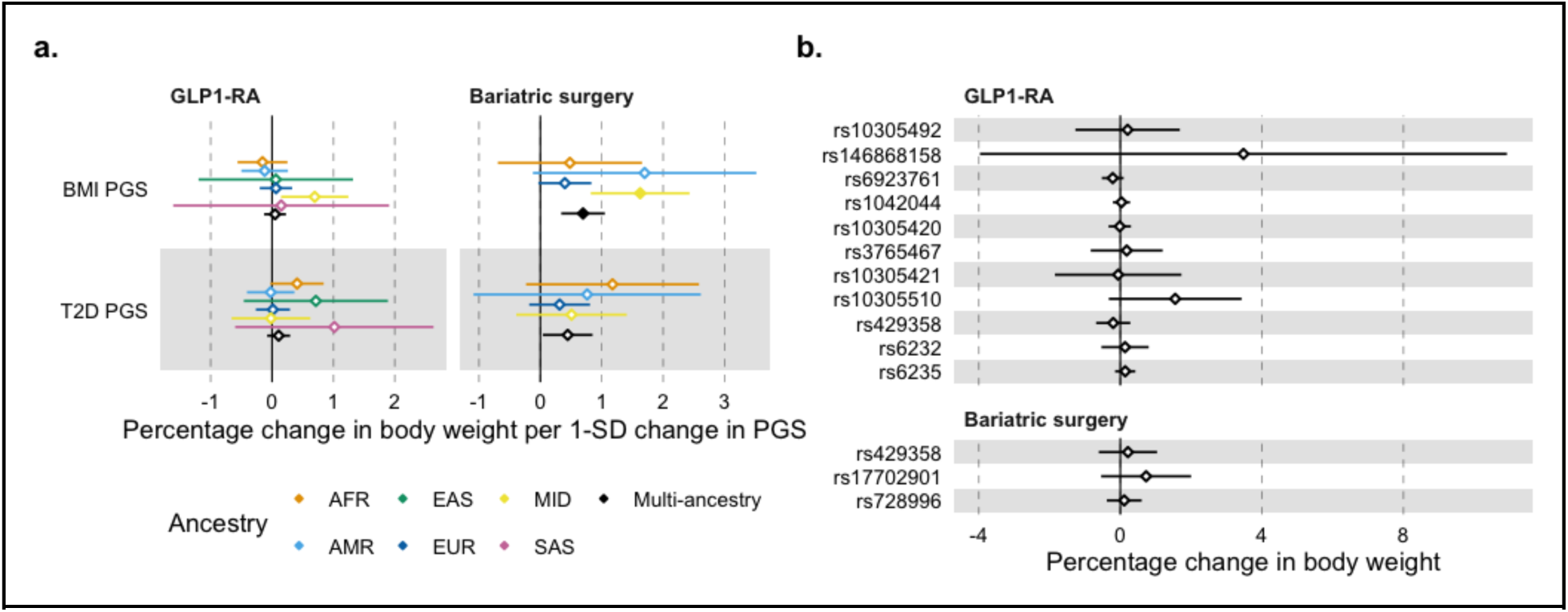
Effect of 15 genetic exposures on body weight changes associated with GLP1-RA treatment and bariatric surgery. a. Ancestry-specific and multi-ancestry meta-analysis effect sizes for association between percentage change in body weight and PGS for BMI and type 2 diabetes. Dots represent the percentage change in body weight per one standard deviation change in PGS, error bars represent the 95% confidence interval. Full dots represent statistical significance at P < 0.004. b. Multi-ancestry meta-analysis effect sizes for the association between percentage change in body weight and genotype at each locus. Dots represent the effect size, error bars represent the 95% confidence interval. PGS, polygenic score.

Among GLP1-RA users, we did not observe any significant genetic exposure associated with weight loss after multiple testing correction (*P* < 0.004, Bonferroni correction for 13 exposures tested).

We further tested whether the lack of associations observed was possibly due to high effect-size heterogeneity across ancestries, or due to lack of statistical power. We did not observe any significant heterogeneity across ancestries for any of the genetic exposure (average I^2^ across genetic exposures for GLP1-RA was 0.06 **-Supplementary Tables 7-8**). Moreover, given the current sample size for GLP1-RA users (N = 6 750) our power calculation showed we would have at least 80% power to detect a statistically significant effect (at *P* < 0.05) of at least a 0.3% change in body weight for 1 standard deviation (SD) change in the polygenic score, or for SNPs with a minor allele frequency of at least 1% (**Methods**).

### A higher polygenic score for BMI associates with lower weight loss due to BS

Among BS patients, a higher polygenic score for BMI was significantly associated with lower weight loss (*β_BMI PGS_* = 0.70 % weight change compared to baseline for 1 SD change in the polygenic score, *P* = 1.24 × 10^-4^) after multiple testing correction (*P* < 0.01, Bonferroni correction for 5 exposures tested), in the opposite direction than observed for weight at baseline (**Figure 2**, **Table 2**). A higher polygenic score for T2D was nominally associated with lower weight loss (*β_T2D PGS_* = 0.45 % weight change compared to baseline for 1 SD change in the polygenic score, *P* = 0.03).

We observed significant heterogeneity in the association between BMI PGS and weight loss across studies (I2 = 0.64, **Supplementary Table 8**).

### Sensitivity analyses

To assess the robustness of our findings, we conducted a series of sensitivity analyses.

First, we consider different outcome definitions. We shortened the follow-up period for BS to 12 months to make it comparable to that of GLP1-RA, we considered only participants with weight measurements available at least 30 days prior to BS to account for the possibility of significant weight loss due to preoperative low-calorie diets and we used minimum rather than median weight during the follow-up period. This latter definition resulted in significant higher weight loss for GLP1-RA (-6.00% compared to -3.93% when using the median, **Supplementary Tables 9-10** and **Supplementary Figures 5-6**). All analyses confirmed the associations observed in primary analysis (**Supplementary Tables 11-19** and **Supplementary Figures 7-14**), although the effect of BMI PGS on weight loss due to BS was slightly reduced when considering a shortened follow-up period (**Supplementary Table 11** and **Supplementary Figure 8**).

Second, we considered different covariate adjustment and performed sub-group analyses. We adjusted the analyses for T2D status and observed similar results than primary analyses (**Supplementary Tables 12-13** and **Supplementary Figure 9)**. We considered only semaglutide and liraglutide as GLP1-RA treatments to capture their specific use for weight loss. While we observed a higher weight loss than considering all GLP1-RA treatments (-4.69% vs -3.93% **Supplementary Figure 10**), we confirmed the lack of effect of BMI and T2D PGS on weight loss observed in primary analyses (**Supplementary Table 14 and Supplementary Figure 11**).

Third, we considered different statistical models. Previous works have argued that using percentage changes in weight loss pre vs post therapy is statistically inappropriate and inefficient.^59–61^ Thus, we considered a model using post-treatment weight as outcome, including the same predictors as in the main model and an additional term modeling the interaction between baseline weight and each genetic exposure. We observed similar results as in the primary model (**Supplementary Tables 16-17**). A second potential issue with the primary model we used relates with adjusting longitudinal change phenotypes for the baseline trait.^62,63^ In particular, when testing for the effect of genetic exposure on drug response outcomes, adjusting phenotype changes (e.g. weight loss) for baseline levels (e.g. weight at treatment initiation) can result in false positive association when genetic variants are also associated with baseline levels^64^. Removing baseline weight from the model led to a reduction in the effect of the BMI PGS on weight loss due to BS (0.15% per 1 SD change in the polygenic score, P = 0.420 compared to 0.70% in the primary model; **Supplementary Tables 18-19** and **Supplementary Figure 14**).

## Discussion

In this study we provide a comprehensive assessment of major demographic factors and plausible genetic factors on body weight changes associated with GLP1-RA use and BS.

In line with previous work,^43,65–68^ our main outcome of interest was the percentage change in body weight between 6 and 12 months after GLP1-RA treatment initiation as well as between 6 and 48 months after BS.

By considering studies from 6 different countries, we were able to highlight the heterogeneity in use of GLP1-RA across different healthcare systems. GLP1-RA users consisted mostly of individuals with type 2 diabetes, reflecting their original use for treatment of this condition. Biobank studies covering Boston (MGBB), New York (BioMe), Los Angeles (UCLA-ATLAS) and Helsinki (HUS) had overall lower rates of type 2 diabetes individuals compared to the other studies and lower weight at baseline, which, together with the availability of more updated electronic health records data, suggest a larger number of individuals being prescribed GLP1-RA for weight loss. However, care should be taken in interpreting variation in rate of type 2 diabetes across biobanks as this might reflect different approaches used for capturing this diagnosis from the electronic health records as well as different sampling strategies of biobanks.

The comparison between GLP1-RA and BS is informative as it highlights the younger age, higher weight and lower percentage of men undergoing BS compared to GLP1-RA users. The weight reduction was also approximately 5 times higher among BS patients compared to GLP1-RA users. The weight reduction among GLP1-RA users was also more heterogeneous across biobanks compared to BS, but in line with what reported previously in the literature using real world data^41^ and lower than results from clinical trials.^11,69^ The lower weight reduction observed among GLP1-RA users is partially influenced by the use of the median postoperative or post-drug therapy weight, while we observed larger weight reduction when using minimum weight. However, our approach provides a more conservative and robust estimate, better accounting for the variability and potential uncertainty in clinical measurement data. Our findings also highlight a consistent effect of baseline body weight and sex on weight loss associated with both GLP1-RA treatment and BS, suggesting that individuals with higher weight at baseline, as well as women benefit more from both interventions. These findings underscore the importance of considering individual characteristics when designing weight loss interventions.

The main aim of this study was to evaluate the impact of genetic exposures on body weight changes induced by GLP1-RA or BS. At the current sample size, a genome-wide scan would have limited power to identify potential new signals. With 6,750 individuals, we would have at least 80% statistical power to identify, at a genome-wide significant threshold, a genetic variant with an allele frequency of at least 1% and an effect larger than 0.3% weight change per change in one SD, which is unlikely given previous effects of common variants on treatment response.^64^ Instead, we formulated a set of hypotheses based on the results from the largest genome-wide association studies of BMI and type 2 diabetes, and, explored biologically plausible mutations in several candidate genes. Polygenic scores represent the most powerful genetic predictors for BMI and type 2 diabetes and we confirm their significant association with the corresponding trait across all major continental ancestry groups. Thus, lack of associations with these polygenic scores would suggest that GLP1-RA and BS-related weight loss is independent of one’s genetic predisposition to BMI or type 2 diabetes, as explained by common genetic variants. Moreover, because the biological underpinning of weight change is partially distinct from weight at baseline, we also consider the few replicated genetic signals for BMI variability and BMI trajectories.

For bariatric surgery we observed that genetically higher BMI was associated with weight gain, in the opposite direction for the phenotypic association with initial body weight. This might suggest that individuals with a higher genetic predisposition for elevated BMI might regain weight, after the operation, faster than those with lower genetic predisposition. To maintain weight loss, they may need additional lifestyle modifications, behavioral health therapy or adjuvant pharmacologic therapies as a GLP1-RA treatment.^70,71^

On the contrary, none of the genetic exposures were associated with weight loss among GLP1-RA users, after multiple-testing correction. These results were not attributable to high heterogeneity across studies or across major continental ancestry groups. Whether we directly tested the impact of genetic exposure on percentage body weight change or used post-treatment weight as outcome and tested the interaction between baseline weight and the genetic exposure, results did not significantly change.

In addition, we conducted extensive sensitivity analyses to further examine the effect of BMI and T2D PGS on weight loss after BS and GLP1-RA treatment. These analyses included alternative definitions of weight outcomes, a shorter follow-up period and stricter inclusion criteria for BS, adjustments for T2D status, subgroup analysis for GLP1-RA medication, and testing different statistical models. Across all these analyses, we did not observe a significant effect of BMI or T2D PGS on weight reduction following GLP1-RA treatment, in concordance with our primary analysis. Regarding the impact of BMI PGS on weight loss due BS, we observed a reduction of effect in some of the sensitivity analyses, in particular when excluding baseline weight adjustment from the model. In this case, the directionality of the effect remained consistent, however the association was no longer statistically significant. Overall, our results showed that the considered genetic influence on weight loss in response to GLP1-RA treatment and BS may be too small to impact clinical decision-making or patient outcomes. This statement should be revisited either by using more powerful polygenic scores or by considering individuals undergoing GLP1-RA treatment only for weight loss.

Another aim of the study was to compare the effects of GLP1-RA treatment and BS on weight loss across major continental ancestry groups. Ancestry can provide more useful information about population health than racial and ethnic categories because it is not directly influenced by geographic, cultural, and sociopolitical forces.^72^ Nonetheless, ancestry is also partially correlated with self-reported race and ethnicity and socio-economic characteristics do differ across racial and ethnic groups. Overall, we did not find large differences in GLP1-RA and BS associated weight loss between major continental ancestry groups. We do observe that GLP1-RA users of African and admixed American ancestry had statistically significantly lower change in body weight compared to Europeans, after accounting for differences in sex, age, weight at baseline and medication type. The effects were consistent in BioMe and UCLA-ATLAS biobanks for admixed American ancestry, and additionally in AoU and MGBB for African ancestry. This might reflect different socio-economic characteristics between individuals of admixed American and European ancestry resulting, for example, in differences in access to treatment.

The main strength of our study is its size and diversity across countries, healthcare systems and ancestries. Our study presents various limitations. First, we cannot be sure that all GLP1-RA users fulfilled their prescription. This is a potential issue only in studies with prescription information but not in studies with purchase information. Nonetheless weight loss was comparable across both study types. Moreover, we ensured that observed changes in body weight are due to the treatment by considering only measurements within one month from the last purchase or prescription. Second, we could not accurately estimate treatment adherence, but overcome this limitation by considering treatments of maximum 12 months, a period for which adherence has been observed to be around 65%.^65^ Third, we used ancestry labels based on major continental ancestry groups, while ancestry can be better characterized as a continuous quantity.^72^ This discretization reflects pragmatic considerations to enable comparable analyses across the different studies. Fourth, the absence of a control group in the GLP1-RA analysis may limit distinguishing intentional from unintentional weight loss. However, large-scale real-world data and consistent sensitivity analysis support the robustness of our results. Fifth, although T2D medications like metformin can influence weight loss, adjustments for T2D in our sensitivity analyses likely accounted for first-line medications. Finally, we did not assess the impact of GLP1-RA dosing, as dose adjustments vary over time, higher doses do not consistently correlate with greater weight loss and inconsistent reporting across studies could introduce biases.

In conclusion, our study suggests that GLP1-RA treatments work equally well in individuals carrying common non-synonymous mutations in the GLP1R gene and in individuals at high genetic risk for BMI and type 2 diabetes. It also suggests that BS is less effective among individuals with higher polygenic score for BMI, but the effect is not large enough to be clinically relevant. In fact, for each additional SD in the polygenic score for BMI, individuals undergoing BS have on average 0.7% lower weight loss from their initial body weight. Finally, these two major weight loss interventions achieved sustained weight reduction regardless of an individual’s genetic ancestry. However, socio-economic characteristics correlating with ancestry should be further studied to better understand some of the remaining differences observed in this study.

## Methods

### Study population

In the current study, we included samples from 10 960 individuals from the following 9 biobanks: Helsinki University Hospital (HUS, Finland; N = 633), Estonian Biobank (ESTBB, Estonia; N = 464), UK Biobank (UKBB, United Kingdom; N = 810), All of Us (AoU, USA; N = 559), BioMe (BioMe, USA; N = 2,170), Mass General Brigham Biobank (MGBB, USA; N = 2,141), Atlas Biobank (UCLA-ATLAS, USA; N = 1,445), Qatar Biobank (QBB, Qatar; N = 2,383), Bialystok Bariatric Surgery Study (BBSS, Poland, N = 355). The biobank studies include samples from (hospital) biobanks, prospective epidemiological and disease-based cohorts. Follow-up covers a total of 20 years with the earliest study starting follow-up in 2004 (AoU) and the latest study ending follow-up in 2024 (BioMe). In the **Supplementary Material** we provide a detailed description of the population selected and ancestry definitions in each study.

### Inclusion and exclusion criteria for GLP1-RA analysis

For our GLP1-RA analysis we implemented the following inclusion and exclusion criteria. We required an initiation of a GLP1-RA treatment (ATC codes: A10BJ*, A10BX04, A10BX10, A10BX13, A10BX14) defined as the date of first medication purchase or medication prescription of GLP1-RA. Only first-time (GLP1-RA naïve) users within the datasets were included. Individuals were required to be on a GLP1-RA treatment for at least twelve months. This was defined through prescription length, regular drug prescription refills or drug purchases, depending on the dataset and healthcare setting. Individuals aged under 18 were excluded from the analysis. Furthermore, individuals were required to have at least one body weight measurement maximum one year prior to treatment initiation or within 14 days after initiation, if no measure before initiation was available. If we observed multiple weight measurements within this time frame, only the closest one to initiation was considered. This measurement defined our baseline weight variable. At least one body weight measurement between 26 weeks and 52 weeks was necessary for each individual to be included in the analysis. Individuals were excluded if they underwent BS prior to or during the first year of GLP1-RA treatment.

### Inclusion and exclusion criteria for BS analysis

For our BS analysis we implemented the following inclusion and exclusion criteria. Individuals were included if they underwent any type of BS. The respected code definitions for BS varied across countries and healthcare systems and can be found in **Supplementary Table 20**. The proportions of all procedures across cohorts can be found in **Supplementary Table 21**. Individuals aged under 18 at baseline were excluded from the analysis. Analogously to the GLP1-RA analysis, individuals were required to have at least one body weight measurement maximum one year prior to surgery or within 14 days after it, if no measure before baseline was available. If we observed multiple weight measurements within this time frame, only the closest one to initiation was considered. This measurement defined our baseline weight variable. Additionally, at least one body weight measurement between 26 weeks and 208 weeks was necessary for each individual to be included in the analysis.

### Outcome definitions

In our primary model, for both GLP1-RA and BS we defined the outcome to be the percentage change in body weight from baseline. The baseline body weight measured closest to *T_0_* and between -52 weeks and +2 weeks from *T_0_* was defined as *W_0_*.

The second body weight (*mW_1_*) was defined as the median of body weight measurements between 26 weeks and 52 weeks from *T_0_* for the GLP1-RA analysis and between 180 days and 1460 days for the BS analysis. This was to enhance robustness against outlier measurements occurring within these intervals.

Therefore, the percentage change in body weight from baseline was calculated using 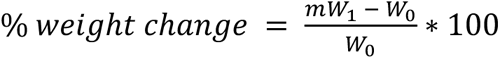.

In our secondary model, we used *mW_1_* as an outcome.

### Genetic exposures

We identified 15 genetic variants (single nucleotide polymorphism, SNP) and two PGS of interest for our analysis. Among the included SNPs 3 are functionally characterized *GLP1R* variants (*rs10305492*, *rs146868158*, *rs6923761*) associated with random glucose levels that have shown decreased/increased response to different endogenous and exogenous GLP1-RA.^57^ Seven SNPs are *GLP1R* missense variants (*rs1042044*, *rs10305420*, *rs3765467*, *rs10305421*, *rs2295006*, *rs10305510*, *rs201672448*) with a frequency over 1% in at least one ancestry group in gnomAD (version 4.1).^56^ One SNP is the only robustly replicated lead genome-wide significant variant (*rs429358*, *ApoE*) in a GWAS of BMI change over time.^54,74^ Two SNPs are *PCSK1* missense variants (*rs6232*, *rs6235*) that have been found associated with BMI variations.^58^ And two SNPs (*rs728996*, *rs17702901*) were associated with an effect on excess body mass index loss amongst BS patients.^49^ A list of all genetic exposures and their frequencies across all major continental ancestry groups can be found in **Supplementary Table 4**. The included PGS are for body mass index (BMI) and type 2 diabetes (T2D).^75^ The PGS weights were taken from Weissbrod et al. 2022^75^ and obtained from UK Biobank. The scores were computed in each study using PLINK 2.0.^76^ Because the PGS scores were derived from UK Biobank, we couldn’t use the same score in UK Biobank, instead we used Thompson et al. 2022.^77^

### Statistical model

We employed a linear regression model to investigate the effect of the chosen genetic exposures on weight change after initiation of GLP1-RA treatment and BS, respectively. In the primary model we adjusted for the baseline weight, sex, age at initiation of treatment or surgery in years, the first 20 genetic principal components and study-specific covariates, such as the genotyping batch. In the GLP1-RA analysis we additionally adjusted for the medication type within the drug class.

We defined % *weight change* as the outcome variable in the primary model.

#### Primary model

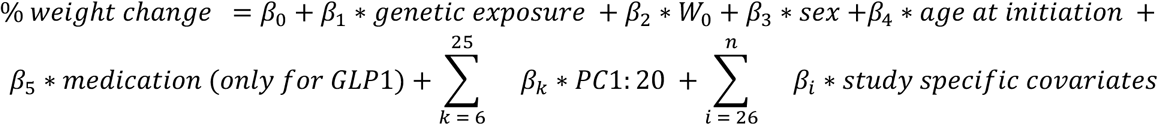

### Meta-analyses

We conducted ancestry-specific and multi-ancestry fixed-effect meta-analyses for each of the genetic exposures. Effect sizes were combined using fixed-effect inverse-variance-weighting (IVW), as implemented in the R package “meta”.^78^ For each exposure we reported the unadjusted *P* values, but considered as threshold for statistical significance *P* < 0.004 for GLP1-RA (i.e. P < 0.05/13, Bonferroni correction for 13 exposures tested) and *P* < 0.01 for BS (i.e. P < 0.05/5, Bonferroni correction for 5 exposures tested). We tested for heterogeneity in effect sizes between ancestries using Cochran’s Q-test^79^ and by inspecting the I^2^ statistics^80^, which is defined as:

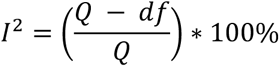

where *df* is equal to *number of studies* − 1.

### Power calculation

To calculate the statistical power for the PGS effect sizes, we used a t-test for linear regression coefficient as implemented in the R package pwrs^81^ assuming a standard deviation for the outcome variable SDy = 7.8 (weighted combine SD across studies), a sample size of n = 6 750, a total number of predictors k = 25 and an adjusted r2 = 0.05, to calculate power at P < 0.05 significance level.

For the coefficients of association with single variants, we estimated statistical power via the non-centrality parameter (NCP) of the chi-square distribution. We defined NCP = 2 f (1 – f) n β^2 where f is MAF, n is the effective sample size, and β is the expected effect size.^82^ We set a beta of 0.3 (% weight change from baseline) as the expected genetic effect on GLP1 response, based on findings from a recent large-scale study on genetic influences on drug response.^64^

## Code Availability

The study utilized previously published analysis tools as described in the **Methods** section. Additional code used for these analyses is available at https://github.com/dsgelab/glp1-bs-genetics/

## Data Availability

The Helsinki biobank can provide access for research projects within the scope regulated by the Finnish Biobank Act, which is research utilizing the biobank samples or data for the purposes of promoting health, understanding the mechanisms of disease or developing products and treatment practices used in health and medical care.

De-identified data of the MGBB that supports this study is available from the MGB Biobankportal at https://www.massgeneralbrigham.org/en/research-and-innovation/participate-in-research/biobank/for-researchers. Restrictions apply to the availability of these data, which are available to MGB-affiliated researchers via a formal application.

UK Biobank data are available through a procedure described at http://www.ukbiobank.ac.uk/using-the-resource/.

Registered researchers whose institutions have Data Use and Registration Agreements in place with All of Us that include the Controlled Tier can access genomic data.

Clinical and genotype data from UCLA ATLAS Community Health Initiative patients, de-identified for research purposes, are accessible to UCLA-approved researchers through the Discovery Data Repository (DDR).

BioMe data is available through a process described at https://icahn.mssm.edu/research/ipm/programs/biome-biobank/researcher-faqs.

Pseudonymised data and/or biological samples can be accessed for research and development purposes in accordance with the Estonian Human Genome Research Act. To access data, the research proposal must be approved by the Scientific Advisory Committee of the Estonian Biobank as well as by the Estonian Committee on Bioethics and Human Research. For more details on data access and relevant documents, please see https://genomics.ut.ee/en/content/estonian-biobank#dataaccess.

For access to the Bialystok Bariatric Surgery Study please refer to the Medical University of Bialystok.

The data and biosamples collected or generated by QBB will be made available to researchers employed within or otherwise contractually bound to public and private institutions that conduct scientific research and that meet the requirements detailed in the Qatar Biobank Research Access policy.

The PGS weights used in this study are available in Weissbrod et al. 2022 and Thompson et al. 2022.^75^

## Data Availability

All data utilized in this study can be accessed by applying to each of the participating studies individually. Additional details are provided in the manuscript under the Data Availability section.

## Acknowledgements

We gratefully acknowledge the resources provided by the Institute for Precision Health and participating patients from the UCLA ATLAS Community Health Initiative (https://www.uclahealth.org/international-services/consulting-services/clinical-research-consultation/ucla-atlas-community-health-initiative). The UCLA ATLAS Community Health Initiative in collaboration with UCLA ATLAS Precision Health Biobank is a program of the Institute for Precision Health, which directs and supports the biobanking and genotyping of biospecimen samples from participating patients from UCLA in collaboration with the David Geffen School of Medicine, UCLA Clinical and Translational Science Institute and UCLA Health. The ATLAS Community Health Initiative is supported by UCLA Health, the David Geffen School of Medicine and a grant from the UCLA Clinical and Translational Science Institute (UL1TR001881). This work was financially supported in part by National Institutes of Health awards U01HG011715, R01HG009120 and R01MH115676. The content is solely the responsibility of the authors and does not necessarily represent the official views of the National Institutes of Health.

We would furthermore like to thank all participants of HUS (https://www.helsinginbiopankki.fi/en/Helsinki-Biobank; https://www.hus.fi/en), ESTBB (https://genomics.ut.ee/en/content/estonian-biobank), BBSS (https://classic.clinicaltrials.gov/ct2/show/NCT04634591), QBB (https://www.qphi.org.qa/), BioMe (https://icahn.mssm.edu/research/ipm/programs/biome-biobank), UKBB (https://www.ukbiobank.ac.uk/), MGBB (https://www.massgeneralbrigham.org/en/research-and-innovation/participate-in-research/biobank) and AOU (https://allofus.nih.gov/) for their generous contribution. This work was further supported in part by funding from the Eric and Wendy Schmidt Center at the Broad Institute of MIT and Harvard.

U.V. and K.A. have been funded by Estonian Research Council’s personal research funding start-up grant PSG759 and European Research Council’ starting grant under the grant agreement no 101117251 (OBECAUSE). J.H.L. is supported by NIDDK K23 DK131345.

The activities of the EstBB are regulated by the Human Genes Research Act, which was adopted in 2000 specifically for the operations of the EstBB. Individual level data analysis in the EstBB was carried out under ethical approvals of 1.1-12/1409 and 1.1-12/2161 from the Estonian Committee on Bioethics and Human Research (Estonian Ministry of Social Affairs), using data according to release application 6-7/GI/18857 from the Estonian Biobank.

## Notes

### Competing Interest Statement

The authors have declared no competing interest.

### Funding Statement

This study was in part supported by funding from the Eric and Wendy Schmidt Center at the Broad Institute of MIT and Harvard.
This study was in part supported by funding from Estonian Research Council's personal research funding start-up grant PSG759 and European Research Council' starting grant under the grant agreement no 101117251 (OBECAUSE).
This study was in part supported by NIDDK K23 DK131345.
This work was further financially supported in part by National Institutes of Health awards U01HG011715, R01HG009120 and R01MH115676.

### Author Declarations

No IRB needed

### Summary of Updates

Sensitivity analyses section added; All sections revised (mainly discussion); Table 1 revised; author affiliations updated; Supplemental files updated

