## Supplementary Material for "Association between plausible genetic factors and weight loss from GLP1-RA and bariatric surgery: a multi-ancestry study in 10 960 individuals from 9 biobanks"

### Supplementary Figure 1 – Study Design

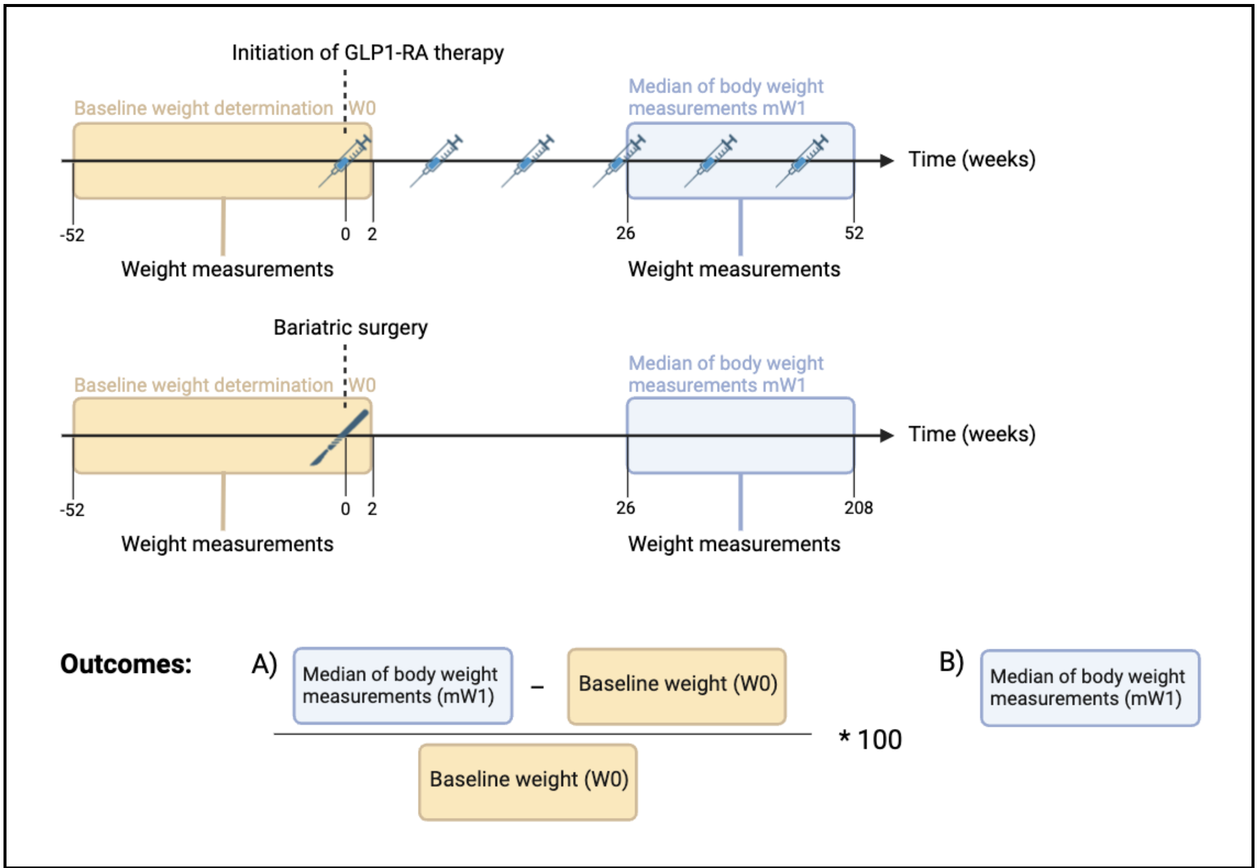


**Supplementary Figure 1 | Study Design**

### Supplementary Figure 2 – Effect sizes of baseline weight on weight loss amongst GLP1-RA users across studies by ancestries

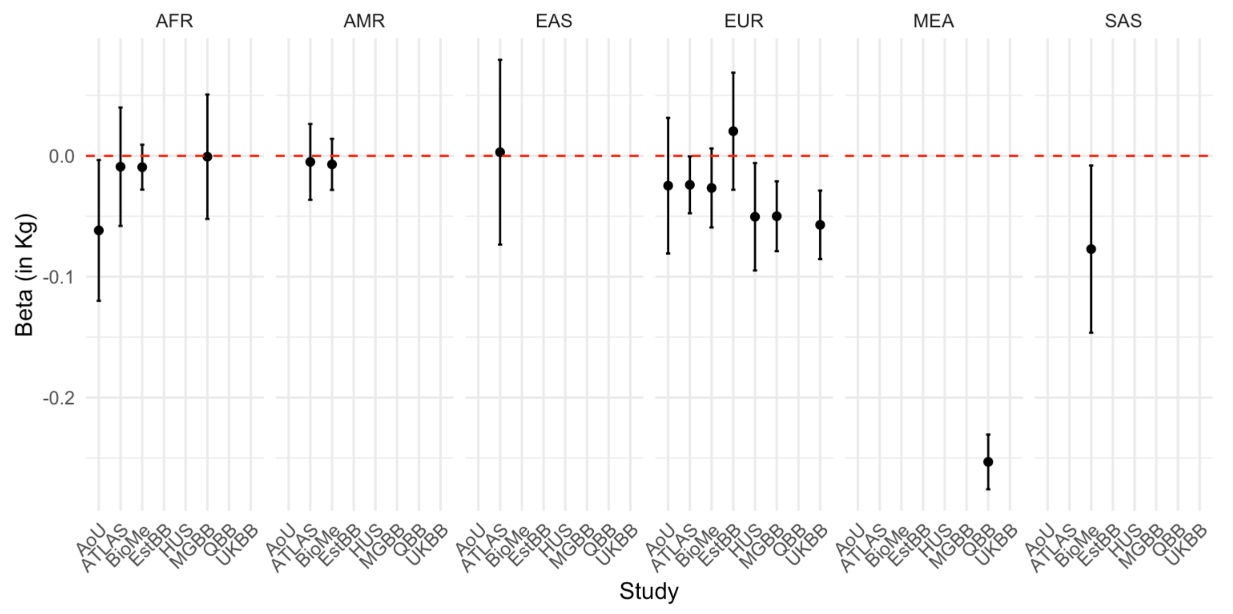


**Supplementary Figure 2 | Effect sizes of baseline weight on weight loss amongst GLP1-RA users across studies by ancestries.** Points represent beta coefficients in Kg and error bars represent the 95% confidence intervals. Negative beta coefficients indicate that higher baseline weight is associated with greater weight loss, while positive beta coefficients indicate the opposite.
AoU: All of Us, ATLAS: UCLA-Atlas Biobank, BioMe: BioMe Biobank, ESTBB: Estonian Biobank, HUS: Helsinki University Hospital, MGBB: Mass General Brigham Biobank, QBB: Qatar Biobank, UKBB: UK Biobank.

### Supplementary Figure 3 – Effect sizes of sex on weight loss amongst GLP1-RA users across studies by ancestries

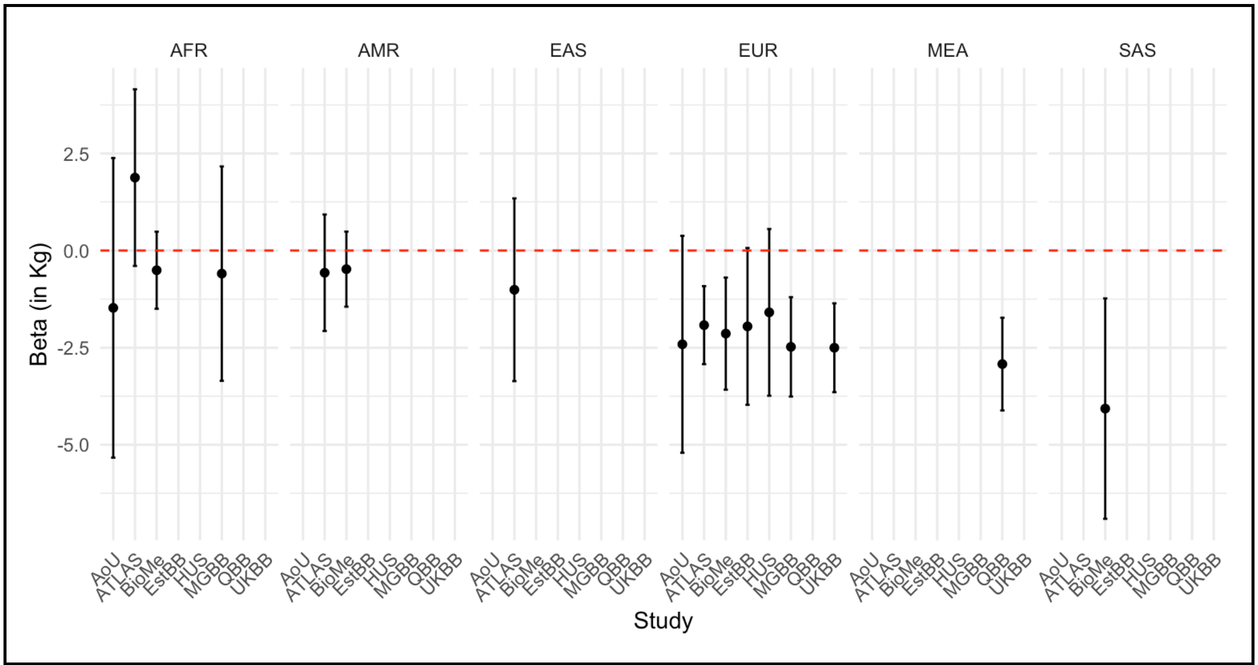


**Supplementary Figure 3 | Effect sizes of sex on weight loss amongst GLP1-RA users across studies by ancestries.** Points represent beta coefficients in Kg and error bars represent the 95% confidence intervals. Negative beta coefficients indicate that female sex is associated with greater weight loss, while positive beta coefficients indicate the opposite.

AoU: All of Us, ATLAS: UCLA-Atlas Biobank, BioMe: BioMe Biobank, ESTBB: Estonian Biobank, HUS: Helsinki University Hospital, MGBB: Mass General Brigham Biobank, QBB: Qatar Biobank, UKBB: UK Biobank.

### Supplementary Figure 4 – Effect sizes of age at treatment initiation on weight loss amongst GLP1-RA users across studies by ancestries

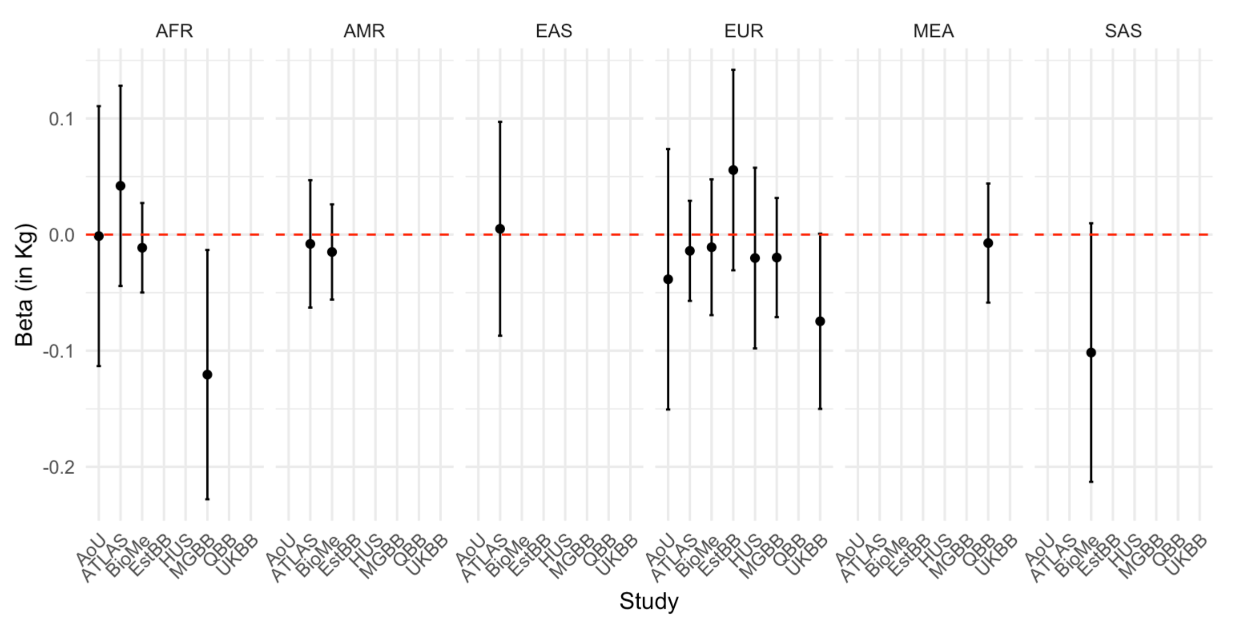


**Supplementary Figure 4 | Effect sizes of age at treatment initiation on weight loss amongst GLP1-RA users across studies by ancestries.** Points represent beta coefficients in Kg and error bars represent the 95% confidence intervals. Negative beta coefficients indicate that baseline age is associated with greater weight loss, while positive beta coefficients indicate the opposite.
AoU: All of Us, ATLAS: UCLA-Atlas Biobank, BioMe: BioMe Biobank, ESTBB: Estonian Biobank, HUS: Helsinki University Hospital, MGBB: Mass General Brigham Biobank, QBB: Qatar Biobank, UKBB: UK Biobank.

### Supplementary Figure 5 – Sensitivity analysis – "utilizing the minimum weight (minW1) instead of the median weight (mW1)": Average percentage change in body weight

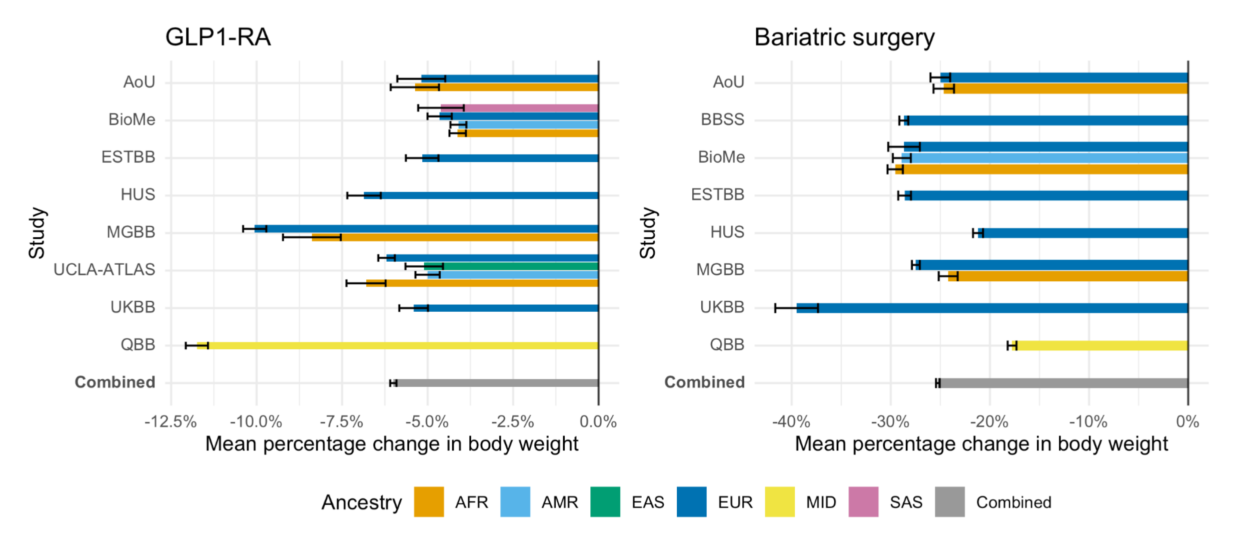


**Supplementary Figure 5 | Sensitivity analysis – utilizing the minimum weight (minW1) instead of the median weight (mW1): Average percentage change in body weight for 6 750 GLP1-RA users and 4 210 bariatric surgery patients.**
Bars represent the mean percentage change in body weight within each study and ancestry, and the overall combined weighted mean change. Error bars represent standard errors.
The average percentage change in body weight is calculated using the minimum postoperative or post-treatment weight, as per the relaxed outcome definition.
The method for this sensitivity analysis is described in the **Supplementary Methods**.
AoU, All of Us; BBSS, Bialystok Bariatric Surgery Study; ESTBB, Estonian Biobank; HUS, Helsinki Biobank; MGBB, Mass General Brigham Biobank; UCLA-ATLAS, Atlas Biobank; UKBB, UK Biobank; QBB, Qatar Biobank; AFR, African; AMR, Admixed American; EAS, East Asian; EUR, European; MID, Middle Eastern; SAS, South Asian

### Supplementary Figure 6 – Sensitivity analysis – "utilizing the minimum weight (minW1) instead of the median weight (mW1)": Effect of T2D and BMI polygenic scores on body weight changes

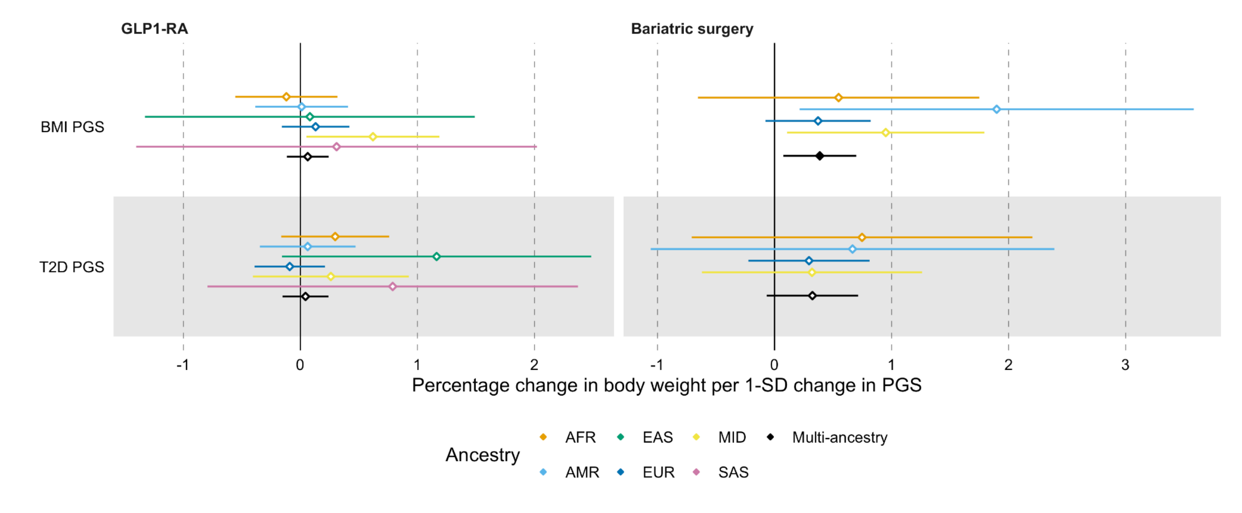


**Supplementary Figure 6 | Sensitivity analysis – utilizing the minimum weight (minW1) instead of the median weight (mW1): Effect of T2D and BMI polygenic scores on body weight changes associated with GLP1-RA treatment and bariatric surgery.**

Ancestry-specific and multi-ancestry meta-analysis effect sizes for association between percentage change in body weight and PGS for BMI and type 2 diabetes. Dots represent the percentage change in body weight per one standard deviation change in PGS, error bars represent the 95% confidence interval. Full dots represent statistical significance at $P < 0.025$.

The average percentage change in body weight is calculated using the minimum postoperative or post-treatment weight, as per the relaxed outcome definition.

The method for this sensitivity analysis is described in the **Supplementary Methods**.

AFR, African; AMR, Admixed American; EAS, East Asian; EUR, European; MID, Middle Eastern; SAS, South Asian

### Supplementary Figure 7 – Sensitivity analysis – "shorter follow-up (12 months) for the BS analysis": Average percentage change in body weight

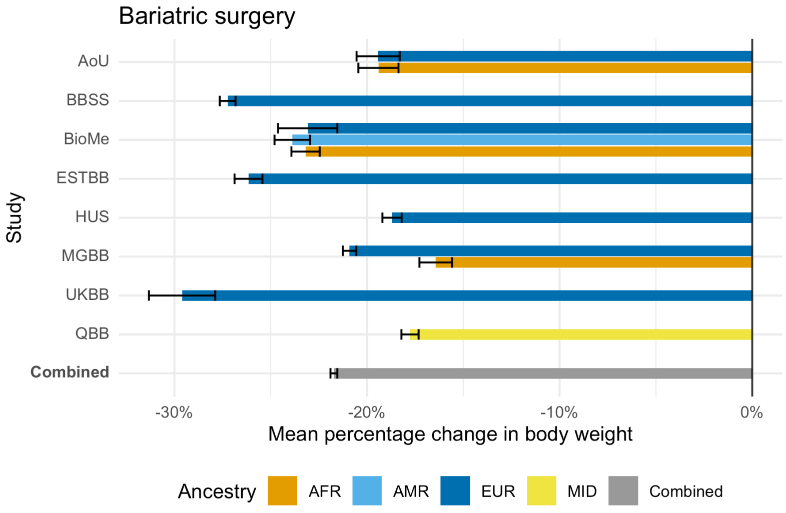


**Supplementary Figure 7 | Sensitivity analysis – shorter follow-up (12 months) for the BS analysis: Average percentage change in body weight for 3 711 bariatric surgery patients.**
Bars represent the mean percentage change in body weight within each study and ancestry, and the overall combined weighted mean change. Error bars represent standard errors.
The method for this sensitivity analysis is described in the **Supplementary Methods**.
AoU, All of Us; BBSS, Bialystok Bariatric Surgery Study; ESTBB, Estonian Biobank; HUS, Helsinki Biobank; MGBB, Mass General Brigham Biobank; UKBB, UK Biobank; QBB, Qatar Biobank; AFR, African; AMR, Admixed American; EAS, East Asian; EUR, European; MID, Middle Eastern; SAS, South Asian

### Supplementary Figure 8 – Sensitivity analysis – “shorter follow-up (12 months) for the BS analysis”: Effect of T2D and BMI polygenic scores on body weight changes

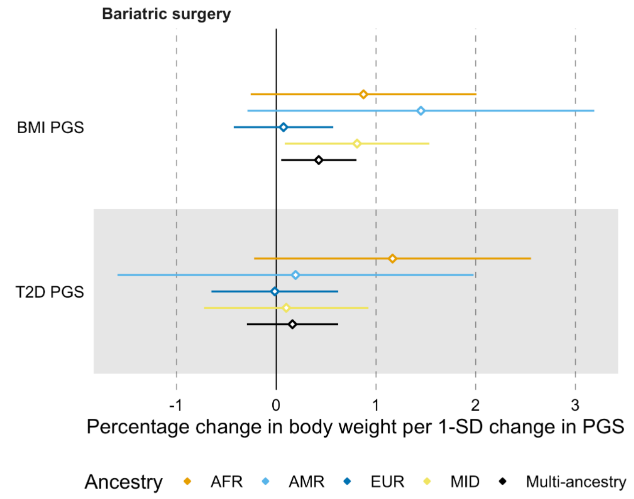


**Supplementary Figure 8 | Sensitivity analysis – shorter follow-up (12 months) for the BS analysis: Effect of T2D and BMI polygenic scores on body weight changes associated with GLP1-RA treatment and bariatric surgery.**
Ancestry-specific and multi-ancestry meta-analysis effect sizes for association between percentage change in body weight and PGS for BMI and type 2 diabetes. Dots represent the percentage change in body weight per one standard deviation change in PGS, error bars represent the 95% confidence interval. Full dots represent statistical significance at $P < 0.025$.
The method for this sensitivity analysis is described in the **Supplementary Methods**.
AFR, African; AMR, Admixed American; EUR, European; MID, Middle Eastern

### Supplementary Figure 9 – Sensitivity analysis – “adjusting for T2D status in the statistical model”: Effect of T2D and BMI polygenic scores on body weight changes

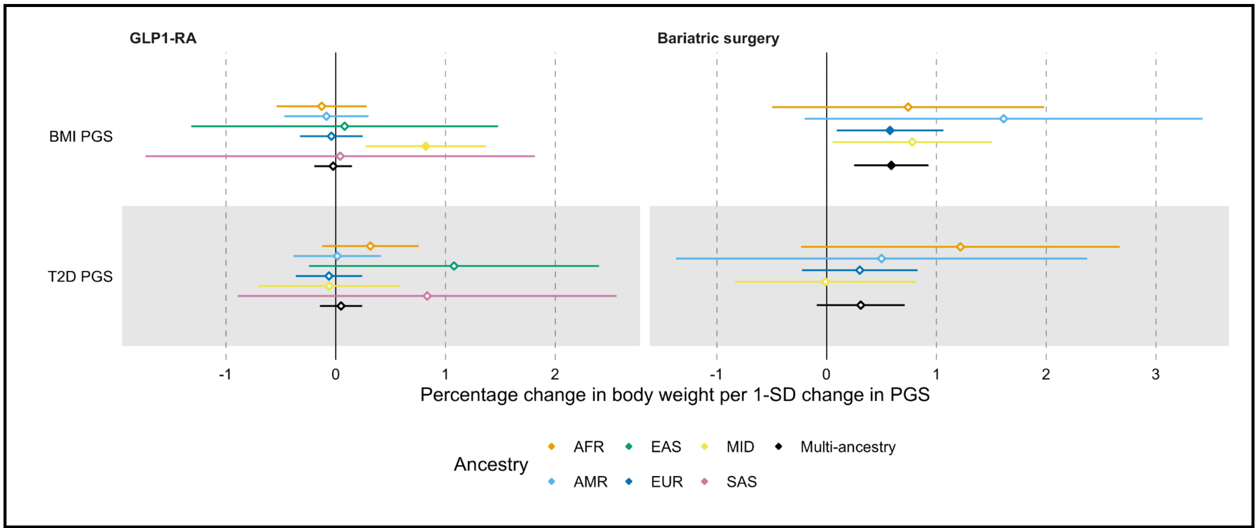


**Supplementary Figure 9 | Sensitivity analysis – adjusting for T2D status in the statistical model: Effect of T2D and BMI polygenic scores on body weight changes associated with GLP1-RA treatment and bariatric surgery.**

Ancestry-specific and multi-ancestry meta-analysis effect sizes for association between percentage change in body weight and PGS for BMI and type 2 diabetes. Dots represent the percentage change in body weight per one standard deviation change in PGS, error bars represent the 95% confidence interval. Full dots represent statistical significance at $P < 0.025$.

The method for this sensitivity analysis is described in the **Supplementary Methods**.

AFR, African; AMR, Admixed American; EAS, East Asian; EUR, European; MID, Middle Eastern; SAS, South Asian

### Supplementary Figure 10 – Sensitivity analysis – “including only liraglutide/semaglutide in GLP1-RA analysis”: Average percentage change in body weight

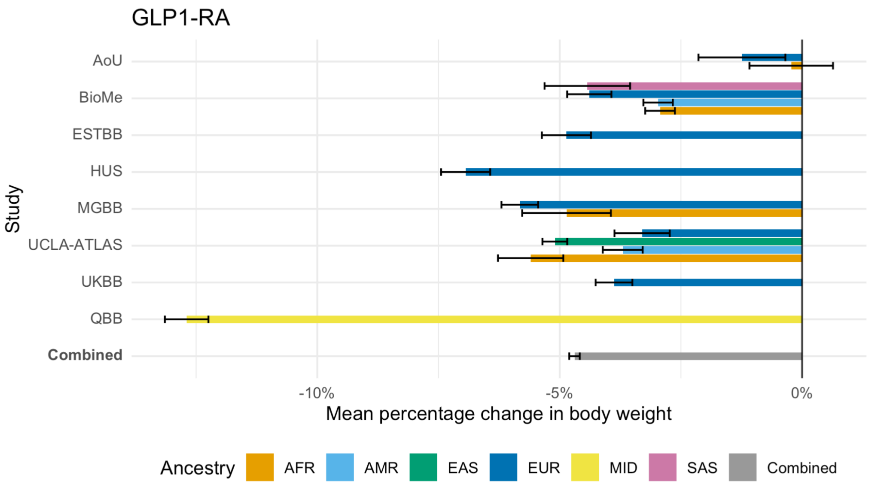


**Supplementary Figure 10 | Sensitivity analysis – including only liraglutide/semaglutide in GLP1-RA analysis: Average percentage change in body weight for 4 431 GLP1-RA users.**
Bars represent the mean percentage change in body weight within each study and ancestry, and the overall combined weighted mean change. Error bars represent standard errors.
The method for this sensitivity analysis is described in the **Supplementary Methods**.
AoU, All of Us; ESTBB, Estonian Biobank; HUS, Helsinki Biobank; MGBB, Mass General Brigham Biobank; UCLA-ATLAS, Atlas Biobank; UKBB, UK Biobank; QBB, Qatar Biobank; AFR, African; AMR, Admixed American; EAS, East Asian; EUR, European; MID, Middle Eastern; SAS, South Asian

### Supplementary Figure 11 – Sensitivity analysis – “including only liraglutide/semaglutide in GLP1-RA analysis”: Effect of T2D and BMI polygenic scores on body weight changes

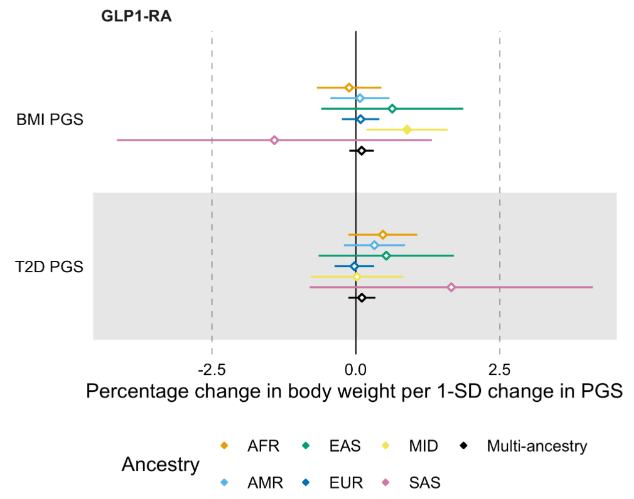


**Supplementary Figure 11 | Sensitivity analysis – including only liraglutide/semaglutide in GLP1-RA analysis: Effect of T2D and BMI polygenic scores on body weight changes associated with GLP1-RA treatment and bariatric surgery.**
Ancestry-specific and multi-ancestry meta-analysis effect sizes for association between percentage change in body weight and PGS for BMI and type 2 diabetes. Dots represent the percentage change in body weight per one standard deviation change in PGS, error bars represent the 95% confidence interval. Full dots represent statistical significance at $P < 0.025$.
The method for this sensitivity analysis is described in the **Supplementary Methods**.
AFR, African; AMR, Admixed American; EAS, East Asian; EUR, European; MID, Middle Eastern; SAS, South Asian

#### Supplementary Figure 12 – Sensitivity analysis – “stricter inclusion criteria for the BS analysis”: Average percentage change in body weight

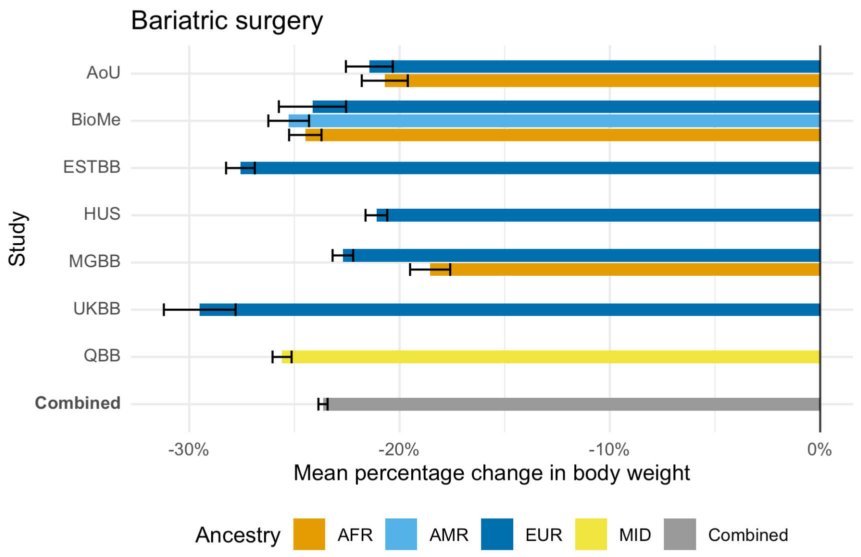


**Supplementary Figure 12 | Sensitivity analysis – stricter inclusion criteria for the BS analysis: Average percentage change in body weight for 3 710 bariatric surgery patients.**
Bars represent the mean percentage change in body weight within each study and ancestry, and the overall combined weighted mean change. Error bars represent standard errors.
Only participants with weight measurements available at least 30 days prior to surgery (from -365 days to -30 days relative to the day of surgery) were included.
The method for this sensitivity analysis is described in the **Supplementary Methods**.
AoU, All of Us; BBSS, Bialystok Bariatric Surgery Study; ESTBB, Estonian Biobank; HUS, Helsinki Biobank; MGBB, Mass General Brigham Biobank; UKBB, UK Biobank; QBB, Qatar Biobank; AFR, African; AMR, Admixed American; EAS, East Asian; EUR, European; MID, Middle Eastern; SAS, South Asian

### Supplementary Figure 13 – Sensitivity analysis – “stricter inclusion criteria for the BS analysis”: Effect of T2D and BMI polygenic scores on body weight changes

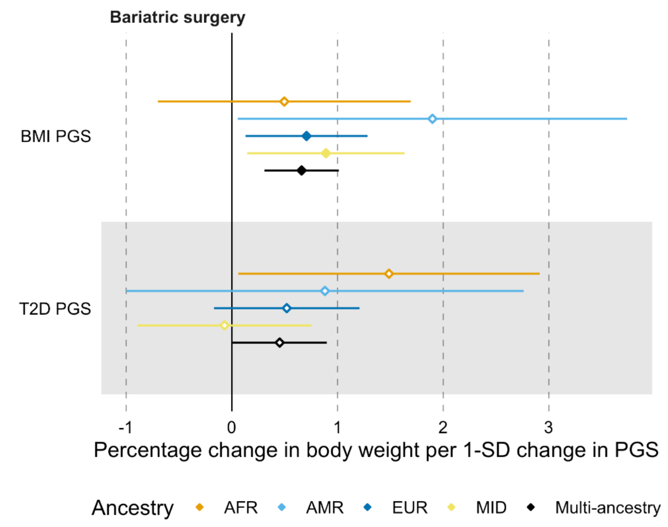


**Supplementary Figure 13 | Sensitivity analysis – stricter inclusion criteria for the BS analysis: Effect of T2D and BMI polygenic scores on body weight changes associated with GLP1-RA treatment and bariatric surgery.**

Ancestry-specific and multi-ancestry meta-analysis effect sizes for association between percentage change in body weight and PGS for BMI and type 2 diabetes. Dots represent the percentage change in body weight per one standard deviation change in PGS, error bars represent the 95% confidence interval. Full dots represent statistical significance at $P < 0.025$.

Only participants with weight measurements available at least 30 days prior to surgery (from -365 days to -30 days relative to the day of surgery) were included.

The method for this sensitivity analysis is described in the **Supplementary Methods**.

AFR, African; AMR, Admixed American; EUR, European; MID, Middle Eastern

### Supplementary Figure 14 – Sensitivity analysis – “model without adjustment for baseline weight (W0)”: Effect of T2D and BMI polygenic scores on body weight changes

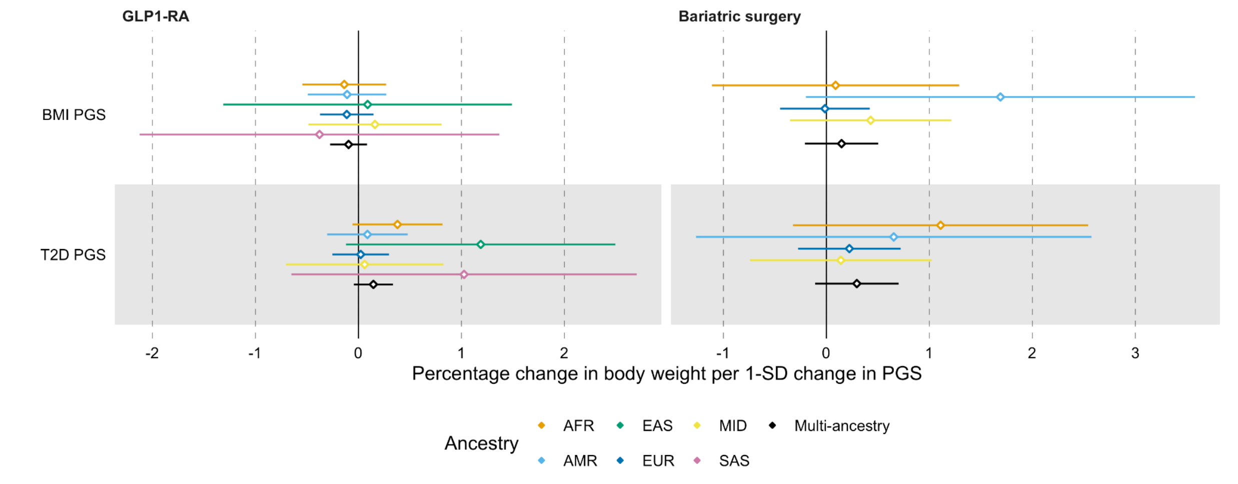


**Supplementary Figure 14 | Sensitivity analysis – model without baseline weight (W0): Effect of T2D and BMI polygenic scores on body weight changes associated with GLP1-RA treatment and bariatric surgery.**

Ancestry-specific and multi-ancestry meta-analysis effect sizes for association between percentage change in body weight and PGS for BMI and type 2 diabetes. Dots represent the percentage change in body weight per one standard deviation change in PGS, error bars represent the 95% confidence interval. Full dots represent statistical significance at $P < 0.025$.

The method for this sensitivity analysis is described in the **Supplementary Methods**.

AFR, African; AMR, Admixed American; EAS, East Asian; EUR, European; MID, Middle Eastern; SAS, South Asian

### Supplementary Material – Sensitivity analyses

All sensitivity analyses focused on the effects of the two polygenic scores (BMI and T2D PGS) rather than single genetic variants. Although the primary analyses had sufficient statistical power to detect effects from individual variants, no significant associations were observed. Consequently, we prioritized the polygenic scores in the sensitivity analyses, as they capture aggregated genetic effects and provide a more comprehensive evaluation of potential genetic contributions.

#### Supplementary Methods

##### Sensitivity analysis: Utilizing the minimum weight (minW1) instead of the median weight (mW1)

To explore the impact of different approaches to defining postoperative or post-drug therapy weight measurements, we conducted a sensitivity analysis using the minimum observed weight (minW1) instead of the median weight (mW1) as the primary outcome. This analysis aimed to evaluate whether our choice of using the median weight, which provides a more conservative and robust estimate by minimizing the influence of outliers, influenced the observed results.
The modified outcome variable was defined as:

$$\%\,weight\ change\ = \frac{minW_1\ -\ W_0}{W_0} * 100$$

This sensitivity analysis was applied to both the GLP1-RA and BS cohorts.

The results of this analysis are reported in **Supplementary Tables 9-10, Supplementary Figures 5-6** and further discussed in the **Supplementary Results**.

##### Sensitivity analysis: Shorter follow-up (12 months) for the BS analysis

To assess whether the observed effects in the bariatric surgery (BS) analysis were influenced by the longer follow-up periods typically associated with this intervention, we conducted a sensitivity analysis limiting the follow-up period to 12 months post-surgery. This shorter follow-up period aligns with the timeframe used in the GLP1-RA analysis, allowing for a more direct comparison of weight loss dynamics between the two treatment groups.

For this analysis, only weight measurements within the first 12 months post-surgery were considered. This sensitivity analysis included a total of 3,711 individuals, categorized by genetic ancestry as follows: AFR - 461; AMR - 124; EUR - 1979; MID - 1147.

The results of this analysis are reported in **Supplementary Table 11, Supplementary Figures 7-8** and further discussed in the **Supplementary Results**.

#### Sensitivity analysis: Adjusting for T2D status in the statistical model

To evaluate whether a diagnosis of type 2 diabetes (T2D) independently affects weight loss outcomes and to explore the robustness of our findings when accounting for T2D, we conducted a sensitivity analysis. In this analysis, T2D status was included as an additional covariate in the statistical model alongside the covariates used in the primary analysis. This adjustment allowed us to assess the potential contribution of T2D to weight loss variability and ensure that our findings were not biased by the exclusion of this factor.

The modified statistical model was defined as follows:

$$\%\,weight\ change\ = \beta_0 + \beta_1 * genetic\ exposure\ + \beta_2 * W_0 + \beta_3 * sex\ + \beta_4 * age\ at\ initiation\ +$$

$$\beta_5 * medication\ (only\ for\ GLP1) + \beta_6 * T2D\ status + \sum_{k=7}^{26} \beta_k * PC1{:}20$$

$$+ \sum_{i=27}^{n} \beta_i * study\ specific\ covariates$$

This sensitivity analysis was applied to both the GLP1-RA and BS cohorts.

The results of this analysis are reported in **Supplementary Table 12-13, Supplementary Figure 9** and further discussed in the **Supplementary Results**.

#### Sensitivity analysis: Including only liraglutide/semaglutide in GLP1-RA analysis

To explore whether the genetic and weight loss associations differ when focusing solely on the GLP1-RAs approved for obesity treatment, we conducted a sensitivity analysis including only patients treated with liraglutide or semaglutide. These medications have demonstrated greater efficacy for weight loss compared to other GLP1-RAs and are next to a T2D indication approved for obesity treatment.

Patients in this analysis may have been prescribed liraglutide or semaglutide for either T2D or obesity, as both indications are common. We did not restrict the analysis to only patients with obesity to ensure sufficient sample size and statistical power, as including patients with T2D broadened the cohort and extended the treatment time horizon.

This sensitivity analysis included a total of 4,431 individuals, categorized by genetic ancestry as follows: AFR - 635; AMR - 505; EAS - 769; EUR - 1670; MID - 701; SAS - 51.

The results of this analysis are reported in **Supplementary Table 14, Supplementary Figures 10-11** and further discussed in the **Supplementary Results**.

#### Sensitivity analysis: Stricter inclusion criteria for the BS analysis

To address the potential influence of preoperative low-calorie meal replacement diets on weight measurements, we conducted a sensitivity analysis with stricter inclusion criteria for the BS cohort. This analysis included only participants with weight measurements taken at least 30 days prior to surgery, within the time window of -365 days to -30 days relative to the day of surgery.

The Polish cohort (BBSS) was excluded from this analysis, as weight measurements for this cohort were only available on the day of surgery. This adjustment aimed to minimize the potential confounding effects of pre-surgical dietary interventions on baseline weight measurements and subsequent weight change calculations.

This sensitivity analysis included a total of 3,710 individuals categorized by genetic ancestry as follows: AFR - 510; AMR - 140; EUR - 1913; MID - 1147.

The results of this analysis are reported in **Supplementary Table 15, Supplementary Figures 12-13** and further discussed in the **Supplementary Results**.

#### Sensitivity analysis: Model using post-treatment weight as outcome and including interaction between baseline weight and each genetic exposure

As previous works have argued that using percentage changes in weight loss pre vs post therapy may be statistically inappropriate and inefficient [PMID: 11459516, 24022766], we considered a model using post-treatment weight as outcome, including the same predictors as in the main model and an additional term modeling the interaction between baseline weight and each genetic exposure.

The modified statistical model was defined as follows:

$$mW_1 = \beta_0 + \beta_1 * genetic\ exposure + \beta_2 * W_0 + \beta_3 * genetic\ exposure * W_0 + \beta_4 * sex + \beta_5 * age\ at\ initiation + \beta_6 * medication\ (only\ for\ GLP1) + \sum_{k=7}^{26} \beta_k * PC1:20 + \sum_{i=27}^{n} \beta_i * study\ specific\ covariates$$

The results of this analysis are reported in **Supplementary Tables 16-17** and further discussed in the **Supplementary Results**.

#### Sensitivity analysis: Model without adjustment for baseline weight (W0)

It has been proposed that adjusting for a baseline variable, such as baseline weight (W0), which is influenced by the same genetic factors as the outcome variable, may introduce bias and lead to spurious associations or altering the directionality of observed effects [PMID: 38633781, 15987729, 25401453]. To evaluate whether the inclusion of baseline weight in our model might have affected the observed genetic associations, we conducted a sensitivity analysis in which W0 was excluded from the statistical model.

The modified statistical model was defined as follows:

$$\% \, weight\ change = \beta_0 + \beta_1 * genetic\ exposure + \beta_2 * sex + \beta_3 * age\ at\ initiation + \beta_4 * medication\ (only\ for\ GLP1) + \sum_{k=5}^{24} \beta_k * PC1{:}20 + \sum_{i=25}^{n} \beta_i * study\ specific\ covariates$$

This sensitivity analysis was applied to both the GLP1-RA and BS cohorts.

The results of this analysis are reported in **Supplementary Table 18-19, Supplementary Figure 14** and further discussed in the **Supplementary Results**.

### Supplementary Results

#### Sensitivity analysis: Utilizing the minimum weight (minW1) instead of the median weight (mW1)

Using the minimum weight (minW1) as the primary outcome resulted in a higher observed weight loss compared to the median weight (mW1), consistent with expectations.

For the GLP1-RA cohort, the average body weight change across studies was -6.00% (ranging from -4.10% to -11.75%) (**Supplementary Figure 5**). As in the primary analysis, no significant genetic effects explaining the heterogeneity in weight loss outcomes were observed in this cohort (**Supplementary Table 9** and **Supplementary Figure 6**).

For the BS cohort, the average body weight change was -25.25% (ranging from -17.77% to -39.50%) (**Supplementary Figure 5**). The previously observed effect of the BMI PGS on weight change was confirmed, albeit with a lower effect estimate ($\beta_{BMI\ PGS}$ = 0.39 % weight change compared to baseline for 1 SD change in the polygenic score, $P = 1.51 \times 10^{-2}$) (**Supplementary Table 10** and **Supplementary Figure 6**).

These findings demonstrate that the choice of postoperative or post-treatment weight definition does not alter the conclusions of our primary analysis. The genetic effect of the BMI PGS remains evident in the context of BS but is not observed in GLP1-RA users, consistent with our main findings.

This sensitivity analysis underscores the robustness of our results and strengthens our conclusion that genetic effects were detected only in the BS cohort for explaining weight loss variability.

#### Sensitivity analysis: Shorter follow-up (12 months) for the BS analysis

Limiting the follow-up period for BS to 12 months yielded an average body weight change of -21.72% (ranging from -16.43% to -29.60%) , which is consistent with our main findings of -21.17% average body weight change. This suggests that the majority of weight reduction occurs within the first year post-surgery (**Supplementary Figure 7**).

The effect of the BMI PGS on weight change during this shorter follow-up period was estimated at $\beta_{BMI\ PGS}$ = 0.43% weight change per 1 SD change in the polygenic score. While this result was nominally significant ($P = 2.69 \times 10^{-2}$), it did not reach statistical significance after correcting for multiple tests **(Supplementary Table 11** and **Supplementary Figure 8**).

These findings suggest that while the majority of weight loss occurs within the first year, the shorter follow-up does not fully capture the statistically significant genetic effect pulling towards higher weight observed in the primary analysis and other sensitivity analyses. This highlights the importance of the extended follow-up period used in the primary analysis to provide a more accurate representation of long-term outcomes and genetic contributions.

#### Sensitivity analysis: Adjusting for T2D status in the statistical model

This sensitivity analysis confirmed the previously observed effect of the BMI PGS on weight change following BS, with a comparable effect estimate to the primary analysis ($\beta_{BMI\ PGS}$ = 0.59% weight change compared to baseline per 1 SD change in the polygenic score, $P = 6.60 \times 10^{-4}$) (**Supplementary Table 13** and **Supplementary Figure 9**).

Among GLP1-RA users, no genetic effect explaining the heterogeneity in weight loss outcomes was observed, consistent with the findings from the primary analysis (**Supplementary Table 12** and **Supplementary Figure 9**).

The analysis further revealed a significant effect of T2D on weight change among GLP1-RA users ($\beta_{T2D}$ = 0.54% weight change compared to baseline, $P = 1.90 \times 10^{-2}$), indicating that individuals with T2D face greater difficulty in losing weight compared to those without T2D, aligning with existing literature (**Supplementary Table 12**). On the other hand, no significant effect of T2D was observed among individuals undergoing BS.

#### Sensitivity analysis: Including only liraglutide/semaglutide in GLP1-RA analysis

The sensitivity analysis revealed an average body weight change of -4.69% (ranging from -1.24% to -12.70%) among patients treated with liraglutide or semaglutide, which was larger than the -3.93% weight loss observed in our main analysis (**Supplementary Figure 10**). This finding aligns with the higher efficacy of these medications for weight loss compared to other GLP1-RAs.

Despite this increased weight loss, no significant genetic effects explaining the variability in weight loss outcomes were observed in this analysis, consistent with the results of the main analysis (**Supplementary Table 14** and **Supplementary Figure 11**).

#### Sensitivity analysis: Stricter inclusion criteria for the BS analysis

Limiting the inclusion criteria to participants with weight measurements available at least 30 days prior to surgery resulted in a slightly larger observed average body weight change of -23.64% (ranging from -18.54% to -29.50%) compared to the primary analysis, which reported an average weight change of -21.17% (ranging from -15.00% to -27.72%) (**Supplementary Figure 12**).

Importantly, this sensitivity analysis confirmed the previously observed effect of the BMI PGS on weight change following bariatric surgery. The effect size ($\beta_{BMI\ PGS}$ = 0.66% weight change compared to baseline per 1 SD change in the polygenic score, $P$ = 2.40 × $10^{-4}$) remained comparable to that of the primary analysis (**Supplementary Table 15** and **Supplementary Figure 13**), which reported an effect size of $\beta_{BMI\ PGS}$ = 0.70%.

This finding supports the assumption that excluding weight measurements taken closer to the surgery date removes the influence of preoperative dietary interventions, which can cause substantial weight loss prior to surgery.

#### Sensitivity analysis: Model using post-treatment weight as outcome and including interaction between baseline weight and each genetic exposure

Using the alternative model, we identified one missense variant in the *GLP1R* genes, *rs3765467*, for which we observed a statistically significant interaction with baseline weight ($\beta_{w0*rs3765467}$ = 0.09 kg weight change from baseline weight, $P$ = 2.7 × $10^{-4}$) on the weight measured post GLP1-RA treatment after multiple testing correction. When considering nominal significance, another missense variant in the *GLP1R* genes, *rs6923761*, had a statistically significant interaction with baseline weight ($\beta_{w0*rs6923761}$ = 0.02 kg weight change from baseline weight, $P$ = 0.01) on the weight measured post GLP1-RA treatment (**Supplementary Table 16**).

As for the BS analysis, we only observed a nominally significant interaction of the BMI PGS with baseline weight ($\beta_{w0*BMI\ PGS}$ = 0.04 kg weight change from baseline weight, $P$ = 8.7 × $10^{-3}$) on the post-operative weight after multiple testing correction (**Supplementary Table 17**).

#### Sensitivity analysis: Model without adjustment for baseline weight ($W_0$)

Removing baseline weight ($W_0$) from the statistical model led to a reduction in the estimated effect size of the BMI PGS in the BS analysis. The effect size was reduced to $\beta_{BMI\ PGS}$ = 0.15% weight change compared to baseline per 1 SD change in the polygenic score ($P$ = 4.20 × $10^{-1}$), compared to $\beta_{BMI\ PGS}$ = 0.70% in the primary model. While the directionality of the effect remained consistent, the association was no longer statistically significant.

For the GLP1-RA analysis, no significant genetic effects were observed, consistent with the findings of the primary analysis.

These results demonstrate that adjusting for baseline weight yielded a statistically significant genetic effect in the primary BS analysis, which was attenuated when baseline weight was removed from the model. Further details can be found in **Supplementary Tables 18-19** and **Supplementary Figure 14**.

### Supplementary Material – Cohort descriptions and ancestry definitions

#### All of Us (AoU)

The All of Us Research Program is a longitudinal cohort study aiming to enroll at least one million individuals across a diverse population in the USA. Over 245,000 individuals have genotype data. The sensitivity for single nucleotide variants was > 98.7% and the precision was > 99.9%. Ancestry categories were based on gnomAD and inferred using PCA data. The patterns of ancestry and admixture were compared to self-identified race and ethnicity, and continuous ancestry inference using genome-wide genotypes resulted in concordant estimates. We restricted participants to an unrelated subset. The participants were further filtered based on the criteria described in the analysis plan (e.g., per ancestry minimum allele count/sample size after filtering on availability of weight measurements and prescription and/or bariatric surgery).

#### Bialystok Bariatric Surgery Study (BBSS)

The Bialystok Bariatric Surgery Study (BBSS) is a prospective cohort study of patients undergoing bariatric surgery at the First Clinical Department of General and Endocrine Surgery at the Medical University of Bialystok. This is the primary receiving center for patients referred for bariatric surgery in the province of Podlaskie Voivodeship and the largest center by number of bariatric surgeries performed in northeastern Poland. This center specializes in several bariatric surgical techniques including Roux-en-Y gastric bypass, gastric banding, and sleeve gastrectomy. For this study, we selected only patients who underwent sleeve gastrectomy since it represents the vast majority (>90%) of all interventions performed at the center and in order to eliminate confounding variation in surgical technique. The BBSS began in 2015 and consisted of a battery of baseline tests established one month prior to the intervention and repeated at one, three, six, and twelve-month follow-up clinical visits. Subsequently, patients are examined every year after the first year. At each visit, all subjects underwent physical examination, body composition analysis, and blood testing, as well as completed diet and physical activity questionnaires. All subjects give their informed consent for inclusion before participating in the study. The study is conducted in accordance with the Declaration of Helsinki, and the protocol was approved by the Ethics Committee of the Medical University of Bialystok (Project identification code: R-I-002/546/2015).

Genetic ancestry in the BBSS cohort was determined using a combination of self-reported race/ethnicity and genotype-based analysis. Self-reported race/ethnicity information was collected during baseline assessments. Additionally, genetic ancestry was further refined through genotype clustering analyses using reference populations from the 1000 Genomes Project.

#### BioMe

The BioMe Biobank, founded in 2007, is an ongoing electronic health record (EHR)-linked biorepository that enrolls participants non-selectively from across the Mountain Sinai Health System (MSHS). Over 60,000 participants have been recruited from >26 outpatients sites located in Manhattan and Queens, and recruitment is ongoing. We restricted participants to an unrelated subset (based on 2nd degree or greater KING-derived kinship coefficients) who had been genotyped with either the Global Screening Array (GSA) or Global Diversity Array (GDA). We then filtered this population to those adhering to the criteria described in the analysis plan (e.g., per-ancestry minimum allele count/sample size after filtering on availability of weight measurements and prescription and/or bariatric surgery).

Genetically determined ancestry group based on ADMIXTURE with K=10, with grouping based on which 1000 Genomes phase 3 reference samples (with superpopulation labels as reference) showed the highest proportion for a given class. The choice for K=10 was based on a combination of minimizing cross-validation errors for admixture analysis, as well as minimizing the Frobenius distance of each ADMIXTURE matrix K with the matrix outputted by Genetic Relationship and Fingerprinting (GrafPop).

#### Estonian Biobank (ESTBB)

ESTBB is a population-based biobank that involves over 212,000 adult participants (about 20% of the Estonian adult population). The Estonian Biobank Project was initiated in 1999; data collection began in 2002. Initially, participants were recruited by general practitioners across Estonia from among individuals visiting general practitioners' offices and hospitals. Consenting adults donated blood samples for DNA extraction, underwent anthropometric, blood pressure, and resting heart rate measurement, and provided extensive information on demographic characteristics, genealogy, education, occupation, and lifestyle, as well as health status and medical history (Leitsalu et al., 2014, 2015). From 2017 onward, the process of joining ESTBB was simplified: new joiners were required to sign an online consent form, visit a healthcare provider or pharmacy to provide a blood sample, and fill in an online questionnaire. Additional data are collected through linkage with various national databases and registries and through subsequent studies involving different subsets of participants.

Participants were genotyped using the Illumina Global Screening Array (GSA) v1.0, v2.0, and v2.0_EST. Samples were genotyped and PLINK format files created using Illumina GenomeStudio v2.0.4. Individuals with call rates below 95% or whose sex in phenotype data did not match X chromosome heterozygosity were excluded. Before imputation, variants were filtered by call rate <95%, Hardy–Weinberg equilibrium *p*-value < $1e^{-4}$ (autosomal variants only), and minor allele frequency <1%. A population-specific imputation reference panel of 2,297 WGS samples was used (Mitt et al., 2017).

We aligned participants to 21 major ancestry groups similarly to Privé (2021). We removed participants with non-European assigned group ancestry, keeping Europeans, Finns and Italians. https://doi.org/10.1093/bioinformatics/btac348.

#### Helsinki University Hospital / Helsinki Biobank (HUS)

The Helsingin Biopankki (Helsinki Biobank) was founded in 2015 by HUS Helsinki University Hospital, University of Helsinki, the Joint Municipality for Health and Social Services in the Kymenlaakso Valley (Kymsote) and the South Karelia Social and Health Care District (Exote), and is the largest hospital biobank in Finland. Approximately 80,000 patients have given a blood sample based on biobank consent and used for research purposes. Diagnostic formalin-fixed paraffin-embedded tissue samples (FFPE) from approximately one million patients have been transferred to the biobank's sample collection. The biobank's samples were combined with clinical patient data from the hospital's database (HUS Datapool) in accordance with the description in the research plan recommended by the ethics committee. Data that are run together can usually include the sampling date, the donor's gender, histological and cytological examinations, diagnoses, care measures and laboratory examination results.

#### Mass General Brigham Biobank (MGBB)

Mass General Brigham is an integrated healthcare system located in Boston, Massachusetts for more than 1.5 million individuals per year. The Mass General Brigham Biobank (MGBB) has recruited over 140,000 participants since 2008. Over 65,000 participants have genotype data sequenced using the Illumina Multi-Ethnic Genotyping MEG) assay or the Illumina Global Screening Array (GSA). All genotyped biospecimens had at least 99% call rate per array and were confirmed for sex concordance using sex in EHR and sex computed from array data. Ancestry was defined using high-dimensional principal components, specifically top 30 genetic PCs and a network-based clustering approach. We restricted participants to an unrelated subset. The participants were further filtered based on the criteria described in the analysis plan (e.g., per ancestry minimum allele count/sample size after filtering on availability of weight measurements and prescription and/or bariatric surgery).

#### UCLA-ATLAS (Atlas Biobank)

ATLAS is an EHR-linked biobank across multiple institutions within the UCLA Health System. After participants signed informed consent for participation to the UCLA ATLAS Community Health Initiative, biological samples in the form of de-identified blood samples were collected and subsequently processed for DNA extraction and genotyping. The data analyzed in this manuscript draws from a “frozen snapshot” of ATLAS data encompassing all available samples up to February 15, 2024, totaling N=53829. The corresponding EHR data for ATLAS participants was sourced from the UCLA Data Discovery Repository (DDR). This repository operates under the auspices of the UCLA Health Office of Health Informatics Analytics and the UCLA Institute of Precision Health. Patient Recruitment and Sample Collection for Precision Health Activities at UCLA is an approved study by the UCLA Institutional Review Board. Extensive details on ATLAS genotyping and quality control were previously reported in [PMID: 36777178]. In short, participants in the ATLAS initiative were genotyped using a custom genotyping array constructed from the Global Screening Array with the Infinium Global Screening Array-24 Kit (Illumina, San Diego, California), a multi-disease drop-in panel aligned with the GRCh38 assembly. SNPs were subjected to removal if they were unmapped, strand-ambiguous, duplicates, or exhibited >5% missingness. Samples with missingness exceeding >5% were excluded, as were duplicates (including identical twins or triplets) with preference given to individuals with the lowest missing rate in each pair. Imputation to the TOPMedFreeze5 was conducted using the Michigan Imputation Server [PMID: 27571263]. After imputation, SNPs with a quality score (R2) below 0.90 were excluded, for polygenic score calculation SNPs with a MAF below 0.1% were filtered out. The genetic ancestry of ATLAS individuals was estimated by assessing their proximity to 1000 Genome super populations on the principal component (PC) space. We computed the top 20 PCs using the bigsnpr R software with default parameters. Subsequently, leveraging the super population label and PCs of the 1000 Genome individuals, we train a K-nearest neighbors model to assign genetic ancestry labels to each ATLAS individual. To optimize model performance, we employed 10-fold cross-validation to select the optimal hyper-parameter (k) from a range of values (k = 5, 10, 15, 20). In instances where an individual was assigned to multiple ancestries with probability larger than 0.5 or was not assigned to any cluster, their ancestry was labelled as unknown. We clustered 94% of the ATLAS participants into one of the five 1000 Genome super population (AFR, AMR, EAS, EUR, SAS) while the remaining individuals' ancestry could not be ascertained.

#### UK Biobank (UKBB)

The UK Biobank (UKBB) is a large-scale biomedical database and research resource containing in-depth genetic and health information from half a million UK participants. Upon signing informed consent, participants provided biological samples, including de-identified blood samples, which were processed for DNA extraction and genotyping. The dataset analyzed in this manuscript is derived from a "frozen snapshot" of the UKBB data, encompassing all available samples up to January 31, 2024, totaling N=502,536. Corresponding electronic health record (EHR) data for UKBB participants were sourced from the UK Biobank Resource, which operates under the auspices of the UK Biobank Steering Committee and has received ethical approval from the North West Multi-centre Research Ethics Committee. Detailed information on UKBB genotyping and quality control has been previously reported in https://biobank.ctsu.ox.ac.uk/crystal/crystal/docs/genotyping_qc.pdf]. Samples were genotyped at the Affymetrix Research Services Laboratory in Santa Clara, California, USA. Upon receipt of a 96-well plate containing 94 UK Biobank samples, Affymetrix added two control individuals (from 1000 Genomes) to the same well positions on each plate. Genotypes were then called from the resulting intensities in batches of ~4,700 samples (~4,800 including the controls) using the Affymetrix Power Tools software and the Affymetrix Best Practices Workflow. After genotype calling, Affymetrix performed quality control in each batch separately, to exclude SNPs with poor cluster properties. If a SNP did not meet the Affymetrix prescribed QC thresholds in a given batch, it was set to missing in all individuals from that batch. Affymetrix also checked sample quality (such as DNA concentration) and genotype calls were provided only for samples with sufficient DNA metrics.

The ancestry composition of the UK Biobank participants was defined based on self-reported ethnicities and genetic principal component analysis (PCA). Participants self-reported their ethnic backgrounds into categories such as White, Mixed, Asian or Asian British, Black or Black British, and Other ethnic groups, with the majority (~94%) identifying as White British. Genetic PCA was used to capture the underlying population structure and define genetic ancestry more precisely, categorizing participants into groups such as European, South Asian, African, East Asian, and other ancestries. This approach ensured a nuanced understanding of the genetic and demographic diversity within the cohort, enabling robust and reliable biomedical research.

To capture population structure specific to the UK Biobank cohort, we performed principal component analysis of ~150,000 UK Biobank samples using ~100,000 SNPs. These PCs can be used to identify samples with similar ancestry or to control for population structure in association studies.

#### Qatar Biobank (QBB)

This study was based on the second release of 14,060 Qatari participants (all from the same ancestry, Middle-Eastern - MID) from the population based Qatar Genome Program (QGP). Ethical approval was provided by the institutional review board of the Qatar Biobank (QBB) (Protocol no. QF-QBB-RES-ACC-00205). A detailed description of the recruitment process and collection of phenotypic data by the QGP and the QBB was previously reported at https://doi.org/10.1093/aje/kwz084. A signed consent was obtained from all included participants by the QBB, and WGS data used in the current study was obtained from the QGP as previously described (https://doi.org/10.1002/humu.24336). All samples were sequenced on Illumina HiSeq X instruments (Illumina, San Diego, CA, USA) to obtain WGS with a target average depth of coverage of 30x. The read mapping, variant calling and joint variant calling were performed using Sentieon’s DNASeq pipeline v201808.03 (Sentieon, San Jose, CA, USA), following the BWA-GATK Best Practice Workflow and using the GRCh38/hg38 reference genome. The produced gVCF were jointly called to produce one multisample VCF file (msVCF) for the whole cohort. Clinical data are extracted from the EMR of the Hamad Medical Corporation (HMC).
