## Supplementary Tables for "Association between plausible genetic factors and weight loss from GLP1-RA and bariatric surgery: a multi-ancestry study in 10 960 individuals from 9 biobanks"

**Supplementary Table 1 | Heterogeneity statistics for baseline variables in GLP1 meta analysis (Model A).**
Heterogeneity across ancestries and studies for the tested genetic exposures. Cochran's Q (P-Value) and the I^2 statistics are reported in the table.

| Baseline variable | Q coefficient | P-value Q coefficient | I2 |
|---|---|---|---|
| **Baseline weight** | 362.590534 | 3.40E-76 | 0.9862103 |
| **Sex** | 29.251478 | 2.07E-05 | 0.8290685 |
| **Age at baseline** | 2.660835 | 0.7521017 | 0 |

*Studies with EUR ancestry only:*

| Baseline variable | Q coefficient | P-value Q coefficient | I2 |
|---|---|---|---|
| **Baseline weight** | 10.071329 | 0.1216802 | 0.4042494 |
| **Sex** | 1.139853 | 0.9797513 | 0 |
| **Age at baseline** | 5.177059 | 0.521313 | 0 |

**Supplementary Table 2 | Heterogeneity statistics for baseline variables in BS meta analysis (Model A).**
Heterogeneity across ancestries and studies for the tested genetic exposures. Cochran's Q (P-Value) and the I^2 statistics are reported in the table.

| Baseline variable | Q coefficient | P-value Q coefficient | I2 |
|---|---|---|---|
| **Baseline weight** | 72.8537968 | 1.04E-15 | 0.9588216 |
| **Sex** | 0.5087383 | 0.9169675 | 0 |
| **Age at baseline** | 0.6844168 | 0.8768630 | 0 |

*Studies with EUR ancestry only:*

| Baseline variable | Q coefficient | P-value Q coefficient | I2 |
|---|---|---|---|
| **Baseline weight** | 66.67497 | 1.96E-12 | 0.9100112 |
| **Sex** | 22.10186 | 1.16E-03 | 0.7285296 |
| **Age at baseline** | 7.784325 | 0.2543339 | 0.2292203 |

**Supplementary Table 3 | Polygenic score (PGS) - phenotype associations.**
Associations between PGS and the respective phenotype for each ancestry. The table contains the R-squared (R2), N, Beta and P-val for the models.
The baseline model included age, sex and top 20 principal components as covariates. We included the following studies: UCLA-ATLAS Biobank (N= 33,253), Qatar Biobank (N=2,258)
In ATLAS, the association between PGS and the corresponding trait were tested in the entire biobank, while in Qatar Biobank only in individuals that were included in the analyses

| | | R2 | | | | | | Regression coefficients (P-val) | | | | | |
|---|---|---|---|---|---|---|---|---|---|---|---|---|---|
| | | **AFR** | **AMR** | **EAS** | **EUR** | **SAS** | **MID** | **AFR** | **AMR** | **EAS** | **EUR** | **SAS** | **MID** |
| **T2D** | **Baseline** | 0.0882 | 0.152 | 0.1553 | 0.0712 | 0.2416 | 0.1903 | | | | | | |
| | **T2D PGS** | 0.1007 | 0.1677 | 0.1703 | 0.0955 | 0.2537 | 0.2138 | 0.202 (p<0.0001) | 0.35 (p<0.0001) | 0.338 (p<0.0001) | 0.424 (p<0.0001) | 0.333 (p=0.0008) | 0.509 (p<0.0001) |
| | **N (cases/controls)** | 800/1781 | 2037/5435 | 1191/3707 | 4670/28583 | 211/639 | 1321/937 | 800/1781 | 2037/5435 | 1191/3707 | 4670/28583 | 211/639 | 1321/937 |
| **BMI** | **Baseline** | 0.0244 | 0.0022 | 0.0948 | 0.0161 | 0.0231 | 0.0301 | | | | | | |
| | **BMI PGS** | 0.0459 | 0.0031 | 0.1221 | 0.1102 | 0.0772 | 0.0410 | 1.12 (p<0.0001) | 1.59 (0.0075) | 0.894 (p<0.0001) | 1.747 (p<0.0001) | 1.098 (p<0.0001) | 0.9023 (p<0.000 |
| | **N** | 2581 | 7472 | 4898 | 33253 | 850 | 2258 | 2581 | 7472 | 4898 | 33253 | 850 | 2258 |

| | | |
|---|---|---|
| Abbreviations | R2 | R squared |
| | AFR | African |
| | AMR | American |
| | EAS | East Asian |
| | EUR | European |
| | SAS | South Asian |
| | MID | Middle Eastern |
| | T2D | Type 2 diabetes |
| | BMI | Body Mass Index |
| | PGS | Polygenic scores |

**Supplementary Table 4 | Genetic exposures and their frequencies across al major continental ancestry groups.**
This table contains the a description of the genetic variants, including chromosome position, reference (Ref) allele, alternative (Alt) allele,
the associated gene, if applicable, a publication, and frequencies across all major continental ancestry groups, derived from GnomAD (version 4.1)

| Genetic exposure (rsID) | Chr | Position (b38) | Ref | Alt | Variant Effect Predictor Annotation | HGVS Nomenclature | Association | Publication/ Reference | Author and year |
|---|---|---|---|---|---|---|---|---|---|
| rs10305492 | 6 | 39079018 | G | A | GoF variant | NC_000006.12:g.39079018G>A | GLP1R | https://doi.org/10.1038/ | Lagou et al. 2023 |
| rs146868158 | 6 | 39085942 | C | T | LoF variant | NC_000006.12:g.39085942C>T | GLP1R | https://doi.org/10.1038/ | Lagou et al. 2023 |
| rs6923761 | 6 | 39066296 | G | A | GoF variant | NC_000006.12:g.39066296G>A | GLP1R | https://doi.org/10.1038/ | Lagou et al. 2023 |
| rs1042044 | 6 | 39073726 | A | C | Missense variant | NC_000006.12:g.39073726A>C | GLP1R | GLP1R missense variants from GnomAD | |
| rs10305420 | 6 | 39048860 | C | T | Missense variant | NC_000006.12:g.39048860C>T | GLP1R | GLP1R missense variants from GnomAD | |
| rs3765467 | 6 | 39065819 | G | A | Missense variant | NC_000006.12:g.39065819G>A | GLP1R | GLP1R missense variants from GnomAD | |
| rs10305421 | 6 | 39048899 | G | A | Missense variant | NC_000006.12:g.39048899G>A | GLP1R | GLP1R missense variants from GnomAD | |
| rs2295006 | 6 | 39056449 | G | A | Missense variant | NC_000006.12:g.39056449G>A | GLP1R | GLP1R missense variants from GnomAD | |
| rs10305510 | 6 | 39085943 | G | A | Missense variant | NC_000006.12:g.39085943G>A | GLP1R | GLP1R missense variants from GnomAD | |
| rs201672448 | 6 | 39086015 | G | C | Missense variant | NC_000006.12:g.39086015G>C | GLP1R | GLP1R missense variants from GnomAD | |
| rs429358 | 19 | 44908684 | T | C | Missense variant | NC_000019.10:g.44908684T>C | High BMI/weight gain | https://doi.org/10.1101/ | Venkatesh et al. 2023 |
| rs6232 | 5 | 96416081 | T | C | Missense variant | NC_000005.10:g.96416081T>C | PCSK1 | https://doi.org/10.1093/ | Nead et al. 2015 |
| rs6235 | 5 | 96393194 | C | G | Missense variant | NC_000005.10:g.96393194C>G | PCSK1 | https://doi.org/10.1093/ | Nead et al. 2015 |
| rs17702901 | 15 | 92387181 | G | A | - | NC_000015.10:g.92387181G>A | ST8SIA2 | https://doi.org/10.1007/ | Aasbrenn et al. 2019 |
| rs728996 | 6 | 51778216 | C | T | Intron | NC_000006.12:g.51778216C>T, | PKHD1 | https://doi.org/10.1007/ | Aasbrenn et al. 2019 |

| Genetic exposure (rsID) | Total Allele Count | Total Allele Number | Total Allele Frequency | Allele Count Admixed American | Allele Number Admixed American | Allele Frequency Admixed American | Allele Count European (Finnish) | Allele Number European (Finnish) | Allele Frequency European (Finnish) | Allele Count Middle Eastern | Allele Number Middle Eastern | Allele Frequency Middle Eastern |
|---|---|---|---|---|---|---|---|---|---|---|---|---|
| rs10305492 | 21639 | 1613926 | 0.013 | 495 | 60008 | 0.008 | 855 | 64034 | 0.013 | 15 | 6062 | 0.002 |
| rs146868158 | 732 | 1613782 | 0.000 | 2 | 59982 | 0.000 | 99 | 64028 | 0.002 | 0 | 6080 | 0.000 |
| rs6923761 | 466030 | 1594086 | 0.292 | 4284 | 74710 | 0.057 | 17469 | 63660 | 0.274 | 1443 | 6046 | 0.239 |
| rs1042044 | 908991 | 1613212 | 0.563 | 36180 | 60004 | 0.603 | 35044 | 63962 | 0.548 | 3430 | 6672 | 0.514 |
| rs10305420 | 527575 | 1492840 | 0.353 | 13779 | 49372 | 0.279 | 17878 | 44662 | 0.400 | 1996 | 5544 | 0.360 |
| rs3765467 | 24714 | 1592830 | 0.016 | 2541 | 59262 | 0.043 | 13 | 63690 | 0.000 | 133 | 6662 | 0.020 |
| rs10305421 | 6846 | 1440018 | 0.005 | 162 | 39530 | 0.004 | 54 | 43542 | 0.001 | 8 | 5124 | 0.002 |
| rs2295006 | 2153 | 1610672 | 0.001 | 7 | 60010 | 0.000 | 6 | 64028 | 0.000 | 1 | 6668 | 0.000 |
| rs10305510 | 1806 | 1613846 | 0.001 | 51 | 60008 | 0.001 | 8 | 64020 | 0.000 | 3 | 6678 | 0.000 |
| rs201672448 | 1436 | 1614086 | 0.001 | 1 | 60018 | 0.000 | 0 | 64042 | 0.000 | 3 | 6678 | 0.000 |
| rs429358 | 232864 | 1567892 | 0.149 | 5611 | 52482 | 0.107 | 11533 | 60090 | 0.192 | 404 | 5934 | 0.068 |
| rs6232 | 72726 | 1612350 | 0.045 | 1154 | 60024 | 0.019 | 2058 | 64026 | 0.032 | 323 | 6056 | 0.053 |
| rs6235 | 424550 | 1613804 | 0.263 | 11890 | 60000 | 0.198 | 18131 | 63972 | 0.283 | 1426 | 6062 | 0.235 |
| rs17702901 | 8528 | 152246 | 0.056 | 2403 | 15304 | 0.157 | 464 | 10606 | 0.044 | 10 | 294 | 0.034 |
| rs728996 | 86882 | 151936 | 0.572 | 9954 | 15226 | 0.654 | 6717 | 10540 | 0.637 | 165 | 294 | 0.561 |

| Genetic exposure (rsID) | Allele Count European (non-Finnish) | Allele Number European (non-Finnish) | Allele Frequency European (non-Finnish) | Allele Count South Asian | Allele Number South Asian | Allele Frequency South Asian | Allele Count Ashkenazi Jewish | Allele Number Ashkenazi Jewish | Allele Frequency Ashkenazi Jewish | Allele Count East Asian | Allele Number East Asian | Allele Frequency East Asian | Allele Count African/African American | Allele Number African/African American | Allele Frequency African/African American |
|---|---|---|---|---|---|---|---|---|---|---|---|---|---|---|---|
| rs10305492 | 18902 | 1179808 | 0.016 | 91 | 91072 | 0.001 | 351 | 29604 | 0.012 | 2 | 44878 | 0.000 | 195 | 75040 | 0.003 |
| rs146868158 | 595 | 1179876 | 0.001 | 3 | 91070 | 0.000 | 1 | 29604 | 0.000 | 2 | 44882 | 0.000 | 8 | 74882 | 0.000 |
| rs6923761 | 396349 | 1161864 | 0.341 | 13510 | 90720 | 0.149 | 6991 | 29480 | 0.237 | 228 | 44794 | 0.005 | 4284 | 74710 | 0.057 |
| rs1042044 | 631461 | 1116620 | 0.566 | 49705 | 86542 | 0.574 | 14725 | 30920 | 0.476 | 22457 | 43162 | 0.520 | 43033 | 74900 | 0.575 |
| rs10305420 | 411741 | 1060544 | 0.388 | 15169 | 75972 | 0.200 | 9896 | 28484 | 0.347 | 6354 | 33690 | 0.189 | 5026 | 68708 | 0.073 |
| rs3765467 | 4335 | 1100328 | 0.004 | 7341 | 84810 | 0.087 | 496 | 30748 | 0.016 | 8120 | 42876 | 0.189 | 177 | 74678 | 0.002 |
| rs10305421 | 5418 | 1037148 | 0.005 | 3 | 65784 | 0.000 | 410 | 24884 | 0.016 | 76 | 33484 | 0.002 | 60 | 67190 | 0.001 |
| rs2295006 | 262 | 1114098 | 0.000 | 141 | 86462 | 0.002 | 18 | 30924 | 0.001 | 1612 | 43132 | 0.037 | 0 | 74944 | 0.000 |
| rs10305510 | 104 | 1117032 | 0.000 | 2 | 86540 | 0.000 | 1 | 30946 | 0.000 | 16 | 43172 | 0.000 | 1549 | 74952 | 0.021 |
| rs201672448 | 6 | 1117130 | 0.000 | 1293 | 86546 | 0.015 | 71 | 30958 | 0.002 | 3 | 43168 | 0.000 | 0 | 75038 | 0.000 |
| rs429358 | 174295 | 1157104 | 0.151 | 8686 | 86316 | 0.101 | 3458 | 28748 | 0.120 | 4094 | 41846 | 0.098 | 16245 | 73686 | 0.220 |
| rs6232 | 59588 | 1178326 | 0.051 | 5966 | 91044 | 0.066 | 499 | 29594 | 0.017 | 28 | 44868 | 0.001 | 648 | 75048 | 0.009 |
| rs6235 | 316942 | 1179872 | 0.269 | 27643 | 91054 | 0.304 | 8613 | 29602 | 0.291 | 12066 | 44850 | 0.269 | 11300 | 74974 | 0.151 |
| rs17702901 | 2316 | 68004 | 0.034 | 572 | 4824 | 0.119 | 83 | 3470 | 0.024 | 732 | 5180 | 0.141 | 1800 | 41538 | 0.043 |
| rs728996 | 41933 | 67952 | 0.617 | 3565 | 4824 | 0.739 | 1884 | 3466 | 0.544 | 4277 | 5160 | 0.829 | 16759 | 41452 | 0.404 |

**Supplementary Table 5 | Meta analysis results for GLP1-RA (primary model).**
Ancestry-specific and multi-ancestry meta-analyses of the GLP1-RA analysis.

| | | | Main genetic exposure | | | Baseline body weight | | | Sex | | | Age at initiation | | |
|---|---|---|---|---|---|---|---|---|---|---|---|---|---|---|
| **Study** | **Ancestry** | **Model (main exposure)** | **Beta** | **Standard Error** | **P-value** | **Beta** | **Standard Error** | **P-value** | **Beta** | **Standard Error** | **P-value** | **Beta** | **Standard Error** | **P-value** |
| **Meta_AFR** | AFR | BMI PGS | -0.16 | 0.21 | 0.45 | -0.01 | 0.01 | 0.09 | -0.33 | 0.44 | 0.46 | -0.01 | 0.02 | 0.41 |
| **Meta_AFR** | AFR | T2D PGS | 0.41 | 0.22 | 0.07 | -0.01 | 0.01 | 0.10 | -0.27 | 0.45 | 0.55 | -0.01 | 0.02 | 0.54 |
| **Meta_AFR** | AFR | rs10305492 | -7.63 | 5.68 | 0.18 | 0.00 | 0.03 | 0.89 | -0.77 | 1.41 | 0.58 | -0.13 | 0.05 | 0.02 |
| **Meta_AFR** | AFR | rs146868158 | | | | | | | | | | | | |
| **Meta_AFR** | AFR | rs6923761 | -0.53 | 0.54 | 0.32 | -0.02 | 0.01 | 0.05 | -0.35 | 0.45 | 0.44 | -0.01 | 0.02 | 0.49 |
| **Meta_AFR** | AFR | rs1042044 | 0.06 | 0.27 | 0.82 | -0.01 | 0.01 | 0.08 | -0.35 | 0.45 | 0.44 | -0.01 | 0.02 | 0.45 |
| **Meta_AFR** | AFR | rs10305420 | -0.17 | 0.55 | 0.76 | -0.02 | 0.01 | 0.04 | -0.66 | 0.47 | 0.16 | -0.02 | 0.02 | 0.22 |
| **Meta_AFR** | AFR | rs3765467 | -0.57 | 2.02 | 0.78 | -0.01 | 0.01 | 0.14 | -0.59 | 0.52 | 0.26 | -0.01 | 0.02 | 0.59 |
| **Meta_AFR** | AFR | rs10305421 | -6.80 | 5.45 | 0.21 | 0.00 | 0.03 | 0.94 | -0.39 | 1.42 | 0.79 | -0.12 | 0.05 | 0.04 |
| **Meta_AFR** | AFR | rs10305510 | 1.52 | 1.07 | 0.16 | -0.02 | 0.01 | 0.06 | -0.62 | 0.47 | 0.19 | -0.02 | 0.02 | 0.24 |
| **Meta_AFR** | AFR | rs429358 | -0.61 | 0.67 | 0.36 | -0.02 | 0.02 | 0.22 | 0.16 | 0.89 | 0.86 | -0.02 | 0.03 | 0.61 |
| **Meta_AFR** | AFR | rs6232 | 2.07 | 1.54 | 0.18 | -0.02 | 0.01 | 0.05 | -0.64 | 0.47 | 0.17 | -0.02 | 0.02 | 0.21 |
| **Meta_AFR** | AFR | rs6235 | 0.24 | 0.37 | 0.53 | -0.02 | 0.01 | 0.07 | -0.37 | 0.44 | 0.40 | -0.01 | 0.02 | 0.53 |
| **Meta_AFR** | AFR | Baseline | | | | -0.01 | 0.01 | 0.13 | -0.23 | 0.43 | 0.59 | -0.01 | 0.02 | 0.43 |
| **Meta_AMR** | AMR | BMI PGS | -0.12 | 0.19 | 0.53 | -0.01 | 0.01 | 0.30 | -0.56 | 0.42 | 0.18 | -0.02 | 0.02 | 0.24 |
| **Meta_AMR** | AMR | T2D PGS | -0.02 | 0.20 | 0.91 | -0.01 | 0.01 | 0.23 | -0.57 | 0.42 | 0.18 | -0.02 | 0.02 | 0.28 |
| **Meta_AMR** | AMR | rs10305492 | -0.57 | 1.45 | 0.69 | -0.01 | 0.01 | 0.25 | -0.56 | 0.42 | 0.18 | -0.02 | 0.02 | 0.25 |
| **Meta_AMR** | AMR | rs146868158 | | | | | | | | | | | | |
| **Meta_AMR** | AMR | rs6923761 | -0.09 | 0.38 | 0.81 | -0.01 | 0.01 | 0.25 | -0.56 | 0.42 | 0.18 | -0.02 | 0.02 | 0.25 |
| **Meta_AMR** | AMR | rs1042044 | -0.13 | 0.27 | 0.63 | -0.01 | 0.01 | 0.26 | -0.55 | 0.42 | 0.19 | -0.02 | 0.02 | 0.24 |
| **Meta_AMR** | AMR | rs10305420 | 0.15 | 0.37 | 0.68 | -0.01 | 0.01 | 0.34 | -0.51 | 0.50 | 0.31 | -0.02 | 0.02 | 0.27 |
| **Meta_AMR** | AMR | rs3765467 | 0.24 | 0.81 | 0.77 | -0.01 | 0.01 | 0.23 | -0.57 | 0.42 | 0.17 | -0.02 | 0.02 | 0.25 |
| **Meta_AMR** | AMR | rs10305421 | -0.99 | 1.89 | 0.60 | -0.01 | 0.01 | 0.31 | -0.49 | 0.50 | 0.33 | -0.03 | 0.02 | 0.24 |
| **Meta_AMR** | AMR | rs10305510 | 1.67 | 2.11 | 0.43 | -0.01 | 0.01 | 0.31 | -0.48 | 0.50 | 0.33 | -0.02 | 0.02 | 0.25 |
| **Meta_AMR** | AMR | rs429358 | 0.39 | 0.80 | 0.63 | -0.01 | 0.02 | 0.52 | -0.71 | 0.77 | 0.36 | -0.01 | 0.03 | 0.73 |
| **Meta_AMR** | AMR | rs6232 | -0.89 | 0.79 | 0.26 | -0.01 | 0.01 | 0.25 | -0.54 | 0.42 | 0.20 | -0.02 | 0.02 | 0.24 |
| **Meta_AMR** | AMR | rs6235 | 0.18 | 0.33 | 0.58 | -0.01 | 0.01 | 0.23 | -0.58 | 0.42 | 0.17 | -0.02 | 0.02 | 0.25 |
| **Meta_AMR** | AMR | Baseline | | | | -0.01 | 0.01 | 0.47 | -0.51 | 0.41 | 0.22 | -0.01 | 0.02 | 0.46 |
| **Meta_EAS** | EAS | BMI PGS | 0.06 | 0.64 | 0.92 | 0.00 | 0.05 | 0.97 | -1.57 | 1.40 | 0.26 | -0.01 | 0.05 | 0.87 |
| **Meta_EAS** | EAS | T2D PGS | 0.71 | 0.60 | 0.23 | -0.01 | 0.05 | 0.86 | -1.63 | 1.38 | 0.24 | 0.00 | 0.05 | 0.98 |
| **Meta_EAS** | EAS | rs10305492 | | | | | | | | | | | | |
| **Meta_EAS** | EAS | rs146868158 | | | | | | | | | | | | |
| **Meta_EAS** | EAS | rs6923761 | | | | | | | | | | | | |
| **Meta_EAS** | EAS | rs1042044 | -0.43 | 0.88 | 0.63 | 0.00 | 0.05 | 0.96 | -1.48 | 1.40 | 0.29 | -0.01 | 0.05 | 0.82 |
| **Meta_EAS** | EAS | rs10305420 | | | | | | | | | | | | |
| **Meta_EAS** | EAS | rs3765467 | -0.51 | 1.07 | 0.63 | 0.00 | 0.05 | 0.95 | -1.47 | 1.40 | 0.29 | -0.01 | 0.05 | 0.82 |
| **Meta_EAS** | EAS | rs10305421 | | | | | | | | | | | | |
| **Meta_EAS** | EAS | rs10305510 | | | | | | | | | | | | |
| **Meta_EAS** | EAS | rs429358 | | | | | | | | | | | | |
| **Meta_EAS** | EAS | rs6232 | | | | | | | | | | | | |
| **Meta_EAS** | EAS | rs6235 | 0.02 | 0.93 | 0.98 | 0.00 | 0.05 | 0.98 | -1.56 | 1.39 | 0.26 | -0.01 | 0.05 | 0.87 |
| **Meta_EAS** | EAS | Baseline | | | | 0.00 | 0.04 | 0.94 | -1.01 | 1.20 | 0.40 | 0.01 | 0.05 | 0.92 |
| **Meta_EUR** | EUR | BMI PGS | 0.06 | 0.13 | 0.63 | -0.04 | 0.01 | 0.00 | -2.23 | 0.28 | 0.00 | -0.02 | 0.01 | 0.12 |
| **Meta_EUR** | EUR | T2D PGS | 0.01 | 0.14 | 0.92 | -0.04 | 0.01 | 0.00 | -2.25 | 0.28 | 0.00 | -0.02 | 0.01 | 0.11 |
| **Meta_EUR** | EUR | rs10305492 | 0.69 | 0.89 | 0.44 | -0.04 | 0.01 | 0.00 | -2.25 | 0.31 | 0.00 | -0.03 | 0.01 | 0.03 |
| **Meta_EUR** | EUR | rs146868158 | 3.48 | 3.80 | 0.36 | -0.04 | 0.02 | 0.03 | -1.90 | 0.87 | 0.03 | -0.02 | 0.03 | 0.47 |
| **Meta_EUR** | EUR | rs6923761 | -0.19 | 0.20 | 0.36 | -0.04 | 0.01 | 0.00 | -2.23 | 0.28 | 0.00 | -0.02 | 0.01 | 0.14 |
| **Meta_EUR** | EUR | rs1042044 | 0.16 | 0.19 | 0.39 | -0.04 | 0.01 | 0.00 | -2.19 | 0.28 | 0.00 | -0.02 | 0.01 | 0.17 |
| **Meta_EUR** | EUR | rs10305420 | 0.00 | 0.22 | 1.00 | -0.04 | 0.01 | 0.00 | -2.31 | 0.33 | 0.00 | -0.02 | 0.02 | 0.23 |
| **Meta_EUR** | EUR | rs3765467 | 1.71 | 1.49 | 0.25 | -0.04 | 0.01 | 0.00 | -2.27 | 0.32 | 0.00 | -0.03 | 0.01 | 0.03 |
| **Meta_EUR** | EUR | rs10305421 | 0.49 | 1.06 | 0.65 | -0.04 | 0.01 | 0.00 | -2.25 | 0.29 | 0.00 | -0.03 | 0.01 | 0.05 |

| | | | | | | | | | | | | | | |
|---|---|---|---|---|---|---|---|---|---|---|---|---|---|---|
| **Meta_EUR** | EUR | rs10305510 | | | | | | | | | | | | |
| **Meta_EUR** | EUR | rs429358 | -0.32 | 0.31 | 0.31 | -0.04 | 0.01 | 0.00 | -2.30 | 0.31 | 0.00 | -0.02 | 0.01 | 0.13 |
| **Meta_EUR** | EUR | rs6232 | 0.33 | 0.44 | 0.45 | -0.04 | 0.01 | 0.00 | -2.22 | 0.28 | 0.00 | -0.02 | 0.01 | 0.13 |
| **Meta_EUR** | EUR | rs6235 | 0.19 | 0.20 | 0.34 | -0.04 | 0.01 | 0.00 | -2.20 | 0.28 | 0.00 | -0.02 | 0.01 | 0.15 |
| **Meta_EUR** | EUR | Baseline | | | | -0.03 | 0.01 | 0.00 | -2.18 | 0.28 | 0.00 | -0.02 | 0.01 | 0.16 |
| **Meta_SAS** | SAS | BMI PGS | 0.15 | 0.90 | 0.87 | -0.10 | 0.05 | 0.04 | -4.24 | 2.06 | 0.04 | -0.13 | 0.07 | 0.07 |
| **Meta_SAS** | SAS | T2D PGS | 1.02 | 0.82 | 0.22 | -0.09 | 0.04 | 0.03 | -4.22 | 2.02 | 0.04 | -0.12 | 0.07 | 0.10 |
| **Meta_SAS** | SAS | rs10305492 | | | | | | | | | | | | |
| **Meta_SAS** | SAS | rs146868158 | | | | | | | | | | | | |
| **Meta_SAS** | SAS | rs6923761 | -2.07 | 1.76 | 0.24 | -0.11 | 0.04 | 0.02 | -4.85 | 2.09 | 0.02 | -0.16 | 0.07 | 0.03 |
| **Meta_SAS** | SAS | rs1042044 | 1.09 | 1.09 | 0.32 | -0.09 | 0.04 | 0.03 | -4.00 | 2.05 | 0.05 | -0.13 | 0.07 | 0.07 |
| **Meta_SAS** | SAS | rs10305420 | -1.21 | 1.45 | 0.40 | -0.09 | 0.04 | 0.04 | -4.29 | 2.04 | 0.04 | -0.13 | 0.07 | 0.08 |
| **Meta_SAS** | SAS | rs3765467 | 0.34 | 2.03 | 0.87 | -0.09 | 0.04 | 0.03 | -4.20 | 2.08 | 0.04 | -0.14 | 0.07 | 0.06 |
| **Meta_SAS** | SAS | rs10305421 | | | | | | | | | | | | |
| **Meta_SAS** | SAS | rs10305510 | | | | | | | | | | | | |
| **Meta_SAS** | SAS | rs429358 | | | | | | | | | | | | |
| **Meta_SAS** | SAS | rs6232 | -2.62 | 3.04 | 0.39 | -0.09 | 0.04 | 0.03 | -3.86 | 2.09 | 0.06 | -0.12 | 0.07 | 0.09 |
| **Meta_SAS** | SAS | rs6235 | -2.49 | 1.66 | 0.13 | -0.09 | 0.04 | 0.04 | -4.65 | 2.02 | 0.02 | -0.11 | 0.07 | 0.11 |
| **Meta_SAS** | SAS | Baseline | | | | -0.08 | 0.04 | 0.03 | -4.07 | 1.45 | 0.00 | -0.10 | 0.06 | 0.07 |
| **Meta_MID** | MID | BMI PGS | 0.69 | 0.28 | 0.01 | -0.26 | 0.01 | 0.00 | -2.96 | 0.61 | 0.00 | 0.00 | 0.03 | 0.87 |
| **Meta_MID** | MID | T2D PGS | -0.02 | 0.33 | 0.96 | -0.25 | 0.01 | 0.00 | -2.92 | 0.61 | 0.00 | -0.01 | 0.03 | 0.78 |
| **Meta_MID** | MID | rs10305492 | | | | | | | | | | | | |
| **Meta_MID** | MID | rs146868158 | | | | | | | | | | | | |
| **Meta_MID** | MID | rs6923761 | -0.17 | 0.44 | 0.70 | -0.25 | 0.01 | 0.00 | -2.92 | 0.61 | 0.00 | -0.01 | 0.03 | 0.77 |
| **Meta_MID** | MID | rs1042044 | -0.27 | 0.38 | 0.48 | -0.25 | 0.01 | 0.00 | -2.93 | 0.61 | 0.00 | -0.01 | 0.03 | 0.75 |
| **Meta_MID** | MID | rs10305420 | -0.08 | 0.38 | 0.83 | -0.25 | 0.01 | 0.00 | -2.91 | 0.61 | 0.00 | -0.01 | 0.03 | 0.78 |
| **Meta_MID** | MID | rs3765467 | 0.08 | 1.64 | 0.96 | -0.25 | 0.01 | 0.00 | -2.92 | 0.61 | 0.00 | -0.01 | 0.03 | 0.78 |
| **Meta_MID** | MID | rs10305421 | | | | | | | | | | | | |
| **Meta_MID** | MID | rs10305510 | | | | | | | | | | | | |
| **Meta_MID** | MID | rs429358 | 0.28 | 0.64 | 0.66 | -0.25 | 0.01 | 0.00 | -2.92 | 0.61 | 0.00 | -0.01 | 0.03 | 0.77 |
| **Meta_MID** | MID | rs6232 | 0.21 | 0.86 | 0.81 | -0.25 | 0.01 | 0.00 | -2.93 | 0.61 | 0.00 | -0.01 | 0.03 | 0.78 |
| **Meta_MID** | MID | rs6235 | -0.14 | 0.46 | 0.77 | -0.25 | 0.01 | 0.00 | -2.92 | 0.61 | 0.00 | -0.01 | 0.03 | 0.79 |
| **Meta_MID** | MID | Baseline | | | | -0.25 | 0.01 | 0.00 | -2.92 | 0.61 | 0.00 | -0.01 | 0.03 | 0.78 |
| **Meta_analysis** | ALL | BMI PGS | 0.05 | 0.09 | 0.59 | -0.06 | 0.00 | 0.00 | -1.60 | 0.19 | 0.00 | -0.02 | 0.01 | 0.03 |
| **Meta_analysis** | ALL | T2D PGS | 0.11 | 0.10 | 0.26 | -0.05 | 0.00 | 0.00 | -1.60 | 0.19 | 0.00 | -0.02 | 0.01 | 0.03 |
| **Meta_analysis** | ALL | rs10305492 | 0.21 | 0.75 | 0.78 | -0.03 | 0.01 | 0.00 | -1.62 | 0.25 | 0.00 | -0.03 | 0.01 | 0.00 |
| **Meta_analysis** | ALL | rs146868158 | 3.48 | 3.80 | 0.36 | -0.04 | 0.02 | 0.03 | -1.90 | 0.87 | 0.03 | -0.02 | 0.03 | 0.47 |
| **Meta_analysis** | ALL | rs6923761 | -0.21 | 0.16 | 0.18 | -0.06 | 0.00 | 0.00 | -1.60 | 0.19 | 0.00 | -0.02 | 0.01 | 0.03 |
| **Meta_analysis** | ALL | rs1042044 | 0.03 | 0.12 | 0.79 | -0.05 | 0.00 | 0.00 | -1.58 | 0.19 | 0.00 | -0.02 | 0.01 | 0.03 |
| **Meta_analysis** | ALL | rs10305420 | -0.01 | 0.16 | 0.93 | -0.07 | 0.00 | 0.00 | -1.70 | 0.22 | 0.00 | -0.02 | 0.01 | 0.03 |
| **Meta_analysis** | ALL | rs3765467 | 0.18 | 0.52 | 0.73 | -0.06 | 0.00 | 0.00 | -1.65 | 0.21 | 0.00 | -0.02 | 0.01 | 0.01 |
| **Meta_analysis** | ALL | rs10305421 | -0.06 | 0.91 | 0.95 | -0.03 | 0.01 | 0.00 | -1.75 | 0.25 | 0.00 | -0.03 | 0.01 | 0.01 |
| **Meta_analysis** | ALL | rs10305510 | 1.55 | 0.96 | 0.10 | -0.01 | 0.01 | 0.04 | -0.56 | 0.34 | 0.10 | -0.02 | 0.01 | 0.10 |
| **Meta_analysis** | ALL | rs429358 | -0.20 | 0.25 | 0.42 | -0.08 | 0.01 | 0.00 | -2.04 | 0.25 | 0.00 | -0.02 | 0.01 | 0.11 |
| **Meta_analysis** | ALL | rs6232 | 0.13 | 0.34 | 0.69 | -0.06 | 0.00 | 0.00 | -1.67 | 0.20 | 0.00 | -0.02 | 0.01 | 0.01 |
| **Meta_analysis** | ALL | rs6235 | 0.14 | 0.15 | 0.34 | -0.05 | 0.00 | 0.00 | -1.60 | 0.19 | 0.00 | -0.02 | 0.01 | 0.04 |
| **Meta_analysis** | ALL | Baseline | | | | -0.05 | 0.00 | 0.00 | -1.54 | 0.19 | 0.00 | -0.02 | 0.01 | 0.05 |

**Supplementary Table 6 | Meta analysis results for BS (primary model).**
Ancestry-specific and multi-ancestry meta-analyses of the BS analysis.

| | | | Main genetic exposure | | | Baseline body weight | | | Sex | | | Age at initiation | | |
|---|---|---|---|---|---|---|---|---|---|---|---|---|---|---|
| **Study** | **Ancestry** | **Model (main exposure)** | **Beta** | **Standard Error** | **P-value** | **Beta** | **Standard Error** | **P-value** | **Beta** | **Standard Error** | **P-value** | **Beta** | **Standard Error** | **P-value** |
| **Meta_AFR** | AFR | BMI PGS | 0.48 | 0.60 | 0.42 | -0.10 | 0.02 | 0.00 | -1.94 | 1.64 | 0.24 | 0.14 | 0.05 | 0.00 |
| **Meta_AFR** | AFR | T2D PGS | 1.18 | 0.72 | 0.10 | -0.10 | 0.02 | 0.00 | -1.94 | 1.63 | 0.23 | 0.15 | 0.05 | 0.00 |
| **Meta_AFR** | AFR | rs429358 | 1.41 | 1.15 | 0.22 | -0.11 | 0.03 | 0.00 | -1.28 | 2.31 | 0.58 | 0.11 | 0.06 | 0.09 |
| **Meta_AFR** | AFR | rs17702901 | 1.65 | 1.71 | 0.34 | -0.11 | 0.02 | 0.00 | -1.91 | 1.62 | 0.24 | 0.14 | 0.05 | 0.00 |
| **Meta_AFR** | AFR | rs728996 | -1.48 | 0.76 | 0.05 | -0.10 | 0.02 | 0.00 | -1.82 | 1.63 | 0.26 | 0.14 | 0.05 | 0.00 |
| **Meta_AFR** | AFR | Baseline | | | | -0.09 | 0.02 | 0.00 | -1.80 | 1.57 | 0.25 | 0.14 | 0.04 | 0.00 |
| **Meta_EUR** | EUR | BMI PGS | 0.40 | 0.22 | 0.07 | -0.11 | 0.01 | 0.00 | -2.73 | 0.53 | 0.00 | 0.15 | 0.02 | 0.00 |
| **Meta_EUR** | EUR | T2D PGS | 0.31 | 0.25 | 0.21 | -0.10 | 0.01 | 0.00 | -2.88 | 0.54 | 0.00 | 0.15 | 0.02 | 0.00 |
| **Meta_EUR** | EUR | rs429358 | 0.21 | 0.51 | 0.69 | -0.10 | 0.01 | 0.00 | -3.09 | 0.60 | 0.00 | 0.13 | 0.02 | 0.00 |
| **Meta_EUR** | EUR | rs17702901 | 0.67 | 0.86 | 0.44 | -0.10 | 0.01 | 0.00 | -3.17 | 0.59 | 0.00 | 0.13 | 0.02 | 0.00 |
| **Meta_EUR** | EUR | rs728996 | 0.38 | 0.31 | 0.22 | -0.10 | 0.01 | 0.00 | -2.94 | 0.54 | 0.00 | 0.15 | 0.02 | 0.00 |
| **Meta_EUR** | EUR | Baseline | | | | -0.11 | 0.01 | 0.00 | -2.71 | 0.52 | 0.00 | 0.15 | 0.02 | 0.00 |
| **Meta_AMR** | AMR | BMI PGS | 1.70 | 0.93 | 0.07 | -0.16 | 0.05 | 0.00 | -4.45 | 2.60 | 0.09 | 0.05 | 0.09 | 0.56 |
| **Meta_AMR** | AMR | T2D PGS | 0.76 | 0.94 | 0.42 | -0.16 | 0.05 | 0.00 | -5.00 | 2.61 | 0.06 | 0.07 | 0.09 | 0.45 |
| **Meta_AMR** | AMR | rs429358 | | | | | | | | | | | | |
| **Meta_AMR** | AMR | rs17702901 | 2.66 | 1.86 | 0.15 | -0.17 | 0.05 | 0.00 | -4.84 | 2.60 | 0.06 | 0.04 | 0.09 | 0.66 |
| **Meta_AMR** | AMR | rs728996 | -0.82 | 1.49 | 0.58 | -0.16 | 0.05 | 0.00 | -5.04 | 2.62 | 0.05 | 0.06 | 0.09 | 0.51 |
| **Meta_AMR** | AMR | Baseline | | | | -0.13 | 0.05 | 0.01 | -3.81 | 2.56 | 0.14 | 0.08 | 0.08 | 0.33 |
| **Meta_SAS** | SAS | BMI PGS | | | | | | | | | | | | |
| **Meta_SAS** | SAS | T2D PGS | | | | | | | | | | | | |
| **Meta_SAS** | SAS | rs429358 | | | | | | | | | | | | |
| **Meta_SAS** | SAS | rs17702901 | | | | | | | | | | | | |
| **Meta_SAS** | SAS | rs728996 | | | | | | | | | | | | |
| **Meta_SAS** | SAS | Baseline | | | | | | | | | | | | |
| **Meta_MID** | MID | BMI PGS | 1.63 | 0.41 | 0.00 | -0.28 | 0.02 | 0.00 | -2.90 | 0.91 | 0.00 | 0.15 | 0.04 | 0.00 |
| **Meta_MID** | MID | T2D PGS | 0.51 | 0.46 | 0.27 | -0.28 | 0.02 | 0.00 | -2.66 | 0.92 | 0.00 | 0.15 | 0.04 | 0.00 |
| **Meta_MID** | MID | rs429358 | -0.64 | 1.00 | 0.52 | -0.28 | 0.02 | 0.00 | -2.63 | 0.92 | 0.00 | 0.15 | 0.04 | 0.00 |
| **Meta_MID** | MID | rs17702901 | -1.22 | 1.57 | 0.44 | -0.28 | 0.02 | 0.00 | -2.70 | 0.92 | 0.00 | 0.14 | 0.04 | 0.00 |
| **Meta_MID** | MID | rs728996 | 0.24 | 0.56 | 0.67 | -0.28 | 0.02 | 0.00 | -2.66 | 0.92 | 0.00 | 0.15 | 0.04 | 0.00 |
| **Meta_MID** | MID | Baseline | | | | -0.28 | 0.02 | 0.00 | -2.67 | 0.92 | 0.00 | 0.15 | 0.04 | 0.00 |
| **Meta_analysis** | ALL | BMI PGS | 0.70 | 0.18 | 0.00 | -0.14 | 0.01 | 0.00 | -2.76 | 0.44 | 0.00 | 0.15 | 0.02 | 0.00 |
| **Meta_analysis** | ALL | T2D PGS | 0.45 | 0.21 | 0.03 | -0.13 | 0.01 | 0.00 | -2.82 | 0.44 | 0.00 | 0.15 | 0.02 | 0.00 |
| **Meta_analysis** | ALL | rs429358 | 0.22 | 0.42 | 0.61 | -0.14 | 0.01 | 0.00 | -2.87 | 0.49 | 0.00 | 0.13 | 0.02 | 0.00 |
| **Meta_analysis** | ALL | rs17702901 | 0.73 | 0.65 | 0.26 | -0.14 | 0.01 | 0.00 | -3.00 | 0.47 | 0.00 | 0.13 | 0.02 | 0.00 |
| **Meta_analysis** | ALL | rs728996 | 0.11 | 0.25 | 0.66 | -0.13 | 0.01 | 0.00 | -2.86 | 0.44 | 0.00 | 0.15 | 0.02 | 0.00 |
| **Meta_analysis** | ALL | Baseline | | | | -0.14 | 0.01 | 0.00 | -2.66 | 0.43 | 0.00 | 0.15 | 0.02 | 0.00 |

**Supplementary Table 7 | Heterogeneity statistics for GLP1-RA meta analysis (Model A).**
Heterogeneity across ancestries and studies for the tested genetic exposures. Cochran's Q (P-Value) and the I^2 statistics are reported in the table.

| Exposure | Q coefficient | P-value Q coefficient | I2 |
|---|---|---|---|
| BMI PGS | 7.098522782 | 0.2134155646 | 0.295628097 |
| T2D PGS | 5.087704484 | 0.4052707326 | 0.01723851768 |
| rs10305492 | 2.484500517 | 0.2887337607 | 0.1950092237 |
| rs146868158 | 0 | | |
| rs6923761 | 1.586471311 | 0.8112215842 | 0 |
| rs1042044 | 2.698569045 | 0.746337287 | 0 |
| rs10305420 | 1.009796784 | 0.9083068566 | 0 |
| rs3765467 | 1.628175585 | 0.897821255 | 0 |
| rs10305421 | 2.037812598 | 0.360989539 | 0.01855548335 |
| rs10305510 | 0.004196699736 | 0.9483476211 | 0 |
| rs429358 | 1.624767074 | 0.6537871815 | 0 |
| rs6232 | 4.288551001 | 0.3683609415 | 0.06728403153 |
| rs6235 | 3.050406612 | 0.6922146717 | 0 |

**Supplementary Table 8 | Heterogeneity statistics for BS meta analysis (Model A).**
Heterogeneity across ancestries and studies for the tested genetic exposures. Cochran's Q (P-Value) and the I^2 statistics are reported in the table.

| Exposure | Q coefficient | P-value Q coefficient | I2 |
|---|---|---|---|
| **BMI PGS** | 8.242450086 | 0.04125793808 | 0.6360305529 |
| **T2D PGS** | 1.434179294 | 0.6975428336 | 0 |
| **rs429358** | 1.81672726 | 0.4031834417 | 0 |
| **rs17702901** | 2.898567328 | 0.4075299326 | 0 |
| **rs728996** | 5.616315641 | 0.1318448264 | 0.4658419875 |

**Supplementary Table 9 | Meta analysis results for GLP1-RA sensitivity analysis 1.**
Ancestry-specific and multi-ancestry meta-analyses of the GLP1-RA sensitivity analysis using a less conservative outcome definition.

| | | | Main genetic exposure | | | Baseline body weight | | | Sex | | | Age at initiation | | |
|---|---|---|---|---|---|---|---|---|---|---|---|---|---|---|
| **Study** | **Ancestry** | **Model (main exposure)** | **Beta** | **Standard Error** | **P-value** | **Beta** | **Standard Error** | **P-value** | **Beta** | **Standard Error** | **P-value** | **Beta** | **Standard Error** | **P-value** |
| **Meta_AFR** | AFR | BMI PGS | -0.12 | 0.22 | 0.59 | -0.01 | 0.01 | 0.18 | -0.46 | 0.48 | 0.33 | -0.01 | 0.02 | 0.52 |
| **Meta_AFR** | AFR | T2D PGS | 0.30 | 0.24 | 0.21 | -0.01 | 0.01 | 0.17 | -0.41 | 0.48 | 0.40 | -0.01 | 0.02 | 0.62 |
| **Meta_AFR** | AFR | Baseline | | | | -0.01 | 0.01 | 0.40 | -0.07 | 0.46 | 0.87 | -0.01 | 0.02 | 0.44 |
| **Meta_AMR** | AMR | BMI PGS | 0.01 | 0.20 | 0.96 | -0.01 | 0.01 | 0.28 | -0.47 | 0.45 | 0.29 | -0.03 | 0.02 | 0.09 |
| **Meta_AMR** | AMR | T2D PGS | 0.06 | 0.21 | 0.76 | -0.01 | 0.01 | 0.30 | -0.47 | 0.45 | 0.30 | -0.03 | 0.02 | 0.11 |
| **Meta_AMR** | AMR | Baseline | | | | -0.01 | 0.01 | 0.47 | -0.40 | 0.44 | 0.36 | -0.02 | 0.02 | 0.16 |
| **Meta_EAS** | EAS | BMI PGS | 0.08 | 0.72 | 0.91 | 0.03 | 0.05 | 0.60 | -1.09 | 1.59 | 0.49 | 0.00 | 0.06 | 1.00 |
| **Meta_EAS** | EAS | T2D PGS | 1.16 | 0.67 | 0.08 | 0.02 | 0.05 | 0.70 | -1.40 | 1.54 | 0.37 | 0.00 | 0.06 | 0.96 |
| **Meta_EAS** | EAS | Baseline | | | | 0.02 | 0.04 | 0.70 | -1.47 | 1.39 | 0.29 | -0.02 | 0.06 | 0.76 |
| **Meta_EUR** | EUR | BMI PGS | 0.13 | 0.15 | 0.38 | -0.03 | 0.01 | 0.00 | -1.92 | 0.31 | 0.00 | -0.02 | 0.01 | 0.23 |
| **Meta_EUR** | EUR | T2D PGS | -0.09 | 0.15 | 0.55 | -0.03 | 0.01 | 0.00 | -1.90 | 0.31 | 0.00 | -1.90 | 0.31 | 0.00 |
| **Meta_EUR** | EUR | Baseline | | | | -0.04 | 0.01 | 0.00 | -1.81 | 0.32 | 0.00 | -0.03 | 0.01 | 0.05 |
| **Meta_SAS** | SAS | BMI PGS | 0.31 | 0.87 | 0.72 | -0.09 | 0.04 | 0.05 | -4.09 | 2.00 | 0.04 | -0.13 | 0.07 | 0.08 |
| **Meta_SAS** | SAS | T2D PGS | 0.79 | 0.81 | 0.33 | -0.08 | 0.04 | 0.05 | -4.10 | 1.98 | 0.04 | -0.12 | 0.07 | 0.10 |
| **Meta_SAS** | SAS | Baseline | | | | -0.06 | 0.04 | 0.11 | -3.51 | 1.45 | 0.02 | -0.08 | 0.06 | 0.15 |
| **Meta_MID** | MID | BMI PGS | 0.62 | 0.29 | 0.03 | -0.27 | 0.01 | 0.00 | 3.50 | 0.64 | 0.00 | 0.06 | 0.03 | 0.05 |
| **Meta_MID** | MID | T2D PGS | 0.26 | 0.34 | 0.44 | -0.27 | 0.01 | 0.00 | 3.47 | 0.63 | 0.00 | 0.06 | 0.03 | 0.05 |
| **Meta_MID** | MID | Baseline | | | | -0.27 | 0.01 | 0.00 | 3.48 | 0.64 | 0.00 | 0.05 | 0.03 | 0.10 |
| **Meta_analysis** | ALL | BMI PGS | 0.06 | 0.09 | 0.49 | -0.07 | 0.00 | 0.00 | -0.81 | 0.19 | 0.00 | -0.02 | 0.01 | 0.06 |
| **Meta_analysis** | ALL | T2D PGS | 0.04 | 0.10 | 0.66 | -0.07 | 0.00 | 0.00 | -0.79 | 0.19 | 0.00 | -0.02 | 0.01 | 0.13 |
| **Meta_analysis** | ALL | Baseline | | | | -0.06 | 0.00 | 0.00 | -0.58 | 0.19 | 0.00 | -0.02 | 0.01 | 0.02 |

**Supplementary Table 10 | Meta analysis results for BS sensitivity analysis 1.**
Ancestry-specific and multi-ancestry meta-analyses of the BS sensitivity analysis using a less conservative outcome definition.

| | | | Main genetic exposure | | | Baseline body weight | | | Sex | | | Age at initiation | | |
|---|---|---|---|---|---|---|---|---|---|---|---|---|---|---|
| **Study** | **Ancestry** | **Model (main exposure)** | **Beta** | **Standard Error** | **P-value** | **Beta** | **Standard Error** | **P-value** | **Beta** | **Standard Error** | **P-value** | **Beta** | **Standard Error** | **P-value** |
| **Meta_AFR** | AFR | BMI PGS | 0.55 | 0.61 | 0.37 | -0.10 | 0.02 | 0.00 | -0.12 | 1.69 | 0.94 | 0.12 | 0.05 | 0.01 |
| **Meta_AFR** | AFR | T2D PGS | 0.75 | 0.74 | 0.31 | -0.10 | 0.02 | 0.00 | -0.13 | 1.68 | 0.94 | 0.13 | 0.05 | 0.01 |
| **Meta_AFR** | AFR | Baseline | | | | -0.09 | 0.02 | 0.00 | -0.10 | 1.59 | 0.95 | 0.12 | 0.05 | 0.01 |
| **Meta_AMR** | AMR | BMI PGS | 1.90 | 0.86 | 0.03 | -0.17 | 0.05 | 0.00 | -5.33 | 2.40 | 0.03 | 0.04 | 0.09 | 0.65 |
| **Meta_AMR** | AMR | T2D PGS | 0.67 | 0.88 | 0.45 | -0.17 | 0.05 | 0.00 | -5.94 | 2.43 | 0.01 | 0.06 | 0.09 | 0.53 |
| **Meta_AMR** | AMR | Baseline | | | | -0.15 | 0.05 | 0.00 | -5.07 | 2.38 | 0.03 | 0.08 | 0.08 | 0.32 |
| **Meta_EUR** | EUR | BMI PGS | 0.37 | 0.23 | 0.11 | -0.11 | 0.01 | 0.00 | -2.99 | 0.56 | 0.00 | 0.15 | 0.02 | 0.00 |
| **Meta_EUR** | EUR | T2D PGS | 0.29 | 0.26 | 0.27 | -0.11 | 0.01 | 0.00 | -2.95 | 0.56 | 0.00 | 0.15 | 0.02 | 0.00 |
| **Meta_EUR** | EUR | Baseline | | | | -0.14 | 0.01 | 0.00 | -3.43 | 0.70 | 0.00 | 0.16 | 0.03 | 0.00 |
| **Meta_SAS** | SAS | BMI PGS | | | | | | | | | | | | |
| **Meta_SAS** | SAS | T2D PGS | | | | | | | | | | | | |
| **Meta_SAS** | SAS | Baseline | | | | | | | | | | | | |
| **Meta_MID** | MID | BMI PGS | 0.95 | 0.43 | 0.03 | -0.31 | 0.02 | 0.00 | 5.03 | 0.97 | 0.00 | 0.15 | 0.04 | 0.00 |
| **Meta_MID** | MID | T2D PGS | 0.32 | 0.48 | 0.50 | -0.31 | 0.02 | 0.00 | 4.89 | 0.97 | 0.00 | 0.15 | 0.04 | 0.00 |
| **Meta_MID** | MID | Baseline | | | | -0.31 | 0.02 | 0.00 | 4.89 | 0.97 | 0.00 | 0.15 | 0.04 | 0.00 |
| **Meta_analysis** | ALL | BMI PGS | 0.39 | 0.16 | 0.02 | -0.14 | 0.01 | 0.00 | -1.47 | 0.39 | 0.00 | 0.14 | 0.01 | 0.00 |
| **Meta_analysis** | ALL | T2D PGS | 0.32 | 0.20 | 0.10 | -0.14 | 0.01 | 0.00 | -1.49 | 0.39 | 0.00 | 0.14 | 0.01 | 0.00 |
| **Meta_analysis** | ALL | Baseline | | | | -0.16 | 0.01 | 0.00 | -1.32 | 0.42 | 0.00 | 0.14 | 0.01 | 0.00 |

**Supplementary Table 11 | Meta analysis results for BS sensitivity analysis 2.**
Ancestry-specific and multi-ancestry meta-analyses of the BS sensitivity analysis: "shorter follow-up".

| | | | Main genetic exposure | | | Baseline body weight | | | Sex | | | Age at initiation | | |
|---|---|---|---|---|---|---|---|---|---|---|---|---|---|---|
| **Study** | **Ancestry** | **Model (main exposure)** | **Beta** | **Standard Error** | **P-value** | **Beta** | **Standard Error** | **P-value** | **Beta** | **Standard Error** | **P-value** | **Beta** | **Standard Error** | **P-value** |
| **Meta_AFR** | AFR | BMI PGS | 0.87 | 0.58 | 0.13 | -0.08 | 0.02 | 0.00 | 0.40 | 1.60 | 0.80 | 0.12 | 0.04 | 0.00 |
| **Meta_AFR** | AFR | T2D PGS | 1.17 | 0.71 | 0.10 | -0.08 | 0.02 | 0.00 | 0.50 | 1.60 | 0.75 | 0.13 | 0.04 | 0.00 |
| **Meta_AFR** | AFR | Baseline | | | | -0.07 | 0.02 | 0.00 | 0.90 | 1.51 | 0.55 | 0.12 | 0.04 | 0.00 |
| **Meta_AMR** | AMR | BMI PGS | 1.45 | 0.89 | 0.10 | -0.08 | 0.05 | 0.08 | -2.13 | 2.39 | 0.37 | 0.04 | 0.09 | 0.64 |
| **Meta_AMR** | AMR | T2D PGS | 0.19 | 0.91 | 0.83 | -0.09 | 0.05 | 0.07 | -2.67 | 2.41 | 0.27 | 0.06 | 0.09 | 0.52 |
| **Meta_AMR** | AMR | Baseline | | | | -0.09 | 0.05 | 0.08 | -2.94 | 2.52 | 0.24 | 0.14 | 0.08 | 0.10 |
| **Meta_EUR** | EUR | BMI PGS | 0.07 | 0.25 | 0.78 | -0.11 | 0.01 | 0.00 | -2.35 | 0.62 | 0.00 | 0.15 | 0.02 | 0.00 |
| **Meta_EUR** | EUR | T2D PGS | -0.02 | 0.32 | 0.96 | -0.11 | 0.01 | 0.00 | -2.45 | 0.62 | 0.00 | 0.15 | 0.02 | 0.00 |
| **Meta_EUR** | EUR | Baseline | | | | -0.11 | 0.01 | 0.00 | -2.29 | 0.65 | 0.00 | 0.15 | 0.02 | 0.00 |
| **Meta_SAS** | SAS | BMI PGS | | | | | | | | | | | | |
| **Meta_SAS** | SAS | T2D PGS | | | | | | | | | | | | |
| **Meta_SAS** | SAS | Baseline | | | | | | | | | | | | |
| **Meta_MID** | MID | BMI PGS | 0.81 | 0.37 | 0.03 | -0.21 | 0.02 | 0.00 | 4.43 | 0.84 | 0.00 | 0.08 | 0.03 | 0.01 |
| **Meta_MID** | MID | T2D PGS | 0.10 | 0.42 | 0.81 | -0.21 | 0.02 | 0.00 | 4.31 | 0.84 | 0.00 | 0.08 | 0.03 | 0.01 |
| **Meta_MID** | MID | Baseline | | | | -0.21 | 0.02 | 0.00 | 4.31 | 0.84 | 0.00 | 0.08 | 0.03 | 0.01 |
| **Meta_analysis** | ALL | BMI PGS | 0.43 | 0.19 | 0.03 | -0.12 | 0.01 | 0.00 | 0.00 | 0.47 | 1.00 | 0.12 | 0.02 | 0.00 |
| **Meta_analysis** | ALL | T2D PGS | 0.16 | 0.23 | 0.49 | -0.12 | 0.01 | 0.00 | -0.10 | 0.47 | 0.83 | 0.12 | 0.02 | 0.00 |
| **Meta_analysis** | ALL | Baseline | | | | -0.12 | 0.01 | 0.00 | 0.13 | 0.48 | 0.79 | 0.12 | 0.02 | 0.00 |

**Supplementary Table 12 | Meta analysis results for GLP1-RA sensitivity analysis 3.**
Ancestry-specific and multi-ancestry meta-analyses of the GLP1-RA sensitivity analysis: "adjustment for T2D".

| | | | Main genetic exposure | | | Baseline body weight | | | Sex | | | Age at initiation | | | T2D status | | |
|---|---|---|---|---|---|---|---|---|---|---|---|---|---|---|---|---|---|
| **Study** | **Ancestry** | **Model (main exposure)** | **Beta** | **Standard Error** | **P-value** | **Beta** | **Standard Error** | **P-value** | **Beta** | **Standard Error** | **P-value** | **Beta** | **Standard Error** | **P-value** | **Beta** | **Standard Error** | **P-value** |
| **Meta_AFR** | AFR | BMI PGS | -0.13 | 0.21 | 0.54 | -0.01 | 0.01 | 0.08 | -0.29 | 0.45 | 0.53 | -0.01 | 0.02 | 0.39 | 0.18 | 0.50 | 0.73 |
| **Meta_AFR** | AFR | T2D PGS | 0.32 | 0.22 | 0.16 | -0.02 | 0.01 | 0.07 | -0.23 | 0.45 | 0.61 | -0.01 | 0.02 | 0.48 | 0.24 | 0.51 | 0.64 |
| **Meta_AFR** | AFR | Baseline | | | | -0.01 | 0.01 | 0.17 | -0.07 | 0.44 | 0.88 | -0.02 | 0.02 | 0.37 | 0.39 | 0.48 | 0.41 |
| **Meta_AMR** | AMR | BMI PGS | -0.08 | 0.20 | 0.67 | -0.01 | 0.01 | 0.21 | -0.56 | 0.43 | 0.19 | -0.03 | 0.02 | 0.10 | -0.09 | 0.51 | 0.86 |
| **Meta_AMR** | AMR | T2D PGS | 0.01 | 0.20 | 0.95 | -0.01 | 0.01 | 0.19 | -0.57 | 0.43 | 0.19 | -0.03 | 0.02 | 0.11 | -0.05 | 0.51 | 0.92 |
| **Meta_AMR** | AMR | Baseline | | | | -0.01 | 0.01 | 0.33 | -0.53 | 0.42 | 0.22 | -0.02 | 0.02 | 0.19 | 0.23 | 0.47 | 0.62 |
| **Meta_EAS** | EAS | BMI PGS | 0.08 | 0.71 | 0.91 | 0.02 | 0.05 | 0.71 | -0.76 | 1.59 | 0.63 | -0.02 | 0.06 | 0.73 | 4.48 | 3.04 | 0.14 |
| **Meta_EAS** | EAS | T2D PGS | 1.08 | 0.67 | 0.11 | 0.01 | 0.05 | 0.80 | -1.07 | 1.55 | 0.49 | -0.02 | 0.06 | 0.78 | 4.01 | 3.00 | 0.18 |
| **Meta_EAS** | EAS | Baseline | | | | 0.01 | 0.04 | 0.77 | -1.04 | 1.40 | 0.46 | -0.04 | 0.06 | 0.52 | 4.38 | 2.74 | 0.11 |
| **Meta_EUR** | EUR | BMI PGS | -0.04 | 0.15 | 0.78 | -0.03 | 0.01 | 0.00 | -1.53 | 0.31 | 0.00 | -0.03 | 0.01 | 0.03 | 0.07 | 0.39 | 0.85 |
| **Meta_EUR** | EUR | T2D PGS | -0.06 | 0.15 | 0.69 | -0.03 | 0.01 | 0.00 | -1.54 | 0.31 | 0.00 | -0.03 | 0.01 | 0.03 | -0.02 | 0.39 | 0.96 |
| **Meta_EUR** | EUR | Baseline | | | | -0.03 | 0.01 | 0.00 | -1.55 | 0.31 | 0.00 | -0.03 | 0.01 | 0.01 | 0.33 | 0.38 | 0.39 |
| **Meta_SAS** | SAS | BMI PGS | 0.04 | 0.91 | 0.96 | -0.09 | 0.05 | 0.05 | -3.64 | 2.15 | 0.09 | -0.14 | 0.07 | 0.06 | -2.08 | 2.14 | 0.33 |
| **Meta_SAS** | SAS | T2D PGS | 0.83 | 0.88 | 0.34 | -0.09 | 0.04 | 0.04 | -3.82 | 2.14 | 0.07 | -0.12 | 0.07 | 0.09 | -1.39 | 2.23 | 0.53 |
| **Meta_SAS** | SAS | Baseline | | | | -0.08 | 0.04 | 0.03 | -3.68 | 1.49 | 0.01 | -0.12 | 0.06 | 0.05 | -1.63 | 1.55 | 0.29 |
| **Meta_MID** | MID | BMI PGS | 0.82 | 0.28 | 0.00 | -0.25 | 0.01 | 0.00 | 3.03 | 0.61 | 0.00 | -0.03 | 0.03 | 0.32 | 1.82 | 0.70 | 0.01 |
| **Meta_MID** | MID | T2D PGS | -0.06 | 0.33 | 0.86 | -0.26 | 0.01 | 0.00 | 2.99 | 0.61 | 0.00 | -0.04 | 0.03 | 0.18 | 1.85 | 0.71 | 0.01 |
| **Meta_MID** | MID | Baseline | | | | -0.26 | 0.01 | 0.00 | 2.99 | 0.61 | 0.00 | -0.04 | 0.03 | 0.18 | 1.83 | 0.71 | 0.01 |
| **Meta_analysis** | ALL | BMI PGS | -0.02 | 0.09 | 0.79 | -0.06 | 0.00 | 0.00 | -0.51 | 0.19 | 0.01 | -0.03 | 0.01 | 0.00 | 0.35 | 0.24 | 0.14 |
| **Meta_analysis** | ALL | T2D PGS | 0.05 | 0.10 | 0.62 | -0.06 | 0.00 | 0.00 | -0.53 | 0.19 | 0.00 | -0.03 | 0.01 | 0.00 | 0.35 | 0.24 | 0.15 |
| **Meta_analysis** | ALL | Baseline | | | | -0.06 | 0.00 | 0.00 | -0.50 | 0.19 | 0.01 | -0.03 | 0.01 | 0.00 | 0.54 | 0.23 | 0.02 |

**Supplementary Table 13 | Meta analysis results for BS sensitivity analysis 3.**
Ancestry-specific and multi-ancestry meta-analyses of the BS sensitivity analysis: "adjustment for T2D".

| | | | Main genetic exposure | | | Baseline body weight | | | Sex | | | Age at initiation | | | T2D status | | |
|---|---|---|---|---|---|---|---|---|---|---|---|---|---|---|---|---|---|
| **Study** | **Ancestry** | **Model (main exposure)** | **Beta** | **Standard Error** | **P-value** | **Beta** | **Standard Error** | **P-value** | **Beta** | **Standard Error** | **P-value** | **Beta** | **Standard Error** | **P-value** | **Beta** | **Standard Error** | **P-value** |
| **Meta_AFR** | AFR | BMI PGS | 0.74 | 0.63 | 0.24 | -0.11 | 0.02 | 0.00 | 0.69 | 1.67 | 0.68 | 0.11 | 0.05 | 0.02 | -4.19 | 1.32 | 0.00 |
| **Meta_AFR** | AFR | T2D PGS | 1.22 | 0.74 | 0.10 | -0.11 | 0.02 | 0.00 | 0.75 | 1.65 | 0.65 | 0.12 | 0.05 | 0.02 | -4.08 | 1.32 | 0.00 |
| **Meta_AFR** | AFR | Baseline | | | | -0.10 | 0.02 | 0.00 | 0.59 | 1.58 | 0.71 | 0.10 | 0.05 | 0.03 | -4.53 | 1.27 | 0.00 |
| **Meta_AMR** | AMR | BMI PGS | 1.61 | 0.92 | 0.08 | -0.15 | 0.05 | 0.00 | -3.78 | 2.62 | 0.15 | 0.03 | 0.09 | 0.79 | -3.28 | 2.11 | 0.12 |
| **Meta_AMR** | AMR | T2D PGS | 0.50 | 0.96 | 0.60 | -0.15 | 0.05 | 0.00 | -4.29 | 2.64 | 0.10 | 0.04 | 0.10 | 0.69 | -3.30 | 2.17 | 0.13 |
| **Meta_AMR** | AMR | Baseline | | | | -0.13 | 0.05 | 0.01 | -3.38 | 2.60 | 0.19 | 0.06 | 0.09 | 0.49 | -2.21 | 2.15 | 0.30 |
| **Meta_EUR** | EUR | BMI PGS | 0.58 | 0.25 | 0.02 | -0.08 | 0.01 | 0.00 | -1.79 | 0.60 | 0.00 | 0.16 | 0.03 | 0.00 | -0.07 | 0.56 | 0.90 |
| **Meta_EUR** | EUR | T2D PGS | 0.30 | 0.27 | 0.26 | -0.07 | 0.01 | 0.00 | -1.76 | 0.60 | 0.00 | 0.16 | 0.03 | 0.00 | 0.03 | 0.57 | 0.95 |
| **Meta_EUR** | EUR | Baseline | | | | -0.12 | 0.02 | 0.00 | -1.85 | 0.85 | 0.03 | 0.18 | 0.03 | 0.00 | 0.65 | 0.83 | 0.43 |
| **Meta_SAS** | SAS | BMI PGS | | | | | | | | | | | | | | | |
| **Meta_SAS** | SAS | T2D PGS | | | | | | | | | | | | | | | |
| **Meta_SAS** | SAS | Baseline | | | | | | | | | | | | | | | |
| **Meta_MID** | MID | BMI PGS | 0.78 | 0.37 | 0.04 | -0.21 | 0.02 | 0.00 | 4.40 | 0.84 | 0.00 | 0.06 | 0.04 | 0.13 | 1.16 | 0.79 | 0.14 |
| **Meta_MID** | MID | T2D PGS | -0.01 | 0.42 | 0.98 | -0.21 | 0.02 | 0.00 | 4.29 | 0.84 | 0.00 | 0.06 | 0.04 | 0.13 | 1.26 | 0.80 | 0.12 |
| **Meta_MID** | MID | Baseline | | | | -0.21 | 0.02 | 0.00 | 4.29 | 0.84 | 0.00 | 0.06 | 0.04 | 0.13 | 1.25 | 0.79 | 0.11 |
| **Meta_analysis** | ALL | BMI PGS | 0.59 | 0.17 | 0.00 | -0.12 | 0.01 | 0.00 | 0.17 | 0.41 | 0.68 | 0.12 | 0.02 | 0.00 | -0.39 | 0.39 | 0.32 |
| **Meta_analysis** | ALL | T2D PGS | 0.31 | 0.20 | 0.13 | -0.12 | 0.01 | 0.00 | 0.15 | 0.41 | 0.72 | 0.12 | 0.02 | 0.00 | -0.36 | 0.40 | 0.36 |
| **Meta_analysis** | ALL | Baseline | | | | -0.14 | 0.01 | 0.00 | 0.72 | 0.47 | 0.12 | 0.11 | 0.02 | 0.00 | -0.28 | 0.45 | 0.53 |

**Supplementary Table 14 | Meta analysis results for GLP1-RA sensitivity analysis 4.**
Ancestry-specific and multi-ancestry meta-analyses of the GLP1-RA sensitivity analysis: "including only liraglutide and semaglutide".

| | | | Main genetic exposure | | | Baseline body weight | | | Sex | | | Age at initiation | | |
|---|---|---|---|---|---|---|---|---|---|---|---|---|---|---|
| **Study** | **Ancestry** | **Model (main exposure)** | **Beta** | **Standard Error** | **P-value** | **Beta** | **Standard Error** | **P-value** | **Beta** | **Standard Error** | **P-value** | **Beta** | **Standard Error** | **P-value** |
| **Meta_AFR** | AFR | BMI PGS | -0.12 | 0.29 | 0.68 | -0.02 | 0.01 | 0.08 | -0.63 | 0.63 | 0.32 | 0.01 | 0.02 | 0.61 |
| **Meta_AFR** | AFR | T2D PGS | 0.47 | 0.30 | 0.12 | -0.02 | 0.01 | 0.09 | -0.54 | 0.63 | 0.39 | 0.02 | 0.02 | 0.47 |
| **Meta_AFR** | AFR | Baseline | | | | -0.02 | 0.01 | 0.12 | -0.48 | 0.60 | 0.43 | 0.01 | 0.02 | 0.72 |
| **Meta_AMR** | AMR | BMI PGS | 0.07 | 0.26 | 0.79 | -0.02 | 0.01 | 0.12 | -0.90 | 0.59 | 0.13 | -0.01 | 0.02 | 0.67 |
| **Meta_AMR** | AMR | T2D PGS | 0.32 | 0.27 | 0.23 | -0.02 | 0.01 | 0.17 | -0.89 | 0.59 | 0.13 | 0.00 | 0.02 | 0.87 |
| **Meta_AMR** | AMR | Baseline | | | | -0.01 | 0.01 | 0.25 | -0.75 | 0.57 | 0.18 | 0.00 | 0.02 | 0.91 |
| **Meta_EAS** | EAS | BMI PGS | 0.63 | 0.63 | 0.32 | 0.06 | 0.04 | 0.18 | -1.61 | 1.50 | 0.28 | -0.01 | 0.06 | 0.90 |
| **Meta_EAS** | EAS | T2D PGS | 0.53 | 0.60 | 0.38 | 0.05 | 0.04 | 0.22 | -1.54 | 1.50 | 0.31 | -0.01 | 0.06 | 0.84 |
| **Meta_EAS** | EAS | Baseline | | | | 0.05 | 0.04 | 0.20 | -2.74 | 1.26 | 0.03 | -0.02 | 0.05 | 0.75 |
| **Meta_EUR** | EUR | BMI PGS | 0.08 | 0.17 | 0.62 | -0.03 | 0.01 | 0.00 | -2.41 | 0.34 | 0.00 | 0.00 | 0.01 | 0.76 |
| **Meta_EUR** | EUR | T2D PGS | -0.03 | 0.18 | 0.88 | -0.03 | 0.01 | 0.00 | -2.35 | 0.34 | 0.00 | 0.00 | 0.01 | 0.76 |
| **Meta_EUR** | EUR | Baseline | | | | -0.03 | 0.01 | 0.00 | -2.46 | 0.35 | 0.00 | -0.02 | 0.01 | 0.24 |
| **Meta_SAS** | SAS | BMI PGS | -1.42 | 1.40 | 0.31 | -0.18 | 0.08 | 0.02 | -6.34 | 3.16 | 0.04 | -0.15 | 0.11 | 0.17 |
| **Meta_SAS** | SAS | T2D PGS | 1.66 | 1.26 | 0.19 | -0.18 | 0.08 | 0.01 | -4.86 | 3.18 | 0.13 | -0.07 | 0.12 | 0.55 |
| **Meta_SAS** | SAS | Baseline | | | | -0.11 | 0.05 | 0.02 | -5.26 | 2.07 | 0.01 | -0.11 | 0.08 | 0.13 |
| **Meta_MID** | MID | BMI PGS | 0.89 | 0.36 | 0.01 | -0.22 | 0.01 | 0.00 | 3.68 | 0.80 | 0.00 | 0.06 | 0.03 | 0.05 |
| **Meta_MID** | MID | T2D PGS | 0.02 | 0.41 | 0.96 | -0.22 | 0.01 | 0.00 | 3.59 | 0.80 | 0.00 | 0.06 | 0.03 | 0.05 |
| **Meta_MID** | MID | Baseline | | | | -0.23 | 0.01 | 0.00 | 3.59 | 0.80 | 0.00 | 0.06 | 0.03 | 0.05 |
| **Meta_analysis** | ALL | BMI PGS | 0.10 | 0.11 | 0.36 | -0.07 | 0.00 | 0.00 | -1.27 | 0.23 | 0.00 | 0.00 | 0.01 | 0.92 |
| **Meta_analysis** | ALL | T2D PGS | 0.10 | 0.12 | 0.39 | -0.07 | 0.00 | 0.00 | -1.24 | 0.23 | 0.00 | 0.00 | 0.01 | 0.88 |
| **Meta_analysis** | ALL | Baseline | | | | -0.07 | 0.00 | 0.00 | -1.28 | 0.23 | 0.00 | -0.01 | 0.01 | 0.58 |

**Supplementary Table 15 | Meta analysis results for BS sensitivity analysis 5.**
Ancestry-specific and multi-ancestry meta-analyses of the BS sensitivity analysis: "stricter inclusion criteria".

| | | | Main genetic exposure | | | Baseline body weight | | | Sex | | | Age at initiation | | |
|---|---|---|---|---|---|---|---|---|---|---|---|---|---|---|
| **Study** | **Ancestry** | **Model (main exposure)** | **Beta** | **Standard Error** | **P-value** | **Beta** | **Standard Error** | **P-value** | **Beta** | **Standard Error** | **P-value** | **Beta** | **Standard Error** | **P-value** |
| **Meta_AFR** | AFR | BMI PGS | 0.50 | 0.61 | 0.42 | -0.11 | 0.02 | 0.00 | 0.07 | 1.68 | 0.97 | 0.14 | 0.05 | 0.00 |
| **Meta_AFR** | AFR | T2D PGS | 1.49 | 0.73 | 0.04 | -0.11 | 0.02 | 0.00 | 0.04 | 1.66 | 0.98 | 0.16 | 0.05 | 0.00 |
| **Meta_AFR** | AFR | Baseline | | | | -0.10 | 0.02 | 0.00 | 0.08 | 1.59 | 0.96 | 0.14 | 0.05 | 0.00 |
| **Meta_AMR** | AMR | BMI PGS | 1.90 | 0.94 | 0.04 | -0.15 | 0.05 | 0.00 | -4.46 | 2.62 | 0.09 | 0.05 | 0.09 | 0.62 |
| **Meta_AMR** | AMR | T2D PGS | 0.88 | 0.96 | 0.36 | -0.16 | 0.05 | 0.00 | -5.09 | 2.64 | 0.05 | 0.06 | 0.09 | 0.50 |
| **Meta_AMR** | AMR | Baseline | | | | -0.13 | 0.05 | 0.01 | -4.09 | 2.60 | 0.12 | 0.06 | 0.08 | 0.50 |
| **Meta_EUR** | EUR | BMI PGS | 0.71 | 0.29 | 0.02 | -0.16 | 0.01 | 0.00 | -3.81 | 0.74 | 0.00 | 0.14 | 0.03 | 0.00 |
| **Meta_EUR** | EUR | T2D PGS | 0.52 | 0.35 | 0.14 | -0.18 | 0.01 | 0.00 | -3.89 | 0.74 | 0.00 | 0.14 | 0.03 | 0.00 |
| **Meta_EUR** | EUR | Baseline | | | | -0.16 | 0.01 | 0.00 | -3.55 | 0.79 | 0.00 | 0.15 | 0.03 | 0.00 |
| **Meta_SAS** | SAS | BMI PGS | | | | | | | | | | | | |
| **Meta_SAS** | SAS | T2D PGS | | | | | | | | | | | | |
| **Meta_SAS** | SAS | Baseline | | | | | | | | | | | | |
| **Meta_MID** | MID | BMI PGS | 0.89 | 0.38 | 0.02 | -0.28 | 0.01 | 0.00 | 7.40 | 0.84 | 0.00 | 0.05 | 0.03 | 0.10 |
| **Meta_MID** | MID | T2D PGS | -0.07 | 0.42 | 0.87 | -0.28 | 0.01 | 0.00 | 7.23 | 0.83 | 0.00 | 0.05 | 0.03 | 0.10 |
| **Meta_MID** | MID | Baseline | | | | -0.28 | 0.01 | 0.00 | 7.23 | 0.84 | 0.00 | 0.05 | 0.03 | 0.10 |
| **Meta_analysis** | ALL | BMI PGS | 0.66 | 0.18 | 0.00 | -0.21 | 0.01 | 0.00 | -0.19 | 0.44 | 0.67 | 0.11 | 0.02 | 0.00 |
| **Meta_analysis** | ALL | T2D PGS | 0.45 | 0.23 | 0.05 | -0.21 | 0.01 | 0.00 | -0.22 | 0.44 | 0.62 | 0.11 | 0.02 | 0.00 |
| **Meta_analysis** | ALL | Baseline | | | | -0.21 | 0.01 | 0.00 | 0.04 | 0.45 | 0.93 | 0.11 | 0.02 | 0.00 |

**Supplementary Table 16 | Meta analysis results for GLP1-RA sensitivity analysis 6.**
Ancestry-specific and multi-ancestry meta-analyses of the GLP1-RA sensitivity analysis: "Model using post-treatment weight as outcome and including interaction between baseline weight and each genetic exposure"

| | | | Interaction between main genetic exposure and baseline body weight | | | Main genetic exposure | | | Baseline body weight | | | Sex | | | Age at initiation | | |
|---|---|---|---|---|---|---|---|---|---|---|---|---|---|---|---|---|---|
| **Study** | **Ancestry** | **Model (main exposure)** | **Beta** | **Standard Error** | **P-value** | **Beta** | **Standard Error** | **P-value** | **Beta** | **Standard Error** | **P-value** | **Beta** | **Standard Error** | **P-value** | **Beta** | **Standard Error** | **P-value** |
| **Meta_AFR** | AFR | BMI PGS | 0.03 | 0.01 | 0.05 | 0.02 | 0.01 | 0.10 | -0.04 | 0.02 | 0.04 | 0.96 | 0.01 | 0.00 | -0.02 | 0.03 | 0.60 |
| **Meta_AFR** | AFR | T2D PGS | 0.01 | 0.01 | 0.32 | -0.59 | 0.98 | 0.55 | 0.83 | 0.01 | 0.00 | -0.34 | 0.48 | 0.47 | -0.02 | 0.02 | 0.38 |
| **Meta_AFR** | AFR | rs10305492 | 2.54 | 2.06 | 0.22 | -265.28 | 208.99 | 0.20 | 0.00 | 0.03 | 0.85 | -0.59 | 1.42 | 0.68 | -0.13 | 0.05 | 0.02 |
| **Meta_AFR** | AFR | rs146868158 | | | | | | | | | | | | | | | |
| **Meta_AFR** | AFR | rs6923761 | 0.00 | 0.02 | 0.97 | -0.50 | 2.47 | 0.84 | 0.67 | 0.01 | 0.00 | -0.42 | 0.48 | 0.38 | -0.02 | 0.02 | 0.34 |
| **Meta_AFR** | AFR | rs1042044 | -0.02 | 0.01 | 0.23 | 1.59 | 1.32 | 0.23 | 0.66 | 0.02 | 0.00 | -0.44 | 0.48 | 0.36 | -0.02 | 0.02 | 0.35 |
| **Meta_AFR** | AFR | rs10305420 | 0.00 | 0.02 | 0.99 | -0.18 | 2.43 | 0.94 | 0.74 | 0.01 | 0.00 | -0.80 | 0.51 | 0.12 | -0.03 | 0.02 | 0.12 |
| **Meta_AFR** | AFR | rs3765467 | 0.14 | 0.14 | 0.33 | -13.51 | 14.21 | 0.34 | 0.96 | 0.01 | 0.00 | -0.74 | 0.57 | 0.19 | -0.02 | 0.02 | 0.39 |
| **Meta_AFR** | AFR | rs10305421 | 0.31 | 0.84 | 0.71 | -43.06 | 98.17 | 0.66 | 0.00 | 0.03 | 0.93 | -0.38 | 1.42 | 0.79 | -0.12 | 0.06 | 0.04 |
| **Meta_AFR** | AFR | rs10305510 | 0.00 | 0.05 | 0.97 | 2.39 | 5.01 | 0.63 | 0.73 | 0.01 | 0.00 | -0.73 | 0.51 | 0.15 | -0.03 | 0.02 | 0.12 |
| **Meta_AFR** | AFR | rs429358 | 0.00 | 0.03 | 0.88 | 0.24 | 3.13 | 0.94 | -0.02 | 0.02 | 0.44 | 0.20 | 0.89 | 0.83 | -0.02 | 0.03 | 0.57 |
| **Meta_AFR** | AFR | rs6232 | -0.05 | 0.08 | 0.51 | 7.38 | 8.39 | 0.38 | 0.82 | 0.01 | 0.00 | -0.73 | 0.53 | 0.17 | -0.03 | 0.02 | 0.10 |
| **Meta_AFR** | AFR | rs6235 | 0.04 | 0.02 | 0.01 | -3.74 | 1.73 | 0.03 | 0.64 | 0.01 | 0.00 | -0.38 | 0.48 | 0.42 | -0.01 | 0.02 | 0.42 |
| **Meta_AFR** | AFR | Baseline | | | | | | | 0.96 | 0.01 | 0.00 | -0.31 | 0.46 | 0.50 | -0.02 | 0.02 | 0.31 |
| **Meta_AMR** | AMR | BMI PGS | 0.02 | 0.01 | 0.02 | -1.83 | 0.76 | 0.02 | 0.69 | 0.01 | 0.00 | -0.53 | 0.40 | 0.18 | -0.03 | 0.02 | 0.10 |
| **Meta_AMR** | AMR | T2D PGS | 0.01 | 0.01 | 0.38 | -0.62 | 0.77 | 0.42 | 0.72 | 0.01 | 0.00 | -0.53 | 0.40 | 0.18 | -0.02 | 0.02 | 0.18 |
| **Meta_AMR** | AMR | rs10305492 | 0.12 | 0.11 | 0.28 | -11.35 | 10.21 | 0.27 | 0.70 | 0.01 | 0.00 | -0.54 | 0.40 | 0.17 | -0.02 | 0.02 | 0.14 |
| **Meta_AMR** | AMR | rs146868158 | | | | | | | | | | | | | | | |
| **Meta_AMR** | AMR | rs6923761 | 0.03 | 0.02 | 0.03 | -3.14 | 1.48 | 0.03 | 0.66 | 0.01 | 0.00 | -0.52 | 0.40 | 0.19 | -0.03 | 0.02 | 0.13 |
| **Meta_AMR** | AMR | rs1042044 | 0.01 | 0.01 | 0.33 | -1.15 | 1.09 | 0.29 | 0.68 | 0.02 | 0.00 | -0.52 | 0.40 | 0.19 | -0.02 | 0.02 | 0.14 |
| **Meta_AMR** | AMR | rs10305420 | 0.02 | 0.02 | 0.24 | -1.41 | 1.43 | 0.32 | 0.96 | 0.01 | 0.00 | -0.47 | 0.47 | 0.32 | -0.03 | 0.02 | 0.16 |
| **Meta_AMR** | AMR | rs3765467 | 0.08 | 0.04 | 0.02 | -6.84 | 3.46 | 0.05 | 0.69 | 0.01 | 0.00 | -0.60 | 0.40 | 0.13 | -0.02 | 0.02 | 0.16 |
| **Meta_AMR** | AMR | rs10305421 | 0.03 | 0.10 | 0.79 | -2.73 | 8.27 | 0.74 | 0.97 | 0.01 | 0.00 | -0.45 | 0.47 | 0.33 | -0.03 | 0.02 | 0.13 |
| **Meta_AMR** | AMR | rs10305510 | -0.14 | 0.08 | 0.09 | 15.02 | 8.55 | 0.08 | 0.97 | 0.01 | 0.00 | -0.45 | 0.46 | 0.33 | -0.03 | 0.02 | 0.16 |
| **Meta_AMR** | AMR | rs429358 | 0.02 | 0.03 | 0.47 | -1.83 | 3.21 | 0.57 | -0.02 | 0.02 | 0.34 | -0.72 | 0.77 | 0.35 | -0.01 | 0.03 | 0.69 |
| **Meta_AMR** | AMR | rs6232 | -0.04 | 0.03 | 0.23 | 2.54 | 3.05 | 0.40 | 0.70 | 0.01 | 0.00 | -0.51 | 0.40 | 0.20 | -0.03 | 0.02 | 0.12 |
| **Meta_AMR** | AMR | rs6235 | 0.00 | 0.01 | 1.00 | 0.10 | 1.32 | 0.94 | 0.70 | 0.01 | 0.00 | -0.54 | 0.40 | 0.17 | -0.02 | 0.02 | 0.14 |
| **Meta_AMR** | AMR | Baseline | | | | | | | 0.97 | 0.01 | 0.00 | -0.40 | 0.38 | 0.29 | -0.02 | 0.02 | 0.31 |
| **Meta_EAS** | EAS | BMI PGS | 0.01 | 0.04 | 0.87 | -0.47 | 3.17 | 0.88 | 0.00 | 0.05 | 0.98 | -1.54 | 1.41 | 0.28 | -0.01 | 0.05 | 0.87 |
| **Meta_EAS** | EAS | T2D PGS | 0.02 | 0.05 | 0.74 | -0.48 | 3.78 | 0.90 | -0.01 | 0.05 | 0.86 | -1.64 | 1.39 | 0.24 | 0.00 | 0.05 | 0.96 |
| **Meta_EAS** | EAS | rs10305492 | | | | | | | | | | | | | | | |
| **Meta_EAS** | EAS | rs146868158 | | | | | | | | | | | | | | | |
| **Meta_EAS** | EAS | rs6923761 | | | | | | | | | | | | | | | |
| **Meta_EAS** | EAS | rs1042044 | -0.04 | 0.06 | 0.53 | 2.68 | 5.01 | 0.59 | 0.04 | 0.09 | 0.61 | -1.48 | 1.40 | 0.29 | -0.01 | 0.05 | 0.84 |
| **Meta_EAS** | EAS | rs10305420 | | | | | | | | | | | | | | | |
| **Meta_EAS** | EAS | rs3765467 | 0.08 | 0.08 | 0.36 | -6.56 | 6.63 | 0.32 | -0.02 | 0.05 | 0.72 | -1.23 | 1.43 | 0.39 | -0.01 | 0.05 | 0.85 |
| **Meta_EAS** | EAS | rs10305421 | | | | | | | | | | | | | | | |
| **Meta_EAS** | EAS | rs10305510 | | | | | | | | | | | | | | | |
| **Meta_EAS** | EAS | rs429358 | | | | | | | | | | | | | | | |
| **Meta_EAS** | EAS | rs6232 | | | | | | | | | | | | | | | |
| **Meta_EAS** | EAS | rs6235 | -0.01 | 0.05 | 0.87 | 0.74 | 4.40 | 0.87 | 0.00 | 0.05 | 0.95 | -1.57 | 1.40 | 0.26 | -0.01 | 0.05 | 0.84 |
| **Meta_EAS** | EAS | Baseline | | | | | | | 0.99 | 0.03 | 0.00 | -0.46 | 1.01 | 0.65 | 0.02 | 0.04 | 0.69 |
| **Meta_EUR** | EUR | BMI PGS | 0.00 | 0.01 | 0.86 | 0.30 | 0.66 | 0.65 | 0.31 | 0.01 | 0.00 | -2.24 | 0.29 | 0.00 | -0.02 | 0.01 | 0.09 |
| **Meta_EUR** | EUR | T2D PGS | 0.00 | 0.01 | 0.52 | -0.26 | 0.70 | 0.72 | 0.22 | 0.01 | 0.00 | -2.24 | 0.28 | 0.00 | -0.02 | 0.01 | 0.06 |
| **Meta_EUR** | EUR | rs10305492 | 0.05 | 0.05 | 0.30 | -5.12 | 5.58 | 0.36 | 0.24 | 0.01 | 0.00 | -2.30 | 0.32 | 0.00 | -0.03 | 0.01 | 0.03 |
| **Meta_EUR** | EUR | rs146868158 | -0.04 | 0.23 | 0.87 | 7.04 | 25.66 | 0.78 | 0.89 | 0.03 | 0.00 | -1.68 | 1.25 | 0.18 | -0.02 | 0.05 | 0.67 |
| **Meta_EUR** | EUR | rs6923761 | 0.02 | 0.01 | 0.11 | -1.25 | 1.03 | 0.23 | 0.32 | 0.01 | 0.00 | -2.23 | 0.29 | 0.00 | -0.02 | 0.01 | 0.08 |
| **Meta_EUR** | EUR | rs1042044 | 0.00 | 0.01 | 0.59 | 0.04 | 0.94 | 0.96 | 0.31 | 0.01 | 0.00 | -2.20 | 0.28 | 0.00 | -0.02 | 0.01 | 0.13 |
| **Meta_EUR** | EUR | rs10305420 | 0.01 | 0.01 | 0.46 | -0.81 | 1.15 | 0.48 | 0.53 | 0.01 | 0.00 | -2.37 | 0.34 | 0.00 | -0.02 | 0.02 | 0.18 |
| **Meta_EUR** | EUR | rs3765467 | 0.00 | 0.08 | 0.97 | 1.78 | 7.49 | 0.81 | 0.27 | 0.01 | 0.00 | -2.33 | 0.33 | 0.00 | -0.03 | 0.01 | 0.02 |
| **Meta_EUR** | EUR | rs10305421 | 0.02 | 0.04 | 0.59 | 0.60 | 3.91 | 0.88 | 0.36 | 0.01 | 0.00 | -2.26 | 0.30 | 0.00 | -0.03 | 0.01 | 0.06 |
| **Meta_EUR** | EUR | rs10305510 | | | | 200.78 | 859.71 | 0.82 | 0.89 | 0.03 | 0.00 | -1.70 | 1.24 | 0.17 | -0.02 | 0.05 | 0.65 |
| **Meta_EUR** | EUR | rs429358 | -0.01 | 0.01 | 0.44 | 1.53 | 1.60 | 0.34 | 0.19 | 0.01 | 0.00 | -2.35 | 0.32 | 0.00 | -0.03 | 0.01 | 0.08 |
| **Meta_EUR** | EUR | rs6232 | 0.00 | 0.02 | 0.91 | -0.02 | 2.43 | 0.99 | 0.34 | 0.01 | 0.00 | -2.23 | 0.28 | 0.00 | -0.02 | 0.01 | 0.12 |
| **Meta_EUR** | EUR | rs6235 | 0.01 | 0.01 | 0.17 | -1.22 | 1.01 | 0.23 | 0.33 | 0.01 | 0.00 | -2.24 | 0.28 | 0.00 | -0.02 | 0.01 | 0.12 |
| **Meta_EUR** | EUR | Baseline | | | | | | | 0.92 | 0.01 | 0.00 | -2.20 | 0.29 | 0.00 | -0.02 | 0.01 | 0.14 |
| **Meta_SAS** | SAS | BMI PGS | -0.05 | 0.05 | 0.31 | 4.37 | 4.40 | 0.32 | 0.91 | 0.05 | 0.00 | -3.32 | 1.94 | 0.09 | -0.11 | 0.07 | 0.11 |
| **Meta_SAS** | SAS | T2D PGS | 0.03 | 0.05 | 0.50 | -1.65 | 4.07 | 0.68 | 0.88 | 0.04 | 0.00 | -3.48 | 1.90 | 0.07 | -0.10 | 0.07 | 0.13 |
| **Meta_SAS** | SAS | rs10305492 | | | | | | | | | | | | | | | |
| **Meta_SAS** | SAS | rs146868158 | | | | | | | | | | | | | | | |

| | | | | | | | | | | | | | | | | | |
|---|---|---|---|---|---|---|---|---|---|---|---|---|---|---|---|---|---|
| **Meta_SAS** | SAS | rs6923761 | 0.12 | 0.08 | 0.14 | -10.84 | 6.60 | 0.10 | 0.84 | 0.05 | 0.00 | -3.30 | 2.00 | 0.10 | -0.12 | 0.07 | 0.08 |
| **Meta_SAS** | SAS | rs1042044 | 0.11 | 0.05 | 0.02 | -7.88 | 4.24 | 0.06 | 0.75 | 0.07 | 0.00 | -2.45 | 1.85 | 0.18 | -0.09 | 0.06 | 0.15 |
| **Meta_SAS** | SAS | rs10305420 | 0.00 | 0.08 | 0.95 | -1.94 | 6.48 | 0.76 | 0.88 | 0.05 | 0.00 | -3.58 | 1.93 | 0.06 | -0.10 | 0.07 | 0.14 |
| **Meta_SAS** | SAS | rs3765467 | 0.15 | 0.11 | 0.17 | -11.16 | 9.14 | 0.22 | 0.86 | 0.04 | 0.00 | -2.98 | 1.95 | 0.13 | -0.12 | 0.07 | 0.08 |
| **Meta_SAS** | SAS | rs10305421 | | | | | | | | | | | | | | | |
| **Meta_SAS** | SAS | rs10305510 | | | | | | | | | | | | | | | |
| **Meta_SAS** | SAS | rs429358 | | | | | | | | | | | | | | | |
| **Meta_SAS** | SAS | rs6232 | -0.32 | 0.29 | 0.28 | 21.08 | 21.82 | 0.33 | 0.88 | 0.04 | 0.00 | -3.22 | 1.96 | 0.10 | -0.11 | 0.07 | 0.11 |
| **Meta_SAS** | SAS | rs6235 | -0.14 | 0.07 | 0.04 | 9.25 | 5.98 | 0.12 | 0.96 | 0.05 | 0.00 | -3.63 | 1.84 | 0.05 | -0.09 | 0.06 | 0.17 |
| **Meta_SAS** | SAS | Baseline | | | | | | | 0.90 | 0.03 | 0.00 | -3.05 | 1.38 | 0.03 | -0.08 | 0.05 | 0.15 |
| **Meta_MID** | MID | BMI PGS | 0.02 | 0.02 | 0.17 | -0.87 | 1.59 | 0.59 | 0.46 | 0.02 | 0.00 | -5.16 | 0.83 | 0.00 | -0.03 | 0.04 | 0.38 |
| **Meta_MID** | MID | T2D PGS | 0.07 | 0.02 | 0.00 | -6.25 | 1.57 | 0.00 | 0.45 | 0.02 | 0.00 | -5.13 | 0.82 | 0.00 | -0.03 | 0.04 | 0.39 |
| **Meta_MID** | MID | rs10305492 | | | | | | | | | | | | | | | |
| **Meta_MID** | MID | rs146868158 | | | | | | | | | | | | | | | |
| **Meta_MID** | MID | rs6923761 | 0.05 | 0.02 | 0.05 | -4.54 | 2.31 | 0.05 | 0.41 | 0.03 | 0.00 | -4.94 | 0.83 | 0.00 | -0.03 | 0.04 | 0.34 |
| **Meta_MID** | MID | rs1042044 | 0.05 | 0.02 | 0.03 | -4.84 | 2.11 | 0.02 | 0.43 | 0.02 | 0.00 | -5.02 | 0.83 | 0.00 | -0.04 | 0.04 | 0.31 |
| **Meta_MID** | MID | rs10305420 | 0.02 | 0.02 | 0.45 | -1.46 | 2.13 | 0.50 | 0.45 | 0.03 | 0.00 | -4.99 | 0.83 | 0.00 | -0.04 | 0.04 | 0.28 |
| **Meta_MID** | MID | rs3765467 | 0.12 | 0.05 | 0.01 | -12.97 | 6.02 | 0.03 | 0.23 | 0.10 | 0.02 | -5.08 | 0.83 | 0.00 | -0.04 | 0.04 | 0.28 |
| **Meta_MID** | MID | rs10305421 | | | | | | | | | | | | | | | |
| **Meta_MID** | MID | rs10305510 | | | | | | | | | | | | | | | |
| **Meta_MID** | MID | rs429358 | 0.12 | 0.03 | 0.00 | -11.00 | 3.44 | 0.00 | 0.26 | 0.06 | 0.00 | -4.79 | 0.83 | 0.00 | -0.03 | 0.04 | 0.34 |
| **Meta_MID** | MID | rs6232 | 0.03 | 0.05 | 0.60 | -2.07 | 4.92 | 0.67 | 0.42 | 0.09 | 0.00 | -5.04 | 0.83 | 0.00 | -0.04 | 0.04 | 0.29 |
| **Meta_MID** | MID | rs6235 | -0.04 | 0.02 | 0.14 | 3.27 | 2.47 | 0.19 | 0.53 | 0.04 | 0.00 | -5.01 | 0.83 | 0.00 | -0.04 | 0.04 | 0.25 |
| **Meta_MID** | MID | Baseline | | | | | | | 0.47 | 0.02 | 0.00 | -5.04 | 0.83 | 0.00 | -0.04 | 0.04 | 0.28 |
| **Meta_analysis** | ALL | BMI PGS | 0.01 | 0.00 | 0.05 | 0.02 | 0.01 | 0.10 | 0.42 | 0.01 | 0.00 | 0.95 | 0.01 | 0.00 | -0.02 | 0.01 | 0.01 |
| **Meta_analysis** | ALL | T2D PGS | 0.01 | 0.00 | 0.05 | -0.92 | 0.43 | 0.03 | 0.53 | 0.00 | 0.00 | -1.66 | 0.20 | 0.00 | -0.02 | 0.01 | 0.01 |
| **Meta_analysis** | ALL | rs10305492 | 0.07 | 0.05 | 0.15 | -6.70 | 4.89 | 0.17 | 0.41 | 0.01 | 0.00 | -1.58 | 0.24 | 0.00 | -0.03 | 0.01 | 0.00 |
| **Meta_analysis** | ALL | rs146868158 | -0.04 | 0.23 | 0.87 | 7.04 | 25.66 | 0.78 | 0.89 | 0.03 | 0.00 | -1.68 | 1.25 | 0.18 | -0.02 | 0.05 | 0.67 |
| **Meta_analysis** | ALL | rs6923761 | 0.02 | 0.01 | 0.00 | -2.14 | 0.75 | 0.00 | 0.54 | 0.01 | 0.00 | -1.64 | 0.20 | 0.00 | -0.02 | 0.01 | 0.00 |
| **Meta_analysis** | ALL | rs1042044 | 0.01 | 0.01 | 0.26 | -0.50 | 0.59 | 0.40 | 0.49 | 0.01 | 0.00 | -1.62 | 0.20 | 0.00 | -0.02 | 0.01 | 0.01 |
| **Meta_analysis** | ALL | rs10305420 | 0.01 | 0.01 | 0.17 | -1.02 | 0.78 | 0.19 | 0.72 | 0.01 | 0.00 | -1.80 | 0.23 | 0.00 | -0.03 | 0.01 | 0.00 |
| **Meta_analysis** | ALL | rs3765467 | 0.09 | 0.02 | 0.00 | -7.40 | 2.44 | 0.00 | 0.56 | 0.01 | 0.00 | -1.75 | 0.22 | 0.00 | -0.03 | 0.01 | 0.00 |
| **Meta_analysis** | ALL | rs10305421 | 0.02 | 0.04 | 0.54 | -0.06 | 3.53 | 0.99 | 0.53 | 0.01 | 0.00 | -1.69 | 0.25 | 0.00 | -0.03 | 0.01 | 0.01 |
| **Meta_analysis** | ALL | rs10305510 | -0.04 | 0.04 | 0.36 | 5.63 | 4.32 | 0.19 | 0.84 | 0.01 | 0.00 | -0.66 | 0.33 | 0.05 | -0.03 | 0.01 | 0.03 |
| **Meta_analysis** | ALL | rs429358 | 0.01 | 0.01 | 0.45 | -0.71 | 1.22 | 0.56 | 0.14 | 0.01 | 0.00 | -2.18 | 0.26 | 0.00 | -0.02 | 0.01 | 0.04 |
| **Meta_analysis** | ALL | rs6232 | -0.01 | 0.02 | 0.45 | 0.99 | 1.73 | 0.57 | 0.56 | 0.00 | 0.00 | -1.73 | 0.20 | 0.00 | -0.03 | 0.01 | 0.00 |
| **Meta_analysis** | ALL | rs6235 | 0.01 | 0.01 | 0.18 | -0.73 | 0.69 | 0.29 | 0.52 | 0.01 | 0.00 | -1.65 | 0.20 | 0.00 | -0.02 | 0.01 | 0.01 |
| **Meta_analysis** | ALL | Baseline | | | | | | | 0.91 | 0.00 | 0.00 | -1.52 | 0.19 | 0.00 | -0.02 | 0.01 | 0.02 |

**Supplementary Table 17 | Meta analysis results for BS sensitivity analysis 6.**
Ancestry-specific and multi-ancestry meta-analyses of the BS sensitivity analysis: "Model using post-treatment weight as outcome and including interaction between baseline weight and each genetic exposure"

| | | | Interaction between main genetic exposure and baseline body weight | | | Main genetic exposure | | | Baseline body weight | | | Sex | | | Age at initiation | | |
|---|---|---|---|---|---|---|---|---|---|---|---|---|---|---|---|---|---|
| **Study** | **Ancestry** | **Model (main exposure)** | **Beta** | **Standard Error** | **P-value** | **Beta** | **Standard Error** | **P-value** | **Beta** | **Standard Error** | **P-value** | **Beta** | **Standard Error** | **P-value** | **Beta** | **Standard Error** | **P-value** |
| **Meta_AFR** | AFR | BMI PGS | 0.04 | 0.03 | 0.16 | -4.01 | 3.34 | 0.23 | 0.64 | 0.03 | 0.00 | -1.49 | 2.00 | 0.46 | 0.17 | 0.05 | 0.00 |
| **Meta_AFR** | AFR | T2D PGS | 0.03 | 0.03 | 0.36 | -1.86 | 3.78 | 0.62 | 0.62 | 0.04 | 0.00 | -1.71 | 1.98 | 0.39 | 0.17 | 0.06 | 0.00 |
| **Meta_AFR** | AFR | rs429358 | -0.04 | 0.05 | 0.35 | 6.84 | 5.98 | 0.25 | 0.72 | 0.04 | 0.00 | -0.07 | 2.72 | 0.98 | 0.17 | 0.08 | 0.03 |
| **Meta_AFR** | AFR | rs17702901 | -0.10 | 0.07 | 0.18 | 17.70 | 10.62 | 0.10 | 0.68 | 0.03 | 0.00 | -1.44 | 1.97 | 0.46 | 0.17 | 0.05 | 0.00 |
| **Meta_AFR** | AFR | rs728996 | -0.03 | 0.03 | 0.32 | 2.58 | 4.22 | 0.54 | 0.69 | 0.03 | 0.00 | -1.92 | 2.00 | 0.34 | 0.18 | 0.05 | 0.00 |
| **Meta_AFR** | AFR | Baseline | | | | | | | 0.69 | 0.02 | 0.00 | -1.26 | 1.91 | 0.51 | 0.17 | 0.05 | 0.00 |
| **Meta_EUR** | EUR | BMI PGS | 0.03 | 0.01 | 0.01 | -2.81 | 1.43 | 0.05 | 0.67 | 0.01 | 0.00 | -3.58 | 0.69 | 0.00 | 0.19 | 0.03 | 0.00 |
| **Meta_EUR** | EUR | T2D PGS | 0.01 | 0.01 | 0.44 | -0.89 | 1.75 | 0.61 | 0.68 | 0.01 | 0.00 | -3.34 | 0.68 | 0.00 | 0.20 | 0.03 | 0.00 |
| **Meta_EUR** | EUR | rs429358 | -0.03 | 0.03 | 0.21 | 4.57 | 3.37 | 0.17 | 0.67 | 0.02 | 0.00 | -3.72 | 0.76 | 0.00 | 0.17 | 0.03 | 0.00 |
| **Meta_EUR** | EUR | rs17702901 | -0.04 | 0.05 | 0.35 | 3.50 | 6.15 | 0.57 | 0.65 | 0.01 | 0.00 | -3.85 | 0.75 | 0.00 | 0.17 | 0.03 | 0.00 |
| **Meta_EUR** | EUR | rs728996 | 0.03 | 0.02 | 0.11 | -2.70 | 2.09 | 0.20 | 0.64 | 0.02 | 0.00 | -3.50 | 0.68 | 0.00 | 0.19 | 0.03 | 0.00 |
| **Meta_EUR** | EUR | Baseline | | | | | | | 0.67 | 0.01 | 0.00 | -2.97 | 0.67 | 0.00 | 0.20 | 0.03 | 0.00 |
| **Meta_AMR** | AMR | BMI PGS | 0.02 | 0.06 | 0.78 | -0.01 | 7.16 | 1.00 | 0.55 | 0.06 | 0.00 | -4.57 | 3.00 | 0.13 | 0.02 | 0.11 | 0.82 |
| **Meta_AMR** | AMR | T2D PGS | -0.04 | 0.05 | 0.49 | 5.01 | 6.13 | 0.41 | 0.55 | 0.06 | 0.00 | -5.00 | 3.02 | 0.10 | 0.04 | 0.11 | 0.74 |
| **Meta_AMR** | AMR | rs429358 | | | | | | | | | | | | | | | |
| **Meta_AMR** | AMR | rs17702901 | 0.09 | 0.10 | 0.39 | -6.79 | 11.75 | 0.56 | 0.51 | 0.07 | 0.00 | -4.69 | 3.00 | 0.12 | 0.01 | 0.11 | 0.91 |
| **Meta_AMR** | AMR | rs728996 | -0.01 | 0.09 | 0.95 | -0.27 | 10.63 | 0.98 | 0.56 | 0.13 | 0.00 | -5.36 | 3.00 | 0.07 | 0.03 | 0.11 | 0.75 |
| **Meta_AMR** | AMR | Baseline | | | | | | | 0.58 | 0.06 | 0.00 | -4.02 | 2.89 | 0.16 | 0.06 | 0.09 | 0.54 |
| **Meta_SAS** | SAS | BMI PGS | | | | | | | | | | | | | | | |
| **Meta_SAS** | SAS | T2D PGS | | | | | | | | | | | | | | | |
| **Meta_SAS** | SAS | rs429358 | | | | | | | | | | | | | | | |
| **Meta_SAS** | SAS | rs17702901 | | | | | | | | | | | | | | | |
| **Meta_SAS** | SAS | rs728996 | | | | | | | | | | | | | | | |
| **Meta_SAS** | SAS | Baseline | | | | | | | | | | | | | | | |
| **Meta_MID** | MID | BMI PGS | 0.06 | 0.02 | 0.00 | -4.52 | 1.89 | 0.02 | 0.47 | 0.02 | 0.00 | -3.70 | 1.00 | 0.00 | 0.16 | 0.04 | 0.00 |
| **Meta_MID** | MID | T2D PGS | 0.03 | 0.02 | 0.10 | -2.51 | 1.88 | 0.18 | 0.51 | 0.02 | 0.00 | -3.23 | 1.01 | 0.00 | 0.16 | 0.04 | 0.00 |
| **Meta_MID** | MID | rs429358 | 0.03 | 0.05 | 0.52 | -3.77 | 5.17 | 0.47 | 0.46 | 0.09 | 0.00 | -3.12 | 1.01 | 0.00 | 0.16 | 0.04 | 0.00 |
| **Meta_MID** | MID | rs17702901 | 0.01 | 0.08 | 0.93 | -1.46 | 7.97 | 0.85 | 0.50 | 0.16 | 0.00 | -3.17 | 1.01 | 0.00 | 0.16 | 0.04 | 0.00 |
| **Meta_MID** | MID | rs728996 | -0.05 | 0.03 | 0.03 | 5.51 | 2.65 | 0.04 | 0.59 | 0.04 | 0.00 | -3.13 | 1.01 | 0.00 | 0.16 | 0.04 | 0.00 |
| **Meta_MID** | MID | Baseline | | | | | | | 0.52 | 0.02 | 0.00 | -3.15 | 1.01 | 0.00 | 0.16 | 0.04 | 0.00 |
| **Meta_analysis** | ALL | BMI PGS | 0.04 | 0.01 | 0.00 | -3.41 | 1.07 | 0.00 | 0.62 | 0.01 | 0.00 | -3.49 | 0.54 | 0.00 | 0.18 | 0.02 | 0.00 |
| **Meta_analysis** | ALL | T2D PGS | 0.02 | 0.01 | 0.10 | -1.41 | 1.19 | 0.24 | 0.63 | 0.01 | 0.00 | -3.24 | 0.54 | 0.00 | 0.18 | 0.02 | 0.00 |
| **Meta_analysis** | ALL | rs429358 | -0.02 | 0.02 | 0.25 | 2.95 | 2.55 | 0.25 | 0.67 | 0.01 | 0.00 | -3.34 | 0.59 | 0.00 | 0.17 | 0.02 | 0.00 |
| **Meta_analysis** | ALL | rs17702901 | -0.03 | 0.03 | 0.33 | 3.04 | 4.14 | 0.46 | 0.65 | 0.01 | 0.00 | -3.46 | 0.57 | 0.00 | 0.16 | 0.02 | 0.00 |
| **Meta_analysis** | ALL | rs728996 | 0.00 | 0.01 | 0.88 | 0.70 | 1.51 | 0.64 | 0.64 | 0.02 | 0.00 | -3.34 | 0.53 | 0.00 | 0.18 | 0.02 | 0.00 |
| **Meta_analysis** | ALL | Baseline | | | | | | | 0.63 | 0.01 | 0.00 | -2.92 | 0.53 | 0.00 | 0.18 | 0.02 | 0.00 |

**Supplementary Table 18 | Meta analysis results for GLP1-RA sensitivity analysis 7.**
Ancestry-specific and multi-ancestry meta-analyses of the GLP1-RA sensitivity analysis: "excluding baseline weight W0 from statistical model".

| | | | Main genetic exposure | | | Sex | | | Age at initiation | | |
|---|---|---|---|---|---|---|---|---|---|---|---|
| **Study** | **Ancestry** | **Model (main exposure)** | **Beta** | **Standard Error** | **P-value** | **Beta** | **Standard Error** | **P-value** | **Beta** | **Standard Error** | **P-value** |
| **Meta_AFR** | AFR | BMI PGS | -0.14 | 0.21 | 0.51 | -0.35 | 0.44 | 0.42 | -0.01 | 0.02 | 0.63 |
| **Meta_AFR** | AFR | T2D PGS | 0.38 | 0.22 | 0.09 | -0.30 | 0.44 | 0.50 | -0.01 | 0.02 | 0.74 |
| **Meta_AFR** | AFR | Baseline | | | | -0.18 | 0.43 | 0.68 | -0.01 | 0.02 | 0.50 |
| **Meta_AMR** | AMR | BMI PGS | -0.11 | 0.19 | 0.57 | -0.47 | 0.41 | 0.25 | -0.02 | 0.02 | 0.32 |
| **Meta_AMR** | AMR | T2D PGS | 0.09 | 0.20 | 0.66 | -0.46 | 0.41 | 0.26 | -0.01 | 0.02 | 0.39 |
| **Meta_AMR** | AMR | Baseline | | | | -0.48 | 0.40 | 0.24 | -0.01 | 0.02 | 0.49 |
| **Meta_EAS** | EAS | BMI PGS | 0.09 | 0.72 | 0.90 | -1.48 | 1.39 | 0.29 | -0.02 | 0.05 | 0.78 |
| **Meta_EAS** | EAS | T2D PGS | 1.19 | 0.67 | 0.08 | -1.67 | 1.34 | 0.21 | -0.01 | 0.05 | 0.88 |
| **Meta_EAS** | EAS | Baseline | | | | -1.71 | 1.23 | 0.17 | -0.03 | 0.05 | 0.55 |
| **Meta_EUR** | EUR | BMI PGS | -0.11 | 0.13 | 0.39 | -1.61 | 0.26 | 0.00 | 0.00 | 0.01 | 0.70 |
| **Meta_EUR** | EUR | T2D PGS | 0.02 | 0.14 | 0.88 | -1.61 | 0.26 | 0.00 | 0.00 | 0.01 | 0.80 |
| **Meta_EUR** | EUR | Baseline | | | | -1.62 | 0.26 | 0.00 | -0.01 | 0.01 | 0.65 |
| **Meta_SAS** | SAS | BMI PGS | -0.38 | 0.89 | 0.67 | -2.97 | 2.03 | 0.14 | -0.09 | 0.07 | 0.20 |
| **Meta_SAS** | SAS | T2D PGS | 1.03 | 0.86 | 0.23 | -2.82 | 1.99 | 0.16 | -0.07 | 0.07 | 0.32 |
| **Meta_SAS** | SAS | Baseline | | | | -3.10 | 1.41 | 0.03 | -0.07 | 0.06 | 0.21 |
| **Meta_MID** | MID | BMI PGS | 0.16 | 0.33 | 0.63 | -0.26 | 0.71 | 0.71 | 0.01 | 0.03 | 0.74 |
| **Meta_MID** | MID | T2D PGS | 0.06 | 0.39 | 0.88 | -0.26 | 0.71 | 0.71 | 0.01 | 0.03 | 0.74 |
| **Meta_MID** | MID | Baseline | | | | -0.26 | 0.71 | 0.71 | 0.01 | 0.03 | 0.74 |
| **Meta_analysis** | ALL | BMI PGS | -0.10 | 0.09 | 0.30 | -1.06 | 0.19 | 0.00 | -0.01 | 0.01 | 0.29 |
| **Meta_analysis** | ALL | T2D PGS | 0.14 | 0.10 | 0.14 | -1.05 | 0.19 | 0.00 | -0.01 | 0.01 | 0.43 |
| **Meta_analysis** | ALL | Baseline | | | | -1.05 | 0.18 | 0.00 | -0.01 | 0.01 | 0.26 |

**Supplementary Table 19 | Meta analysis results for BS sensitivity analysis 7.**
Ancestry-specific and multi-ancestry meta-analyses of the BS sensitivity analysis: "excluding baseline weight W0 from statistical model".

| | | | Main genetic exposure | | | Sex | | | Age at initiation | | |
|---|---|---|---|---|---|---|---|---|---|---|---|
| **Study** | **Ancestry** | **Model (main exposure)** | **Beta** | **Standard Error** | **P-value** | **Beta** | **Standard Error** | **P-value** | **Beta** | **Standard Error** | **P-value** |
| **Meta_AFR** | AFR | BMI PGS | 0.09 | 0.61 | 0.88 | -0.19 | 1.54 | 0.90 | 0.19 | 0.05 | 0.00 |
| **Meta_AFR** | AFR | T2D PGS | 1.11 | 0.73 | 0.13 | -0.22 | 1.54 | 0.89 | 0.20 | 0.05 | 0.00 |
| **Meta_AFR** | AFR | Baseline | | | | -0.58 | 1.48 | 0.70 | 0.18 | 0.04 | 0.00 |
| **Meta_AMR** | AMR | BMI PGS | 1.69 | 0.96 | 0.08 | -1.39 | 2.50 | 0.58 | 0.14 | 0.09 | 0.13 |
| **Meta_AMR** | AMR | T2D PGS | 0.65 | 0.98 | 0.50 | -1.92 | 2.52 | 0.44 | 0.15 | 0.09 | 0.10 |
| **Meta_AMR** | AMR | Baseline | | | | -1.01 | 2.39 | 0.67 | 0.13 | 0.08 | 0.12 |
| **Meta_EUR** | EUR | BMI PGS | -0.01 | 0.22 | 0.95 | -0.82 | 0.49 | 0.09 | 0.20 | 0.02 | 0.00 |
| **Meta_EUR** | EUR | T2D PGS | 0.22 | 0.25 | 0.38 | -0.83 | 0.49 | 0.09 | 0.21 | 0.02 | 0.00 |
| **Meta_EUR** | EUR | Baseline | | | | -1.05 | 0.60 | 0.08 | 0.24 | 0.03 | 0.00 |
| **Meta_SAS** | SAS | BMI PGS | | | | | | | | | |
| **Meta_SAS** | SAS | T2D PGS | | | | | | | | | |
| **Meta_SAS** | SAS | Baseline | | | | | | | | | |
| **Meta_MID** | MID | BMI PGS | 0.43 | 0.40 | 0.28 | 0.51 | 0.83 | 0.54 | 0.11 | 0.04 | 0.01 |
| **Meta_MID** | MID | T2D PGS | 0.14 | 0.45 | 0.76 | 0.47 | 0.83 | 0.57 | 0.11 | 0.04 | 0.01 |
| **Meta_MID** | MID | Baseline | | | | 0.47 | 0.83 | 0.57 | 0.11 | 0.04 | 0.01 |
| **Meta_analysis** | ALL | BMI PGS | 0.15 | 0.18 | 0.42 | -0.48 | 0.40 | 0.23 | 0.18 | 0.02 | 0.00 |
| **Meta_analysis** | ALL | T2D PGS | 0.30 | 0.21 | 0.15 | -0.51 | 0.40 | 0.20 | 0.19 | 0.02 | 0.00 |
| **Meta_analysis** | ALL | Baseline | | | | -0.54 | 0.46 | 0.23 | 0.20 | 0.02 | 0.00 |

**Supplementary Table 20 | Bariatric suregry (BS) codes.**
List and description of the procedures included in the BS analysis. Procedure, abbreviations, techinque description and CPT-4 Codes are reported in the table.

| Procedure | Abbreviation | Technique | CPT-4 Codes |
|---|---|---|---|
| Roux-en-Y gastric bypass | RYGB | Open<br>Laparoscopic | 43846, 43847<br>43644, 43645, |
| Sleeve gastrectomy | SG | Laparoscopic | 43775, 43659 |
| Adjustable gastric band | AGB | Laparoscopic | 43843, S2082 |
| Adjustable gastric band | AGB | Open | 43770 |
| Vertical banded gastroplasty | VBG | Open | 43842 |
| Biliopancreatic diversion with duodenal switch | BPD/DS | Open | 43845 |

**Supplementary Table 21 | Proportions of bariatric suregry (BS) procedures across cohorts.**
List and description of the procedures included in the BS analysis.
Procedure, abbreviations, CPT-4 Codes and proportions of the respective procedures are reported in the table.

| Procedure | | | Proportion (in %) | | | | | | | Weighted proportion (in %) |
|---|---|---|---|---|---|---|---|---|---|---|
| **Procedure** | **Abbreviation** | **CPT-4 Codes** | **AoU** | **BBSS** | **BioMe** | **EstBB** | **HUS** | **MGBB** | **UKBB** | **QBB** | **All cohorts** |
| Roux-en-Y gastric bypass | RYGB | 43846, 43847 43644, 43645, | 31.5 | 4.2 | 25.9 | 56.0 | 27.2 | 29.2 | 58.2 | 96.0 | **52.3** |
| Sleeve gastrectomy | SG | 43775, 43659 | 64.6 | 95.8 | 67.8 | 40.0 | 71.2 | 66.6 | 23.5 | 2.0 | **43.6** |
| Other (incl. Adjustable gastric band, vertical banded gastroplasty, biliopancreatic diversion with duodenal switch) | AGB, VBG, BPD/DS | 43843, S2082, 43770, 43842, 43845 | 3.9 | 0.0 | 6.3 | 4.0 | 1.6 | 4.2 | 18.3 | 2.0 | **4.1** |
